# A Multimodal Framework for Organ- and Cell-Resolved Biological Aging and Longevity Intervention Discovery

**DOI:** 10.64898/2026.05.08.26352759

**Authors:** Saleem A. Al Dajani, John R. Williams, Matias Fuentealba, Ting Zhai, David Furman, Michael P. Snyder, Omar O. Abudayyeh, Jonathan S. Gootenberg, Vadim N. Gladyshev

## Abstract

Aging is the primary driver of chronic disease and mortality, requiring comprehensive frameworks for quantification of aging and nomination of longevity interventions. We developed mAge (multimodal age), a biological aging framework that integrates plasma proteomics, wearables, and mortality hazard to predict biological age, intrinsic capacity, and mortality risk. By combining proteomic and wearable data in UK Biobank samples, mAge exceeds unimodal baseline age prediction to 0.87 test R² and 2.3 years mean error, and reduces unimodal baseline mortality prediction error by 21%. We further constructed organ-and cell type-specific biological clocks that quantify aging across 49 distinct subsystems, revealing that cardiac, immune, and intracellular protein signatures benefit most from wearable integration. By mapping data to FDA-approved drug targets, we identified interventions, such as GLP-1 receptor agonists, gabapentin, and ACE inhibitors, that are associated with lower overall and subsystem-specific proteomic age and mortality risk or are associated with longer time-to-death and later age-at-death in longitudinal and deceased cohorts. mAge establishes a scalable framework for nominating and validating personalized longevity interventions, bridging continuous digital monitoring with molecular aging diagnostics.

## Introduction

Age is the dominant risk factor for chronic disease^1^, and the rate of multimorbidity accumulation itself accelerates with age, as each additional year carries a greater disease-onset hazard than the last. This compounding trajectory demands detection of aging-associated functional decline before diagnosable pathology emerges. The WHO and FDA operationalize this through intrinsic capacity (IC; ICD-11 MG2A)^2–11^, the composite of locomotion, cognition, psychological well-being, vitality, and sensory function. DNA methylation (DNAm)-derived IC scores correlate with all-cause mortality more strongly than either chronological age or molecular aging markers alone^12^, establishing IC as a diagnostically valid proxy for age, loss of function, and mortality, making it a tractable target for longevity interventions.

Current clinical diagnostics are effective at identifying overt pathology but do not detect the latent physiological decline that precedes chronic disease^1^. Just as a computer’s activity monitor reveals not only that a system is slow but precisely which process is consuming resources, a clinically actionable aging diagnostic must identify not only that a patient is aging, but which biological subsystems are failing and why. Despite this need, no such framework exists^13–16^. A cancer patient weighing treatment options can be given a population-level survival estimate post-surgery^17^, but not tissue-resolved biological states or ranked personalized interventions. Early interventions that are simple and affordable, such as diet, lifestyle, and supplements, can decelerate age-related disease onset^2^ before treatment becomes expensive and palliative, but acting early demands understanding *where* aging in the body proceeds more rapidly.

Biological aging clocks, which quantify various patterns of aging, have advanced from first-generation epigenetic predictors^18–20^ of chronological age to second-generation biomarkers that quantify phenotypic changes and future mortality and third-generation biomarkers that quantify the rate of aging. Another important advance in the field is proteomic aging clocks quantifying age or mortality at organismal or organ-specific levels, with plasma proteomic models achieving r>0.9 with chronological age^21–25^. However, existing organ clocks rely on single expression databases, enforce exclusive protein-to-organ assignment that discards cross-tissue signals, and optimize mean error rather than risk ranking^24^, but they demonstrate that heterogeneous tissue-specific age gaps^22,23^ associate with incident disease^25,26^, which motivates associating proteomic age gaps with IC and functional decline in this study.

Across modalities, current clocks trade off cost, invasiveness, temporal resolution, and explainability: epigenetic and proteomic assays are higher in cost, discretely sampled, and molecularly grounded, whereas wearables are lower in cost, non-invasive, and continuous but molecularly ungrounded. Wearable-derived aging models have advanced in parallel, with accelerometer-based deep-learning predictors estimating biological age and mortality risk from continuous activity, sleep, and circadian signals^27^, and movement-intensity profiles in UK Biobank (UKB) wrist accelerometry independently predicting all-cause mortality^28–31^. Unlike discretely sampled molecular assays, wearables provide a continuous functional readout of the body and its subsystems—cardiovascular load, energy expenditure, sleep architecture, and circadian rhythm—capturing dynamic physiological state that is largely orthogonal to plasma proteomic snapshots and contributing inputs to a biological age predictor that proteomics alone cannot recover. Despite this complementarity, proteomics and wearables have not been jointly modeled. Here, we introduce mAge (multimodal age), a framework that integrates continuous wearable physiological signals with periodic plasma proteomics and data on mortality to construct overall and subsystem-resolved biological activity monitors that identify which subsystems are aging, trace the responsible molecular processes, and link them to druggable targets.

Further advances in biomarkers of aging are critically needed for identifying interventions that target aging. Therapeutically, the NIA Intervention Testing Program (ITP) reported increases in lifespan of mice for, among others, aspirin and captopril (ACE inhibitor)^32,33^, but human translation remains restricted to observational studies^34,35^. While the hallmarks of aging^13,14,16^ provide a framework linking biomarkers to shared cellular processes, these modules obscure the system-level coupling^4^ that continuous multimodal monitoring is uniquely positioned to capture.

We address these limitations with mAge by integrating proteomics, wearables, and mortality features with advanced subsystem^36–38^ and drug-target^39^ mappings, coupled to a longitudinal drug-nomination pipeline validated against mouse ITP outcomes and human UKB data. The immediate objective is a validated multimodal framework that predicts age, mortality, IC, and comorbidities with accuracy exceeding current single-modality state-of-the-art (SOTA), and that identifies FDA-approved drugs associated with lower overall and subsystem-specific biological age in observational data. The broader objective is to deploy mAge as the inference backbone of a virtual human intervention model capable of nominating personalized longevity interventions^40–42^ before experimental verification, completing the process of translating health predictions into real-world validation through continuous wearable monitoring and affordable at-home testing. Whether the forces accelerating biological aging trajectories can be identified and counteracted to decelerate, arrest, or partially reverse biological age is the central question this work addresses.

## Results

### mAge predicts age, mortality, morbidities, and loss of function

To determine whether biological aging can be resolved into interpretable and actionable subsystem-specific processes, we developed mAge as a multimodal framework that integrates plasma proteomics, wearable-derived physiology, mortality risk, IC, and comorbidity outcomes. We applied this approach to UK Biobank data comprising Olink plasma proteomics, wrist accelerometry, longitudinal diagnosis records, mortality data, and repeated proteomic measurements, and linked the resulting protein features to organs, tissues, cell types, and FDA-approved drug targets.

We classified the Olink panel of 2,922 plasma proteins assayed in UK Biobank participants by Human Protein Atlas (HPA) mapping, secretion status, blood detectability, druggable-genome inclusion, and FDA-approved drug-target status (Fig. 1a; detailed coverage in Supplementary Fig. 1 and protein-flow Sankey in Supplementary Fig. 2), and examined the multimodal cohorts spanning proteomics, accelerometry, and mortality follow-up with overlapping subsets (Fig. 1b; full Venn counts in Supplementary Fig. 3; cohort demographics in Supplementary Fig. 4). This panel-level annotation reveals strong overlap across the three independently curated annotation sets (HPA atlas mapping, the druggable genome, and FDA-approved drug targets) with 98.7% of panel proteins indexed in HPA, 41.2% in the druggable genome, and 9.4% (n=276) correspond to FDA-approved drug targets, of which 33.7% are classified as secreted (Supplementary Fig. 1–2). The substantial fraction of secreted, druggable, and approved targets present in plasma supports the use of this panel for both subsystem mapping and intervention nomination based on overall subsystem-specific clocks.

**Figure 1.**
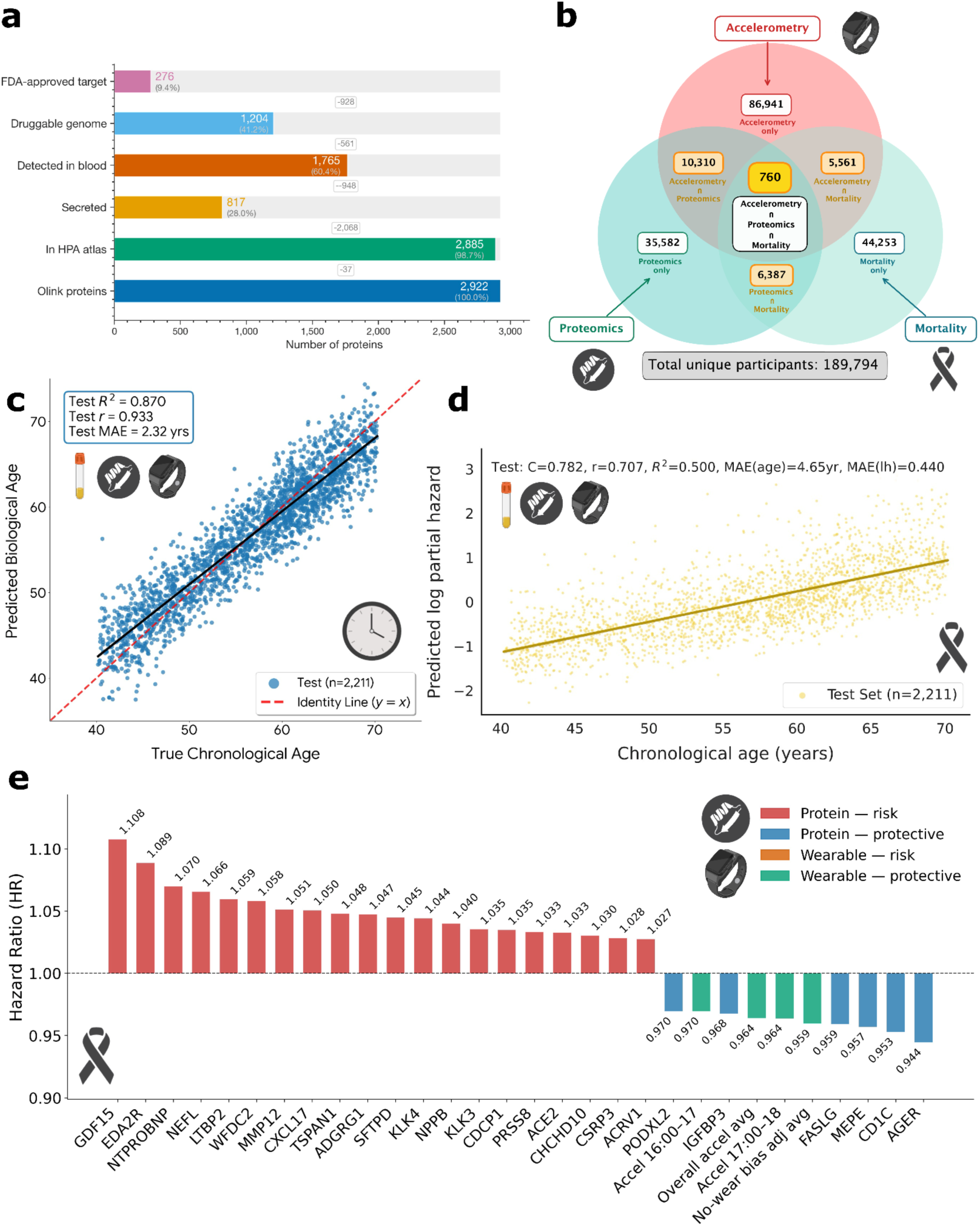
Multimodal age and mortality models with associated features. (a) Annotation of the Olink proteomic panel (n=2,922) by HPA atlas membership, secretion status, blood detectability, druggable genome inclusion, and FDA-approved drug-target status. (b) Overlapping numbers of UK Biobank participants across the proteomics, accelerometry, and mortality modalities. (c) Multimodal (proteomics + wearables) chronological age model on the test set. (d) Multimodal mortality model on the test set, with C-index, correlation, R², and MAE for both predicted log partial hazard and predicted age. (e) Top proteomic (red = risk, blue = protective) and wearable (orange = risk, green = protective) features associated with mortality risk by hazard ratio.

First, we investigated the predictive performance of models trained on different combinations of these modalities. Our unimodal model trained on plasma proteomics predicts proteomic age based on chronological age with a test R² of 86.3% and MAE of 2.4 years, similar to other studies^21–25,43^ (Extended Data Fig. 1a; sex-stratified scatters in Supplementary Fig. 5). Multimodally, mAge improves to 87% accuracy with an error of 2.3 years when plasma proteomics is combined with wearable and mortality-derived modalities (Fig. 1c; per-modality feature contributions in Supplementary Fig. 6), despite the training cohort being reduced approximately 5-fold because of limited participant overlap across modalities. Likewise, our unimodal model trained on plasma proteomics predicts mortality risk from chronological age^24^ with a Pearson correlation of 0.69 and MAE of 4.83 years (0.55 in log partial hazard; Extended Data Fig. 1b). Incorporating multimodal inputs modestly improves calibration-oriented metrics, including a 4% increase in R², a 21.4% reduction in MAE for log hazard, and a 3.7% reduction in MAE for time-to-death, yielding a Pearson correlation of 0.71 with an error of 4.65 years and 0.44 in predicted log partial hazard. These gains suggest that wearable-derived physiology contributes complementary information that refines individualized mortality-risk estimation beyond plasma proteomics alone. Held-out performance, however, decreases slightly from a C-index of 0.81 in the unimodal model to 0.78 in the multimodal model, likely reflecting the substantially smaller subset of participants with both proteomic and wearable data available for training and evaluation (Fig. 1d; full predicted time-to-death scatters with and without wearables in Supplementary Fig. 7; mortality-feature contributions by modality in Supplementary Fig. 8). Mortality risk separation between future decedents and survivors also becomes progressively more pronounced with increasing age at recruitment, whereas variability within age groups is driven more strongly by population heterogeneity than by proximity to death itself (Extended Data Fig. 2). Moreover, integrating wearable-derived features into the proteomic mortality framework^27^ (Fig. 1e) reshapes both risk-associated and protective signatures, indicating that behavioral and physiological dynamics captured by wearables provide information that is not fully represented in circulating proteomic profiles alone.

On the risk axis, the multimodal Cox model remains anchored by canonical aging factors, including GDF15, the premier general mortality marker in longitudinal aging cohorts^44,45^, and EDA2R, a marker of cellular senescence and systemic aging^46,47^, alongside cardiac-load proteins (NTproBNP, NPPB) and the metabolic health marker IGFBP3 (Extended Data Fig. 1c–f). While wearable accelerometry features do not reach the protein-dominated top-25 risk threshold (HR = 1.024), they contribute significantly to the protective profile. The most robust wearable signals associated with reduced mortality are average acceleration adjusted for no-wear bias (HR = 0.959), acceleration during the 5–6 pm window (HR = 0.964), overall daily acceleration (HR = 0.964), and 4–5 pm acceleration (HR = 0.970). These findings align with evidence that afternoon moderate-intensity physical activity confers a survival advantage^28–31^. Conversely, only two wearable features are associated with increased risk (HR > 1): midnight (0–1 am; HR = 1.006) and early-morning (4–5 am; HR = 1.001) acceleration, which likely capture nocturnal activity linked to sleep fragmentation. Incorporating the wearable modality effectively bisects the aggregate model weighting previously assigned to the proteomic modality, resulting in the exclusion of 260 proteins with diminished predictive utility. This phenomenon is interpreted as a feature-level shrinkage effect rather than a decrement in biological signal: model capacity is reallocated toward the proteins harboring the most robust residual mortality information, specifically GDF15, EDA2R, NTproBNP, NPPB, and IGFBP3, and toward the orthogonal physiological data derived from wearables, facilitating a sparser, more interpretable framework while maintaining near-equivalent ranking accuracy. Blood collection times exhibited broad temporal coverage across participant visits and wearable acquisition hours, suggesting that the prioritized features are neither specific to temporally localized patterns of sample acquisition nor appreciably confounded by participant age or blood collection timing (Supplementary Fig. 9).

Next, we quantified intrinsic capacity, the WHO- and FDA-operationalized measure of functional decline (Table S1), as well as its five domains (locomotion, cognition, psychological well-being, vitality, and sensory; baseline domain and total-IC distributions shown in Fig. 2a–f). The model was most predictive of the vitality domain of IC (intrinsic vitality; Fig. 2g) with a test R² of 0.66 and correlation of 0.81 (operationalization differences from prior UK Biobank IC implementations summarized in Table S2), and is more predictive from proteomics in women (R²_test_ = 0.68, r = 0.83; Fig. 2h) than men (R²_test_ = 0.61, r = 0.78; Fig. 2i). The forecast horizon, the interval between baseline assessment and the final prediction target, ranges from 2.17 to 19.46 years (mean 12.14, median 13.43, IQR 9.52–15.26, SD 3.84) across 109,160 participants. Follow-up duration includes 7.1% of the cohort with < 5 years, 21.8% with 5–10 years, 41.5% with 10–15 years, and 29.5% with 15–20 years, placing the majority of participants (71.0%) within a 10–20 year window. Over this period, baseline features highly predicted future overall IC (R²_test_= 0.89) and domain-specific scores (R²_test_= 0.81–0.92; Fig. 2j), as well as prevalent disease burden (*AUC*_test_= 0.70–0.82) and incident disease onset (*AUC*_test_ = 0.62–0.79; Fig. 2k). These results show one of the longest forecasting horizons for proteomics-grounded IC and comorbidity prediction, demonstrating that baseline clinical and biomarker profiles can anticipate functional capacity over a decade in advance for the median participant. Together, these results establish mAge as a high-performance multimodal framework that can integrate molecular, physiological, functional, and survival information to quantify biological aging and forecast clinically relevant age-related outcomes.

**Figure 2.**
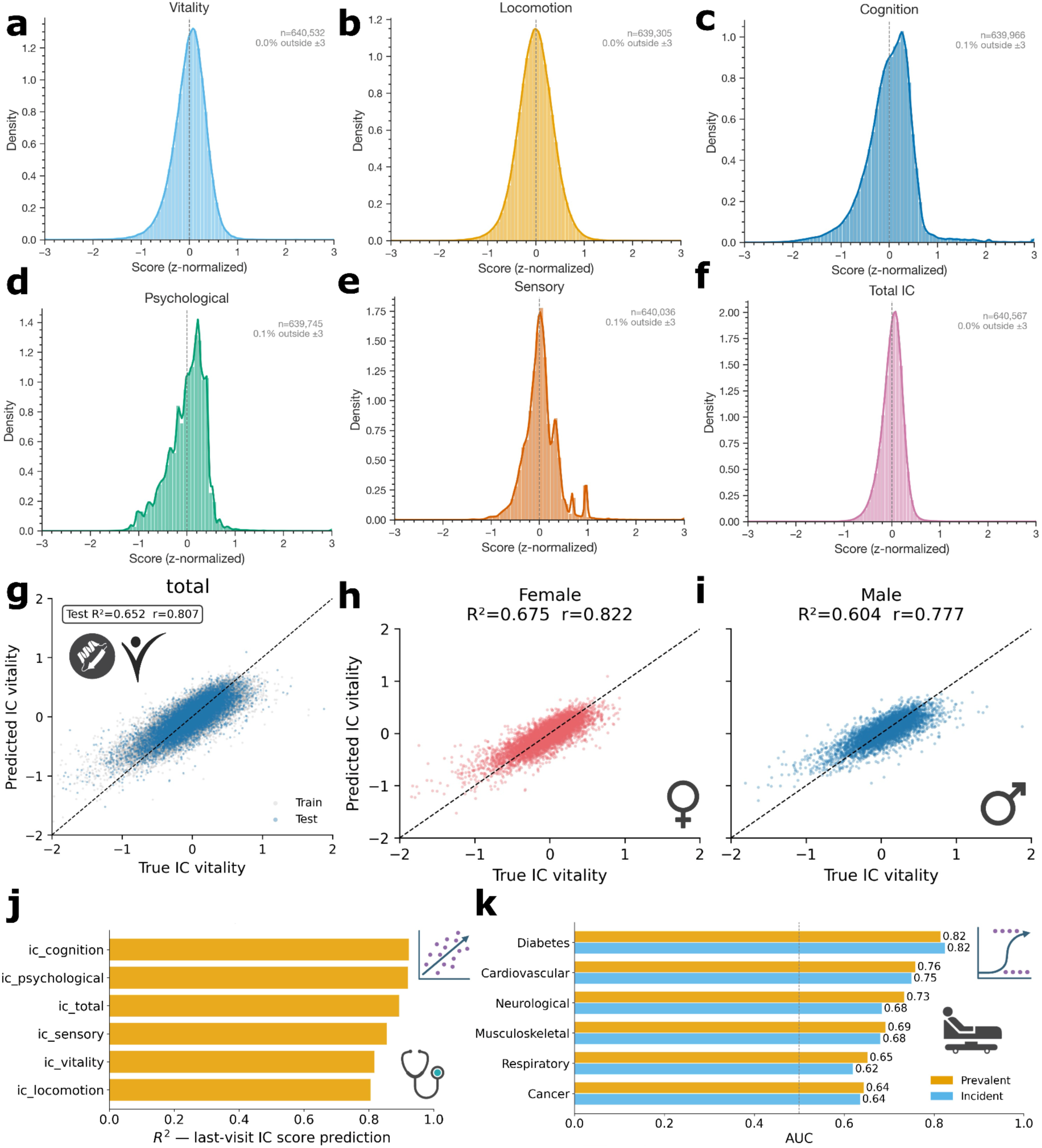
Predicting intrinsic capacity at baseline and forecasting future scores and onset of diseases. (a–f) Distributions of z-normalized intrinsic capacity domain scores across UK Biobank participants for vitality (a), locomotion (b), cognition (c), psychological (d), sensory (e), and total IC (f). (g) Proteomic intrinsic vitality model on train and test sets. (h) Sex-stratified intrinsic vitality models for female (pink) and (i) male (blue) participants. (j) Future last-visit prediction R² for total and domain-specific intrinsic capacity scores from baseline clinical and IC features. (k) Future comorbidity prediction (AUC) from baseline clinical and IC features for prevalent (orange) and incident (blue) disease across major disease categories.

### Linking mAge to origin cells, tissue, and organs enables model interpretability

We next leveraged the panel-level biological annotations to decompose predictive performance across tissue, cellular, secretory, and druggable subsystems. Having demonstrated that mAge effectively captures organism-level biological aging and clinical risk, we sought to determine the biological origins of these signals. We mapped plasma proteins to specific organs, tissues, secretome compartments, and cell types (Tab. S3), subsequently training subsystem-resolved clocks under two configurations: a proteomic age model and a multimodal mortality model that integrates wearable-derived physiological signals. This approach allows for the direct quantification of the marginal benefit provided by wearables across each biological subsystem (Fig. 3a), transforming mAge from a global predictor into an interpretable activity monitor of aging across distinct compartments (Fig. 3 overview; three-tier organelle–cell–body overview of age R^2^ and mortality C-index in Supplementary Fig. 10; tissue-resolved maps overlaid on anatomical templates in Supplementary Fig. 11–12).

**Figure 3.**
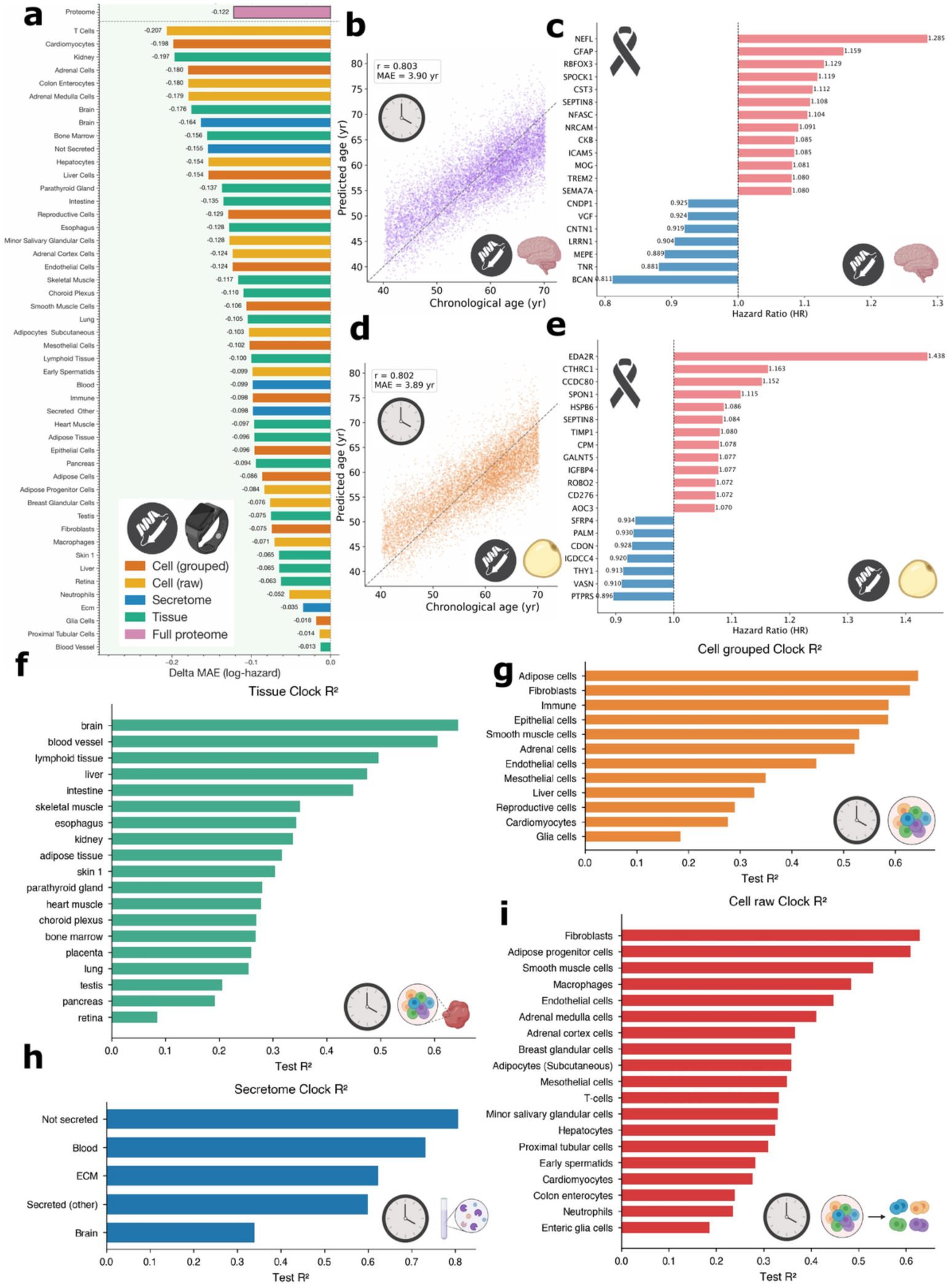
Organ/tissue-, secretome-, and cell-specific aging clock and mortality models based on proteomics and wearables, compared against the full-proteome reference. (a) Improvements in mortality models from adding wearable features by difference in MAE of log partial hazard, across tissue (green), grouped cell (orange), raw cell (yellow), secretome (blue), and full-proteome (pink) models. (b) Brain tissue-specific chronological age model and (c) top brain tissue-specific proteomic features associated with increased (red) and reduced (blue) mortality risk by hazard ratio. (d) Adipose cell-specific chronological age model and (e) top adipose cell-specific proteomic features associated with increased (red) and reduced (blue) mortality risk by hazard ratio. (f) Tissue-specific aging clock R²_test. (g) Grouped cell-specific aging clock R²_test. (h) Secretome-specific aging clock R²_test. (i) Raw cell-specific aging clock R²_test.

We first examined where wearables most significantly enhance subsystem mortality prediction. Wearable integration improved cardiac and immune subsystem models by up to 44% in log-hazard MAE and the intracellular ‘not secreted’ compartment by 28%. These gains likely reflect the complementary biological information captured by the two modalities, whereby intracellular proteins, often released during necrosis or programmed cell death, encode tissue damage–associated mortality signals^28–31^, while wearable-derived features capture orthogonal physiological and behavioral dynamics. Conversely, wearables provided negligible improvement to structural-protein-rich clocks (e.g., blood vessel and proximal tubular cell), suggesting that they capture dynamic physiological states rather than chronic structural deterioration. Detailed analysis of illustrative subsystems reveals that the brain tissue-specific age model achieves a test R^2^ of 0.64 and MAE of 3.9 years (Fig. 3b), with top mortality features (Fig. 3c) including markers of glial and neurovascular biology. Similarly, the adipose cell-specific model achieves R^2^ = 0.64 and MAE = 3.9 years (Fig. 3d), identifying metabolic regulators such as SEPTIN8 as drivers of mortality risk across multiple compartments (Fig. 3e).

Predictive performance differed substantially across subsystem panels, with the strongest signals consistently arising from mesenchymal, stromal, and immune-associated compartments. Across grouped and raw cell-specific analyses (Fig. 3g,i), fibroblasts, adipose progenitors, smooth muscle cells, and immune populations exhibited the highest predictive utility, while tissue-level analyses (Fig. 3f) highlighted vascularized and immune-enriched organs including brain, blood vessel, lymphoid tissue, liver, and intestine. This convergence suggests that fibroblasts and immune cells, being systemically distributed and responsive to inflammaging and senescence, provide a robust proteomic signal recoverable from plasma. In contrast, terminally differentiated or post-mitotic cells (cardiomyocytes, neurons, glia, early spermatids) occupied the bottom of every cell-level panel, likely due to slower turnover or aging mechanisms that do not map cleanly onto plasma signatures. Reproductive aging also appeared partially decoupled from somatic trajectories. The brain provided an instructive contrast: although bulk brain tissue performed well (Fig. 3f), glial and brain-secreted factors ranked lower (Fig. 3g,h), potentially reflecting the relative isolation of brain-specific cellular signals from circulating proteomic measurements, likely because the blood–brain barrier insulates specific cell types while bulk-tissue vasculature carries learnable systemic signals. The secretome analysis confirms that intracellular proteins predict best, followed by circulating and ECM proteins, mirroring established patterns in biological aging clocks. Subsystems that are systemically exposed and turn over rapidly age in predictable, learnable ways, whereas isolated or specialized programs exhibit more idiosyncratic trajectories. Full age–mortality scatters provided in Supplementary Fig. 13–15 and Extended Data Fig. 3a–d, and mortality risk is detailed in Extended Data Fig. 3e–f with organelle-level resolution.

While most proteins are subsystem-specific (Extended Data Fig. 4–5; Supplementary Fig. 16–17), certain metabolic-health markers, such as SEPTIN8 (binds to lipids and is a neurotransmitter and glucose uptake regulator), associate with increased mortality hazard across both the brain and adipose models. Despite fewer features per subsystem, adding wearables to these clocks reduces MAE in predicted log partial hazard by 22.4% (avg. 0.1) and time-to-death by 1.2% (avg. 0.07 years), while maintaining the ranking accuracy and correlation observed in proteomic-only models.

### mAge tracks biological aging longitudinally across subsystems

To determine whether mAge captures within-person changes over time, we next applied overall and subsystem-specific proteomic clocks to participants with repeated proteomic measurements. For 40 random participants with 2 or more visits with proteomic assays, we longitudinally tracked predicted age using mAge with the unimodal overall proteomic (Fig. 4a) and bimodal overall proteomic-wearable (Fig. 4b) clocks, with longitudinal trajectories for all 19 organ/tissue clocks shown in Extended Data Fig. 6 and for all 19 raw cell-type clocks in Extended Data Fig. 7. Distributions of predicted versus chronological age across visits for the same set of tissue clocks plus the overall proteome reference are shown in Extended Data Fig. 8. The maximum within-person slope is achieved with the overall unimodal and bimodal models, and the slopes vary across subsystems (Fig. 4c–e; complete within-person slope rankings across all clock variants—reference, tissue, secretome, grouped cell, and raw cell—in Supplementary Fig. 18). Overall unimodal and bimodal slopes are similar (0.6-0.7), while slopes are slightly higher when unimodal. Highest organ and tissue-specific slopes are for brain and blood vessels (0.4-0.5), highest secretome-specific slopes are for intracellular unsecreted proteins (0.6), highest grouped cell-specific slopes are for adipose, epithelial, and immune cells (0.44–0.5), and highest raw cell-specific slopes are for fibroblasts, adipose progenitors, and macrophages (0.35-0.5). Longitudinal tracking of mAge is best using the full proteome, and mAge varies in its ability to capture age variations over time per subsystem. These findings indicate that repeated mAge measurements can capture both organism-wide and subsystem-specific aging trajectories, providing a basis for longitudinal monitoring of biological age and for detecting compartment-specific responses to interventions.

**Figure 4.**
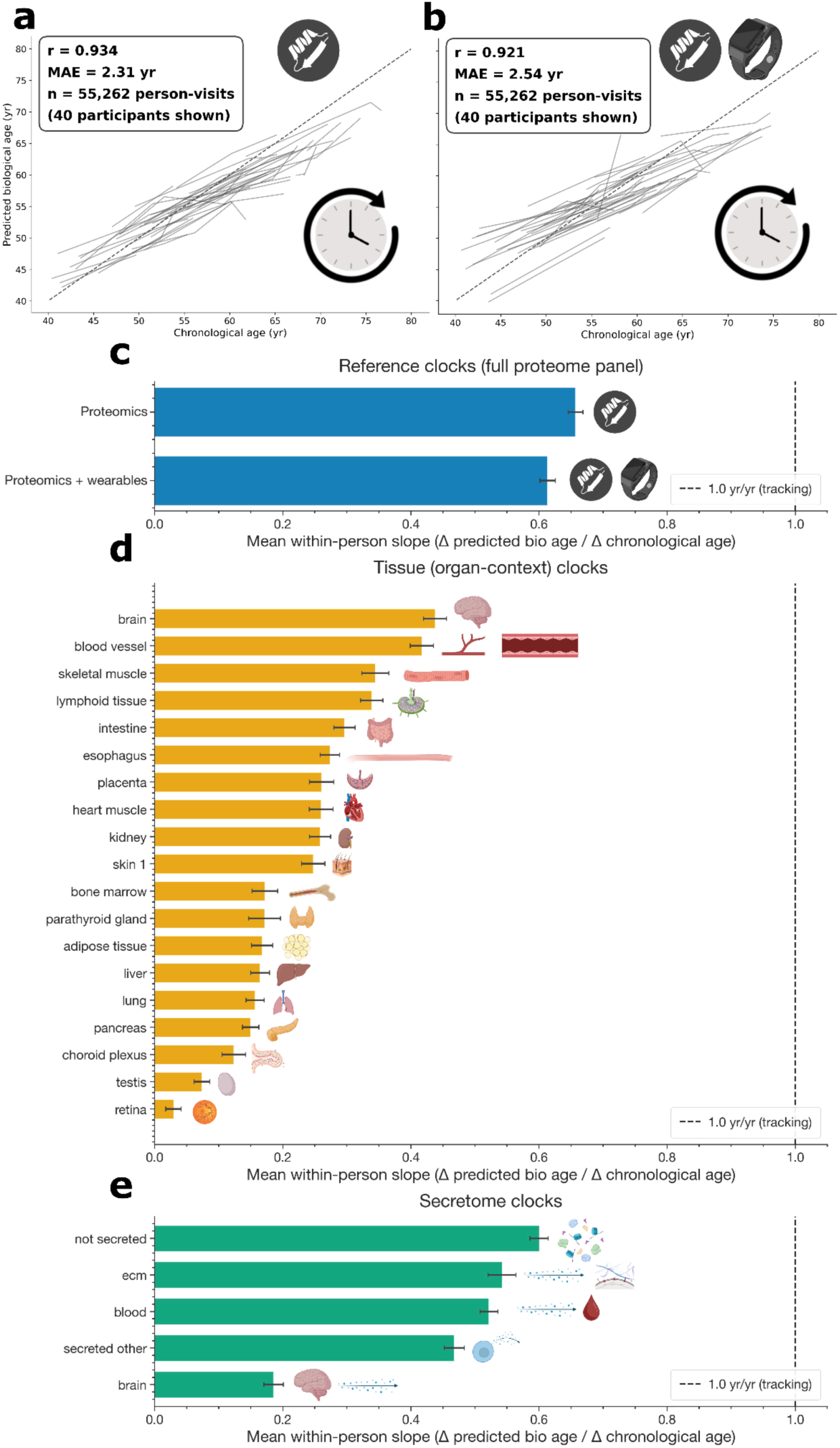
Proteomic aging clocks track age longitudinally, with within-person slopes that differ across organ/tissue and secretome subsystems. (a) Overall proteomic (unimodal) and (b) overall proteomic with wearables (bimodal) aging clock models applied longitudinally to a random subset of participants with ≥2 visits with proteomic assays (dotted diagonal is y=x, thin lines connect longitudinal visits within participants). (c) Mean within-person slopes (Δ predicted bio age / Δ chronological age) for the full-proteome reference clocks: proteomics-only and proteomics + wearables. (d) Mean within-person slopes for tissue (organ-context) clocks across organs/tissues. (e) Mean within-person slopes for secretome clocks (not secreted, ECM, blood, secreted other, brain). Dashed line at 1.0 yr/yr indicates perfect longitudinal tracking.

### Assessing intervention effects on overall and subsystem-specific biological age and mortality

Having established that mAge can quantify biological aging globally and across specific organs, tissues, and cell types, we next asked whether the proteins driving these clocks could be linked to existing pharmacological interventions. The total of 2922 proteins in the proteomic assay were filtered to FDA-approved drug targets, resulting in 276 drug-targets of which 33.7% are secreted (Supplementary Fig. 1–2; the overlap of UKB self-reported medications, GP-script medications, and the FDA-flagged Olink subset is shown in Supplementary Fig. 19 and as the inset of Fig. 5a). To downselect proteins for targeted assays, the overall proteomic model was optimized by number of proteins, where accuracy as R²_test and MAE_test ranged from 65% accurate and 4 years in error with 50 proteins to greater than 85% accurate and less than 2.5 years in error with more than 1000 proteins (Fig. 5a). A proteomic clock only trained on FDA-approved protein targets achieved an R²_test and MAE_test of 0.59 and 4.25 years, respectively (Fig. 5b). The proteomic age gap (predicted - actual age) showed that those who took drugs were significantly proteomically younger relative to chronological age than those who did not (Fig. 5c), even though within-person slopes with and without FDA-approved drugs were not significantly different and predicted ages with drugs were significantly higher than those without drugs (older and more sick; Supplementary Fig. 20-21).

**Figure 5.**
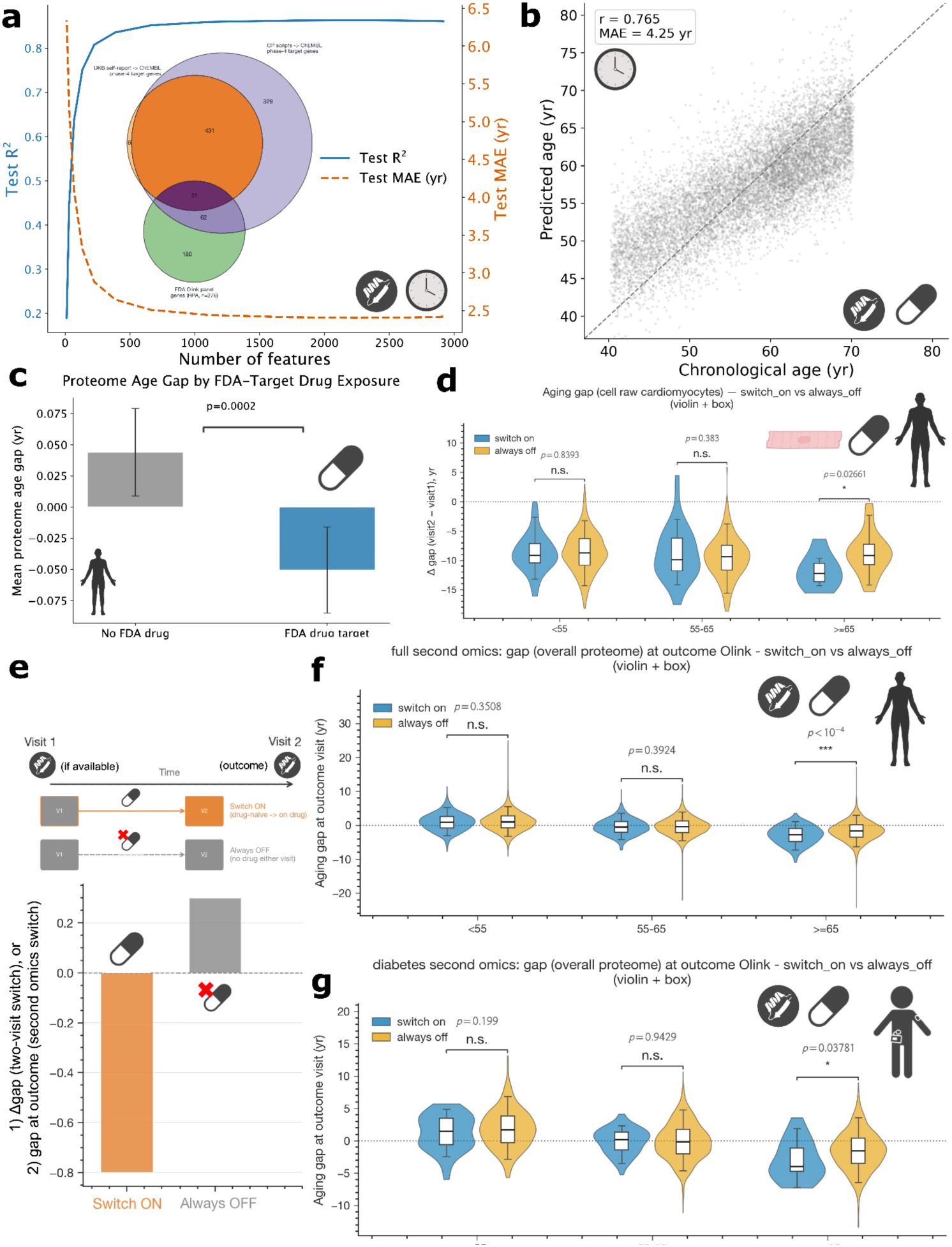
FDA-approved drug-target clock model and longitudinal effects of drug exposure on overall and subsystem-specific clocks across full and diabetic cohorts. (a) Feature optimization of proteomic aging clock models showing test R² and MAE as a function of the number of input proteins (inset: overlap between UKB self-reported medications mapped to ChEMBL phase-4 targets, GP-script-derived ChEMBL phase-4 targets, and FDA-flagged Olink panel proteins from HPA). (b) Proteomic aging clock trained only on FDA-approved drug-target proteins. (c) Mean proteome age gap (predicted age - chronological age) for participants with no FDA drug vs. at least one FDA drug-target exposure (Welch p=0.0002). (d) Two-visit switch comparison of Δgap (visit 2 - visit 1) using the cardiomyocyte raw-cell clock, stratified by age group (<55, 55–65, ≥65), for switch-on vs. always-off participants. (e) Schematic of the two-visit switch and second-omics switch designs, and full-cohort second-omics comparison of overall proteome age gap at the outcome visit between switch-on and always-off participants by age group. (f) Diabetic-cohort second-omics comparison of overall proteome age gap at the outcome visit between switch-on and always-off participants by age group.

Having linked mAge features to FDA-approved drug targets, we evaluated whether new exposure to these drugs was associated with measurable changes in biological age, subsystem-specific aging, mortality risk, and age at death. By comparing participants with proteomics assays at baseline and follow-up visits who started any drug that targets a measured FDA-approved protein to those that never took a drug, a significant reduction in cardiomyocyte-specific age in older cohorts (age ≥ 65) was observed (p = 0.03; Fig. 5d) as measured by Δgap between follow-up and baseline. The proteins measured by the cardiomyocyte-specific clock in these comparisons revealed drug-target pairs like GLP1R with GLP-1 receptor agonists (e.g., semaglutides) and mitochondrial respiratory complex-I forming proteins (NDUFB7 and NDUFS6) with metformin for potential longevity interventions (Tab. S4), even though the age gap reduction using the overall proteomic clock was not significant in any age stratum (Supplementary Fig. 20a, Supplementary Fig. 20b; Supplementary Fig. 21). Importantly, these drug-target associations were identified from the collective set of proteins comprising the cardiomyocyte-specific cloc (Tab. S4)k, rather than from isolated single-protein effects alone.

The schematic outlining the two-visit and second-omics study designs, alongside the primary second-omics results for the full cohort, is presented in Fig. 5e. A significant attenuation of the proteomic age gap at the outcome visit was observed among participants who initiated new drug therapies in the full cohort (p<10^-4^; Fig. 5e), an effect that was further pronounced in the older strata of the diabetic cohort (p=0.04; Fig. 5f). Diabetic participants with drugs also showed lower mortality overall and lower subsystem-specific mortality in several subsystems (e.g. heart, hepatocytes), lower proteomic age, lower proteomic age gap in many organs/tissues (brain, blood, ECM, blood vessel, liver, lung, lymphoid), and in many cell types: adipose (and adipose progenitors), adrenal, grouped fibroblasts (and raw fibroblasts), immune, reproductive (and early spermatids), hepatocytes, macrophages, and neutrophils. Representative diabetic-cohort second-omics analyses for cell-grouped epithelial and raw minor salivary glandular clocks are shown for age gap (Supplementary Fig. 22), predicted age (Supplementary Fig. 23), and mortality readouts (predicted time-to-death in Supplementary Fig. 24 and log partial hazard in Supplementary Fig. 25). For diabetics, these improvements were driven by drugs like gabapentin targeting intravoltage-gated ion channels CACNB1/CACNB3 (intracellular) and CACNA1H (membrane) in brain/fibroblasts, adipose/skeletal muscle, and intestine/overall proteome, respectively, while insulin targets, such as INSR, only showed improvements in mortality via epithelial and salivary gland clocks (Tab. S5).

To test whether mAge-nominated interventions affect lifespan and not only proteomic age, we examined age-at-death in deceased UK Biobank participants and benchmarked the human signal against validated mouse longevity outcomes. By analyzing drugs affecting the deceased cohort and relating them to interventions validated in mice^32,33,48,49^ (Fig. 6a-d; the three-way overlap between NIA ITP interventions, UKB medication names, and FDA-flagged Olink-panel-targeting drugs is shown in Fig. 6e and Supplementary Fig. 26), the increase in Δage-at-death was significant with aspirin (p = 0.007 across all ages and p = 0.002 for ages ≥ 55; Fig. 6a), ramipril (p = 0.003-0.004 for ages ≥ 55; Fig. 6b), metformin (nearly significant: p = 0.07 across all ages and p = 0.057 for cohorts between 55 and 65; Fig. 6c), and simvastatin (p = 0.04 across all ages; Fig. 6d). Using mAge to assess the effect of ACE inhibitors (Extended Data Fig. 9), which were shown to have a large increase in mouse lifespan^33,50^, lisinopril showed a reduction in proteomic age gap in the full proteome (p = 0.02 for ages ≤ 55; Extended Data Fig. 9a), in blood vessels (p = 0.015 for cohorts between 50 and 60; Extended Data Fig. 9b), and in macrophages (p = 0.02 for cohorts between age 50 and 60; Extended Data Fig. 9c). While ramipril did not show significant results in the overall proteome, it showed a significant reduction in lymphoid tissue-specific proteomic age (p = 0.01 for ages <50 and p<10^-4^ for ≥ 60; Extended Data Fig. 10a), in reproductive cells (p = 0.03 in cohorts between age 55 and 65; Extended Data Fig. 10b), and in mesothelial cells (p = 0.04 in cohorts between 50 and 60; Extended Data Fig. 10c). The Pearson correlation between increase in lifespan in mice versus increase in Δage-at-death in humans was found to be 0.55 (Fig. 6f), where aspirin (6-8%) and ACE inhibitors (4-6%) showed similar increases in both mouse and human (Fig. 6g), while metformin and simvastatin only showed an increase in Δage-at-death (4-6%) in humans (Fig. 6g). The mouse-null result is consistent with prior ITP findings for both drugs as monotherapy and likely reflects mechanism specificity: simvastatin and metformin act primarily on cholesterol and glucose pathways whose downstream pathologies, such asatherosclerosis and type 2 diabetes, are leading causes of human mortality but are largely absent in laboratory mice, where cancers account for the majority of natural deaths^51^, whereas aspirin and ACE inhibitors engage anti-inflammatory and renin-angiotensin pathways that influence aging-related tissue damage in mice independent of overt cardiovascular disease.

**Figure 6.**
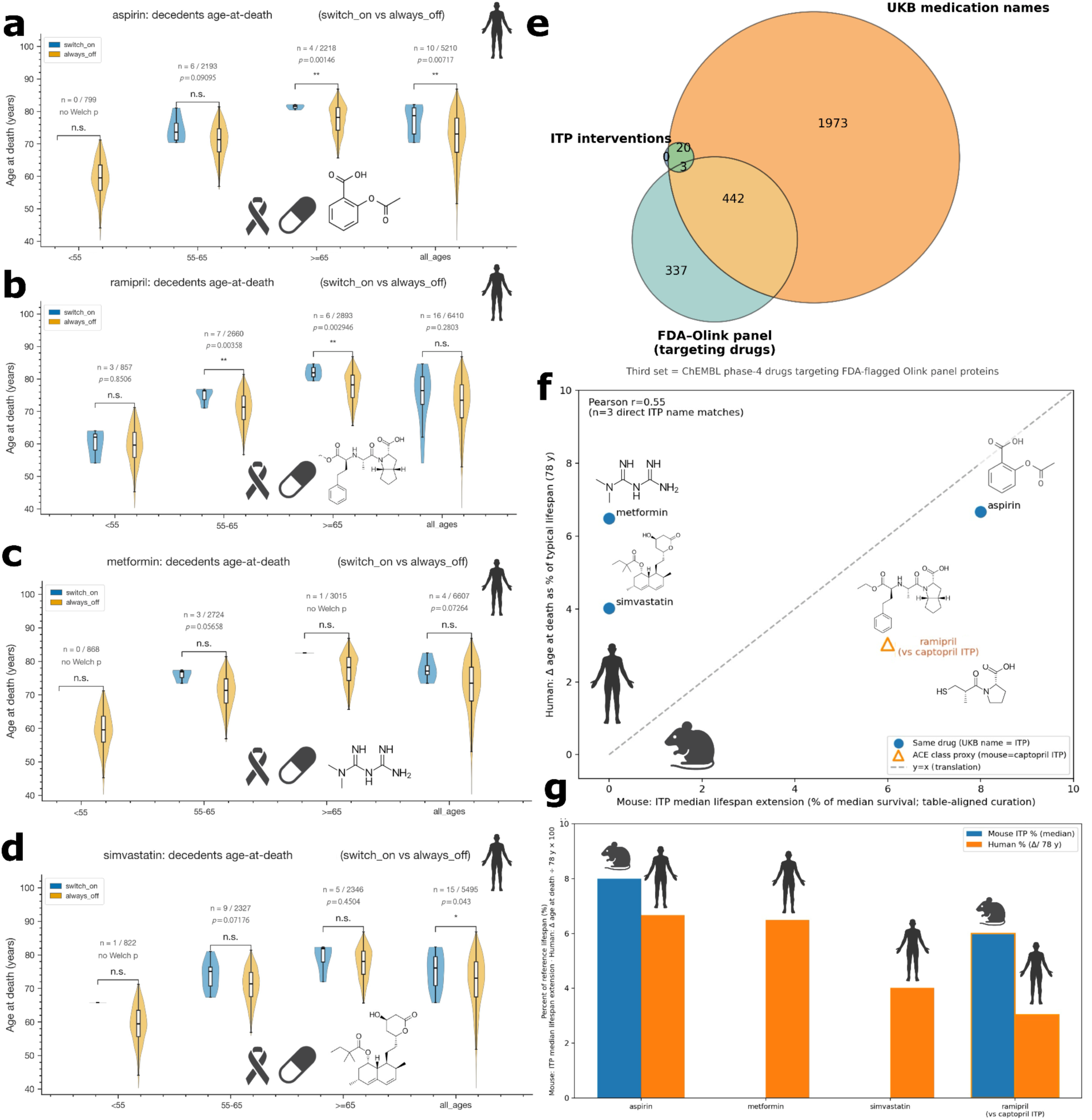
Evaluating the longitudinal effects of overlapping drugs between NIA ITP interventions and HPA FDA-approved proteomic targets, and comparing increases in lifespan from ITP interventions in mice with later age-at-death in UKB humans. (a–d) Age-at-death of deceased UKB participants by switch-on vs. always-off status, stratified by age group, for aspirin (a), ramipril (b), metformin (c), and simvastatin (d). (e) Three-way overlap between NIA ITP interventions, UKB medication names, and FDA-flagged Olink-panel targeting drugs (third set: ChEMBL phase-4 drugs targeting FDA-flagged Olink panel proteins). (f) Correlation between mouse ITP median lifespan extension (% of median survival) and human Δage-at-death (% of typical 78-year lifespan) for FDA-approved drug interventions, with direct ITP name matches in blue and ACE-class proxy (ramipril vs. captopril ITP) in orange (Pearson r=0.55). (g) Side-by-side bar comparison of mouse ITP % lifespan extension and human Δ age-at-death % for aspirin, metformin, simvastatin, and ramipril (vs. captopril ITP).

## Discussion

Our findings establish mAge as a scalable framework for resolving biological aging across multiple levels of organization, from organism-wide age and mortality risk to organ-, tissue-, and cell-associated proteomic signatures. By integrating plasma proteomics with wearable-derived physiology, mortality outcomes, intrinsic capacity, longitudinal measurements, and drug-target mappings, mAge extends biological age modeling beyond prediction toward interpretation and intervention nomination. The central advance is not simply improved clock performance, but the ability to identify which biological subsystems contribute to aging-related risk, monitor their trajectories over time, and connect these signals to existing pharmacological and lifestyle interventions.

### A multimodal model for biological aging

mAge integrates plasma proteomics, wearable accelerometry, and mortality follow-up into a unified biological aging framework. With overall organism-level age prediction already at high accuracy from proteomics alone (Fig. 1c), the largest gains come not from improving age regression but from changing the objective function: mortality models trained to maximize the C-index of risk ranking between living-deceased and deceased-deceased pairs outperform state-of-the-art (SOTA) clocks trained to minimize residual error in time-to-death (Tab. 1)^24^ because time-to-death is sparser by roughly 10× and noisier than mortality risk, which is jointly defined over all participants regardless of survival status. As a result, correlation, R², and MAE in time-to-death all improve compared to SOTA^24^. The multimodal extension that adds wearables further improves correlation, coefficient of determination, and time-to-death MAE (Tab. 1) at a small (<3%) cost in C-index attributable to the smaller proteomics-wearable participant overlap; median imputation across modalities did not recover the lost participants and in fact reduced accuracy, and synthetic-data bridging between modalities and datasets^52^ represents a promising direction for closing this gap. The mAge framework therefore couples two complementary risk predictors, chronological age and mortality hazard, into a single set of features using multimodal inputs that downstream subsystem clocks inherit.

**Table 1.** Comparison of mortality aging clocks.

| Metric | Unimodal (Goeminne et al.<sup>24</sup>) | Unimodal (this study) | Multimodal (this study) |
| --- | --- | --- | --- |
| C_test | — | 0.81 | 0.78 |
| r_test | 0.67 | 0.69 | 0.71 |
| R<sup>2</sup>_test | 0.45 | 0.48 | 0.50 |
| MAE_test (yr) | — | 4.83 | 4.65 |
| MAE_test (log-hazard) | 0.55 | 0.56 | 0.44 |

### Subsystem-resolved aging offers interpretable maps

Beyond global organism-level prediction, mAge maps plasma proteins to the cell types, tissues, organs, and secretome compartments where they originate, transforming clocks into interpretable maps of where aging occurs. By using HPA to combine multiple data sources without removing overlapping proteins (immunohistochemistry, subsystem-specific RNA-seq, GTEx and FANTOM5 expression atlases, NCBI/Uniprot annotations, and SignalP/DeepPeptide secretion calls) to define aging models for subsystems (Tab. S6), we find that brain- and blood vessel-specific age prediction (Tab. 2) outperforms SOTA models that rely on single sources or that exclude overlapping proteins. The full-proteome organism-level models that exceed mAge by ∼1% in accuracy do so by pruning the input feature set (240 proteins in Wang et al.^25^ with LightGBM, 2,315 in Goeminne et al.^24^ with similar ElasticNet), whereas mAge retains all assayed proteins (Fig. 5a–b) so that all possible drug-target pairs can be evaluated for intervention nomination. The resulting subsystem decomposition reveals that age and mortality signals concentrate unevenly across compartments. This biological resolution cannot be achieved with organism-level clocks.

**Table 2.** Comparison of proteomic aging clocks. MSE is noted where RMSE was not available.

| Tissue | Metric | Oh et al.<sup>22</sup><br>(Nature '23) | Vladimirova et al.<sup>47</sup><br>(bioRxiv '24) | Argentieri et al.<sup>21</sup> (Nat. Med. '24) | Oh et al.<sup>23</sup><br>(Nature '25) | Wang et al.<sup>25</sup><br>(Nat. Aging '26) | Goeminne<sup>24</sup><br>(Cell Met. '25) | This study<br>(ours) |
| --- | --- | --- | --- | --- | --- | --- | --- | --- |
| Overall | r | 0.72 | — | 0.94 | 0.91 | 0.94 | ≈0.93 | ≈0.93 |
|  | r<sup>2</sup> | ≈0.52 | — | 0.88 | ≈0.83 | 0.88 | 0.88 | 0.87 |
|  | MAE | — | — | 2.24 | 2.69 | — | 2.10 | 2.41 |
|  | RMSE | — | — | 2.85 | — | MSE 6.72 | — | — |
| Brain | r | 0.4–0.6 | ≈0.70 | — | 0.72 | 0.77 | ≈0.61 | ≈0.77 |
|  | r<sup>2</sup> | 0.20–0.36 | ≈0.50 | — | ≈0.52 | 0.60 | 0.37 | 0.60 |
|  | MAE | — | — | — | — | — | 3.14 | 4.10 |
|  | RMSE | — | — | — | — | MSE 14.92 | — | — |
| Blood Vessel / Artery | r | 0.2–0.4 | — | — | 0.71 | 0.75 | ≈0.73 | ≈0.76 |
|  | r<sup>2</sup> | ≈0.04–0.16 | — | — | ≈0.50 | 0.56 | 0.53 | 0.58 |
|  | MAE | — | — | — | — | — | 3.27 | 4.28 |
|  | RMSE | — | — | — | — | MSE 16.45 | — | — |

### Wearables add dynamic physiological information to molecular data

A central finding of mAge is that wearable accelerometry contributes information that is not redundant with plasma proteomics. Adding wearables roughly halves the sum of model weights assigned to proteins overall, drops 260 less-predictive proteins from the multimodal mortality model, and reweights the remaining proteins toward markers of cardiac load (NTproBNP, NPPB) and metabolic health (IGFBP3) while preserving or strengthening the canonical mortality drivers GDF15 and EDA2R (Fig. 1e). The wearable features that take their place, e.g. no-wear-time-adjusted average acceleration, total daily acceleration, and acceleration during the late-afternoon window (4–6 pm), enter the model as protective signals at hazard ratios comparable to leading proteomic markers. Subsystem-resolved clocks reveal where wearables matter most: cardiac and immune mortality models gain up to 44% in MAE log-hazard, and intracellular proteins (often released upon cell death and necrosis) gain 28%, while structural-protein-based clocks (blood vessel, proximal tubular cell) gain little (Fig. 3a). This pattern is biologically coherent: wearables capture dynamic physiological state, such as cardiopulmonary capacity, sleep regularity, daily activity load, which complements the more chronic structural and damage signals encoded in plasma proteins. Together, proteomics and wearables form a longitudinal aging readout that neither modality achieves alone, and they make a stronger case for multimodal aging biomarkers than either modality could in isolation.

### From biomarkers to interventions

mAge bridges biomarker discovery and intervention nomination through three complementary axes. First, longitudinal tracking of within-person mAge trajectories (Fig. 5d–f, Extended Data Fig. 9–10) deconvolves age-related drug effects from population trends that dominate cross-sectional cohorts of older, sicker, and more medicated individuals. Second-visit omics readouts in well-defined cohorts, most notably the diabetes cohort with second-visit assays, are sufficient to surface drug effects too subtle to detect in the full cohort (Fig. 5f, Supplementary Fig. 22–25). Second, mAge can connect each clock feature to FDA-approved drug-target pairs, recovering known aging-relevant interventions (cardiomyocyte-specific reduction with GLP-1 agonists and metformin via NDUFB7/NDUFS6; gabapentin reducing brain/fibroblast/intestine aging via voltage-gated channels CACNB1/3 and CACNA1H in diabetics) while nominating mechanistically interpretable new candidates. Third, mortality-based validation in deceased UKB participants converges with the NIA Interventions Testing Program: aspirin, ramipril, metformin, and simvastatin all show significantly later age-at-death (Fig. 6a–d), with a Pearson correlation of 0.55 between mouse ITP percent lifespan extension and human percent age-at-death increase (Fig. 6f–g).

Beyond pharmacology, multimodal mAge supports lifestyle-based intervention discovery: daytime activity load and 4–6 pm acceleration predict reduced mortality (Fig. 1e), and current-smoker baseline age gaps exceed those of non-smokers (Supplementary Fig. 27–28), enabling generalization across health states^53^. The full hazard-ratio spectrum of all wearable features retained by the multimodal Cox mortality model is summarized in Supplementary Fig. 29: 15 of the 17 retained wearable features are protective, led by no-wear-bias-adjusted average acceleration (HR = 0.959), afternoon acceleration 17:00–18:00 (HR = 0.964), and overall daily average acceleration (HR = 0.964), while only midnight (00:00–01:00, HR = 1.006) and early-morning (04:00–05:00, HR = 1.001) acceleration cross to the risk side, both far below the protein-level top-25 risk-feature threshold (HR ≥ 1.024). This wearable-side decomposition shows that the protective contribution of accelerometry is not concentrated in any single time window but is broadly distributed across daytime moderate-intensity activity, with the only nominal risk signals tracking nocturnal/predawn activity that is more parsimoniously interpreted as sleep disruption than as independent biological risk. Pairing this clock-side evidence with virtual-cell perturbation models that predict transcriptomic and proteomic responses to gene knockdown, overexpression, or knockout would extend the same framework from drug-level to gene-level intervention nomination.

### Limitations

This study has several limitations. First, within-person mAge slopes currently quantify model quality in addition to the rate of biological aging itself, since clocks are trained at baseline and then applied longitudinally. Predictions can drift off-diagonal at the input-distribution extremes through regression to the mean. Second, some overlap in the terms used is unavoidable; for example the terms age gap, age acceleration, and age deviation are used interchangeably in the field to denote the difference between predicted and chronological age, consistent with prior usage. Third, statistical comparisons between switch-on and always-off arms use Welch’s t-test (which adjusts degrees of freedom for unequal variances and unequal n) rather than Student’s, which is appropriate for these unbalanced cohorts but does not correct for confounding by polypharmacy: drug exposures are encoded as binary flags, and the prevalence of ≥5 medications in UKB participants aged 40–69 is approximately 18.3%^54^. Fourth, no ground-truth proteome exists; cross-platform integration of multiple proteomic technologies, with overlap-based confidence calls, will be required to make mAge platform-agnostic. Mass spectrometry could additionally probe post-translational modifications, and coupled transcriptomics could reveal post-transcriptional decoupling between gene and protein expression with age. Fifth, our study employs UK Biobank data and could be limited to populations profiled^55^.

### Conclusions and future directions

mAge demonstrates that a multimodal, subsystem-resolved framework can simultaneously diagnose biological aging globally and across organs, tissues, and cell types, while also nominating both pharmacological and lifestyle interventions whose effects can be quantified longitudinally. By pinpointing the origin of plasma proteins, subsystem-specific mAge can longitudinally track health states across compartments, and through mapping protein targets to FDA-approved drugs, it nominates longevity interventions with overall and subsystem-specific impacts. The architecture lends itself to several natural extensions. By integrating plasma proteomics with wearable physiology and clinical measurements such as grip strength, mAge can be experimentally verified in real time^56^ on individuals unseen during training. Future work will examine whether a wearables-only, proteomic-imputed readout retains predictive value when proteomic assays are unavailable, enabling continuous monitoring without periodic blood draws. Additional modalities are expected to further improve prediction power of the models. As more data are collected and the multimodal overlap between proteomics, wearables, mortality, and clinical endpoints grows, we expect mAge to scale in predictive power: the small (<3%) C-index cost from the current proteomics–wearable overlap should diminish, and subsystem-specific clocks with fewer features per compartment should benefit most from larger training sets. Together with prospective and randomized cohorts to confirm or refute the drug-exposure associations reported here, this scaling trajectory positions mAge as a foundation for personalized longevity intervention nomination rather than as a passive readout.

## Methods

### Data consolidation

Patient multimodal data was consolidated from the UKB^57^ (n = 503,317) using the UKB Research Analysis Platform (RAP)^58^, with plasma proteomics^23^ for 53,039 participants (Olink, 2,922 proteins), wrist accelerometry^27–28^ for 103,572 (Axivity AX3, 7-day), 56,961 recorded deaths (ICD-10; 2,369 unique codes), and 450,740 with ≥1 diagnosis (12,353 ICD-10 codes; 18.4M records). Protein–gene mapping and source organs/tissue, and grouped/raw cell annotations were extracted from the Human Protein Atlas (HPA)^36–38,59^. Drug-target annotations were extracted from the overlap between HPA drug targets and ChEMBL^39^ drug-target pairs.

### Intrinsic capacity modeling

IC was calculated from 137 UKB fields across five World Health Organization-defined domains^8–16^(Table S2 compares this continuous functional-reserve operationalization with prior UK Biobank IC implementations that use deficit-accumulation scoring): locomotion (20 fields), cognition (24 fields), psychological (33 fields), vitality (40 fields), and sensory (35 fields), where each item was z-scored with signs flipped, such that higher indicates better health outcomes. The total IC score is the z-score mean of ≥2 valid domains. Elastic net models were trained in a sequential two-stage pipeline: predicted outputs from upstream models serve as input features to downstream models. In stage 1, baseline IC is predicted from proteomics, accelerometry, or their combination; in stage 2, predicted baseline IC is combined with clinical features (BMI, grip strength, gait pace, demographics) to predict future-visit IC and prevalent/incident comorbidities. All stages use an 80/20 train–test split with 5-fold cross-validation (CV) on the training set for hyperparameter selection. Functional decline, as quantified by decreasing IC with age, are shown in Supplementary Fig. 30.

### Continuous outcome AI models

For IC prediction, elastic net linear regression minimizes ‖y − Xβ‖² + λ₂‖β‖² + λ₁‖β‖₁, where X are input features, y the outcome, and λ₁, λ₂ control regularization strength. Hyperparameters were selected by CV to minimize MAE, MAE = (1/n) Σᵢ |yᵢ,true − ŷᵢ,predicted|. The same model was used for biological age prediction from proteomic and/or wearable features, with grid search over α ∈ {10⁻⁵, …, 10⁵} and ℓ₁-ratio ∈ {0.0, 0.1, …, 1.0}. Imputation and standardization were fit on training folds only to prevent data leakage. Biological aging deviation gap was defined as predicted age − chronological age.

### Binary outcome AI models

For comorbidity prediction, prevalent disease (any hospital record prior to baseline) and incident disease (first diagnosis on or after baseline, among those without prevalent disease), elastic net logistic regression minimizes binary cross-entropy loss, −Σᵢ [yᵢ log pᵢ + (1−yᵢ) log(1−pᵢ)] + λ₂‖β‖² + λ₁‖β‖₁, where yᵢ ∈ {0,1} is the binary disease label, pᵢ = (1 + e⁻ˣᵢ·β)⁻¹ is the predicted disease probability, Xᵢ the feature vector, β the coefficient vector, and λ₁, λ₂ control regularization strength. Hyperparameters were selected by CV to maximize the area under the receiver operating characteristic (AUC-ROC), where the ROC curve plots true positive rate (sensitivity, TP/(TP+FN)) against false positive rate (1 − specificity, FP/(FP+TN)) across all classification thresholds, and AUC discriminates the probability that a randomly drawn positive case is ranked above a randomly drawn negative case.

### Survival outcome AI models

Mortality risk was modeled from proteomic and multimodal features using ElasticNet-penalized Cox proportional hazards^60^, λ(t | Xᵢ) = λ₀(t) exp(Xᵢ·β), where λ(t | Xᵢ) is the instantaneous event hazard at time t for individual i, λ₀(t) the baseline hazard, Xᵢ the covariate vector, and β the coefficient vector. Parameters were estimated by penalized partial likelihood arg minᵥ [−ℓ·ₐᵣ·ᵢₐ·(β) + λ₂‖β‖² + λ₁‖β‖₁], where ℓ·ₐᵣ·ᵢₐ·(β) is constructed from observed event times and risk sets. A 3-fold CV grid search was performed over an auto-generated α path (up to 50 values) and ℓ₁-ratio ∈ {0.1, 0.5, 1.0}, selecting hyperparameters to maximize Harrell’s C-index, defined as the probability that a participant who dies earlier has a higher predicted risk than one who survives longer.

### Definition of mortality model readouts

Throughout the manuscript the Cox linear predictor η_i = X_i·β is referred to as the *predicted log partial hazard*; we report Pearson r, R², and MAE of η_i against chronological age (“MAE log-hazard”) and against time-to-death in years (“MAE time-to-death”). The *predicted mortality age* (used in Tab. 1 and Fig. 1d) is obtained by ordinary-least-squares calibration of η_i to chronological age on the training set, â_i = μ_a + (σ_a/σ_η) (η_i − μ_η), where μ and σ denote training-set mean and standard deviation; â_i is therefore a years-scaled rank of mortality risk rather than a calibrated estimate of calendar age. Reported R² values for the mortality model are computed against chronological age unless otherwise stated; we additionally report C-index for held-out risk ranking and MAE in years for time-to-death calibration so that the differing strengths and weaknesses of these readouts can be compared transparently. The same calibration procedure is applied to all subsystem-specific mortality clocks. Final performance for all models was evaluated on the held-out 20% test set. For interpretability, the top 30 features by coefficient magnitude were visualized as bar/lollipop hazard ratio plots, where each hazard ratio HR_j = exp(β_j) represents the multiplicative change in instantaneous mortality risk per unit increase in feature j, displayed on a log scale with a neutral risk reference line at HR = 1. The linear predictor X·β was also used as a continuous mortality score to compute Pearson r, R^2^, and MAE against chronological age. Final performance for all models was evaluated on the held-out 20% test set. For interpretability, the top 30 features by coefficient magnitude were visualized as lollipop hazard ratio plots, where each hazard ratio HR□ = exp β□ represents the multiplicative change in instantaneous mortality risk per unit increase in feature j, displayed on a log scale with a neutral risk reference line at HR = 1.

### Multimodal combinations

Models spanned a unimodal-to-multimodal hierarchy across five modalities: proteomics (2,922 Olink NPX proteins), accelerometry (96 wrist-accelerometer features), clinical biomarkers and lifestyle (55 features), multimorbidity (37 diseases and comorbidity index features), and demographics (sex). Each model was evaluated independently (unimodal), in all (5 choose 2) = 10 pairwise (bimodal), (5 choose 3) = 10 trimodal combinations, (5 choose 4) = 5 quadmodal combinations, and jointly as a multimodal (pentamodal) model with 3,111 features.

### Organ, tissue, and cell-type mapping

Plasma proteins were mapped to biological origin at three levels using Human Protein Atlas (HPA)^36–38,59^ annotations: (i) secretome location (e.g., secreted to blood, secreted in brain, intracellular) from protein class fields; (ii) top-expressing tissue/organ, assigned as the subsystem with maximum normalized transcripts per million (nTPM = TPM□ / (Σ· TPM·) × 10⁶, where j indexes tissue and k all tissues); and (iii) primary cell type from HPA single-cell enrichment annotations. Proteins overlapping multiple subsystems were retained in each (non-exclusive assignment) prior to independent per-subsystem clock training.

### Drug-target mapping and intervention testing

Proteins were cross-referenced with FDA-approved drug targets via HPA^36–37,59^ and ChEMBL^39^. Two complementary switch-on designs assessed intervention effects. The *two-visit* design estimates within-person change Δ = x_{t2} − x_{t1} between baseline and follow-up, where treatment is defined by drug initiation between visits. The *second-omics* design evaluates the effect of treatment using the proteomics assay once treatment is flagged on for the first time after no prior treatment. In both designs, treated vs untreated groups were compared using Welch’s t-test, t = (x̄₁ − x̄₂) / √(s₁²/n₁ + s₂²/n₂), where x̄·, s·², and n· denote the sample mean, variance, and sample size of group k ∈ {treated, untreated}. Degrees of freedom were estimated as ν = (s₁²/n₁ + s₂²/n₂)² / [(s₁²/n₁)²/(n₁−1) + (s₂²/n₂)²/(n₂−1)].

## Data Availability

The UK Biobank data are available under controlled access. This project is approved under application number 21988. Researchers requiring access should register, complete trainings, and apply via the following steps (https://www.ukbiobank.ac.uk/enable-your-research/apply-for-access) and pay a fee depending on the tier of data requested.

## Author Contributions

S.A.A., O.O.A., J.S.G., and V.N.G. conceptualized and designed the study. S.A.A. performed the work. J.R.W., O.O.A., J.S.G., and V.N.G supervised the work and provided technical input to the data curation and analysis. M.F., T.Z, D.F. and M.P.S. contributed to the conceptual interpretation of the results, provided scientific input and discussion. All authors reviewed the final version.

## Acknowledgments and Disclosures

This work was supported by grants from the NIA and the Hevolution Foundation to V.N.G. S.A.A. was funded by the MIT Department of Civil and Environmental Engineering (CEE) under the Friesecke (1961) Fellowship Fund. J.S.G. and O.O.A. are supported by NIH grants R01-EB031957, R01-AG074932, and R01-GM148745; G. Harold & Leila Y. Mathers Charitable Foundation; Rett Syndrome Research Trust; The Gordon and Betty Moore Foundation; Impetus Grants; Cystic Fibrosis Foundation Pioneer Grant; Google Deepmind; Sanofi; Yosemite; Michelson Foundation; Hevolution Foundation; American Federation for Aging Research; Pivotal Life Sciences; and the MGB Gene and Cell Therapy Institute.

## Code Availability

All custom code used for the analyses will be made available on GitHub upon publication.

## Competing Interest Statement

J.S.G. and O.O.A. are co-founders of Terrain Biosciences, Monet Therapeutics, and Transit Therapeutics. M.P.S. is a cofounder and scientific advisor of Crosshair Therapeutics, Exposomics, Filtricine, Fodsel, Iollo, InVu Health, January AI, Marble Therapeutics, Mirvie, Next Thought AI, Orange Street Ventures, Personalis, Protos Biologics, Qbio, RTHM, and SensOmics. M.P.S. is a scientific advisor of Abbratech, Applied Cognition, Enovone, Jupiter Therapeutics, M3 Helium, Mitrix, Neuvivo, Onza, Sigil Biosciences, TranscribeGlass, WndrHLTH, and Yuvan Research. M.P.S. is a co-founder of NiMo Therapeutics. M.P.S. is an investor and scientific advisor of R42 and Swaza. M.P.S. is an investor in Repair Biotechnologies. D.F. is a co-founder of Edifice Health and Cosmica Inc. The remaining authors declare no competing interests.

## Main Text Figures and Tables

*Icon glossary (used across all figures).* A consistent set of small inline icons identifies the data modality and the model output across panels: **protein/molecule** icon (a stylized double-helix/molecule glyph) marks proteomic inputs; **watch/wrist** icon marks wearable accelerometry inputs; **plasma vial** icon marks plasma-based proteomic assays; **ribbon** icon marks mortality/survival outcomes; **clock** icon marks chronological-age regression outputs; **tissue/anatomy** icon marks organ- or tissue-resolved subsystem clocks; **cell** icon marks cell-type-resolved subsystem clocks; **pill/capsule** icon marks drug-exposure analyses. The same glyph next to the protein icon in panels (e.g., Fig. 2g, Fig. 3b/3d) denotes the source modality of features used by that panel’s model. Where a panel uses two modalities, both icons appear adjacent to the panel title. Color coding in mortality plots is applied uniformly: red = increased mortality risk (proteomic) / orange = increased mortality risk (wearable); blue = protective (proteomic) / green = protective (wearable).

## Notes

### Author Declarations

This study used controlled-access data from the UK Biobank under approved application number 21988. UK Biobank has ethical approval from the North West Multi-centre Research Ethics Committee, and all participants provided informed consent.

