## Supplementary Information for "A Multimodal Framework for Organ- and Cell-Resolved Biological Aging and Longevity Intervention Discovery"

### Extended Data Figures

#### *Extended Data Figure Captions*

**Extended Data Fig. 1.** Performance of the unimodal proteomic aging and mortality clocks and representative protein–age associations. (a) Predicted biological age versus chronological age for the unimodal proteomic clock in train (blue) and test (orange) sets, with $R^2$ and MAE reported for each split and an OLS fit (test) ±95% CI. (b) Predicted log partial hazard versus chronological age for the unimodal proteomic mortality model in train and test sets, with C-index, correlation, $R^2$, MAE(age), and MAE(lh) reported. (c–f) Hexbin density plots of protein expression (NPX) versus age for representative age-associated proteins: GDF15 (c) and EDA2R (d) increase with age (positive Pearson r, blue), while IGDCC4 (e) and CTSV (f) decrease with age (negative Pearson r, orange).
**Extended Data Fig. 2.** Relationship between proteomic mortality risk, chronological age, and time-to-death. (a) Mortality risk (SD from population mean) within age bands for survivors (blue) vs. participants deceased during follow-up (orange), with Cohen's d effect sizes annotated per band. (b) Cohen's d effect sizes (future decedents - survivors) by age at assessment, with reference lines at small (d=0.2) and medium (d=0.5) effect. (c) Mortality risk for decedents stratified by proximity to death (0–2, 2–5, 5–10, 10–18 yr; ^2=0.0003). (d) Mortality risk for decedents stratified by chronological age at recruitment (^2=0.0282). (e) Within-group standard deviation of mortality risk comparing time-to-death bins (blue) and age bins (orange). (f) Time-to-death distribution within age bands among deceased participants (n=56,961), with mean ±95% CI overlaid. (g) Within-age standard deviation of time-to-death across age bands. (h) Annual change in IC score (health decline) by proximity to death among deceased participants (^2=0.0003). (i) Annual change in IC score by chronological age at recruitment for deceased (orange) vs. survivors (green; ^2_deceased=0.0053).
**Extended Data Fig. 3.** Performance of tissue- and cell-specific proteomic aging and mortality clocks. (a) Predicted biological age versus chronological age for the tissue-specific clock (train vs. test). (b) Predicted log partial hazard versus chronological age for the brain-tissue mortality clock (80/20 split). (c) Predicted biological age versus chronological age for the raw cell-specific clock (train vs. test). (d) Predicted log partial hazard versus chronological age for the grouped adipose-cell mortality clock (80/20 split). (e) Test $R^2$ across all aging clock variants (cell raw in orange, cell grouped in yellow, full proteome in pink, secretome in blue, tissue in green), with MAE overlaid as a yellow line. (f) Concordance index (C-index) for time-to-death across all subsystem mortality clocks, grouped by secretome (blue), tissue (green), cell grouped (orange), and cell raw (red); dashed line at C=0.5 indicates chance performance.
**Extended Data Fig. 4.** Age-correlation patterns of secreted versus intracellular plasma proteins. (a) Density distribution of Pearson correlations between protein level (NPX) and age for secreted (n=817, blue) and not-secreted/intracellular (n=2,105, gray) proteins; 80% of secreted vs. 61% of intracellular proteins show r>0. (b) Percentage of proteins with positive age correlation, broken out by secretome class (blood, ECM, secreted other, brain, reproductive male/female, not secreted). (c) Mean Pearson r (protein vs age) by secretome class. (d) Full distribution of age correlations for secreted vs. not-secreted proteins (violin + box; p<0.001). (e) Subcellular breakdown of the not-secreted compartment (cytoplasm/nucleus, membrane/organelle, other/unknown) shown as age-r KDEs. (f) Direct comparison of age correlations between secreted (n=817) and cytoplasmic/nuclear (n=936) proteins (p<0.001). Summary statistics for all groups are tabulated below the panels.
**Extended Data Fig. 5.** Top age-associated proteins among secreted and intracellular plasma proteins. (a) Empirical cumulative distribution function (ECDF) of age correlations for secreted (blue, n=817) and not-secreted (gray, n=2,105) proteins; 20% of secreted vs. 39% of intracellular proteins are negatively correlated with age. (b–c) Top 15 proteins with most positive age correlation among secreted (b) and

not-secreted (c) compartments. (d–e) Top 15 proteins with most negative age correlation among secreted (d) and not-secreted (e) compartments.

**Extended Data Fig. 6.** Longitudinal trajectories of tissue (organ-context) proteomic aging clocks. (a–s) Predicted biological age versus chronological age across repeated visits for the same random subset of n=40 participants with ≥2 proteomic visits per panel, shown for adipose tissue (a), blood vessel (b), bone marrow (c), brain (d), choroid plexus (e), esophagus (f), heart muscle (g), intestine (h), kidney (i), liver (j), lung (k), lymphoid tissue (l), pancreas (m), parathyroid gland (n), placenta (o), retina (p), skeletal muscle (q), skin (r), and testis (s). Thick blue lines are mean predicted ages; thin gray lines connect within-participant longitudinal visits; dotted diagonals indicate y=x (perfect tracking).

**Extended Data Fig. 7.** Longitudinal trajectories of raw cell-type-specific proteomic aging clocks. (a–s) Predicted biological age versus chronological age across repeated visits for the same random subset of n=40 participants with ≥2 proteomic visits per panel, shown for adipocytes (subcutaneous) (a), adipose progenitor cells (b), adrenal cortex cells (c), adrenal medulla cells (d), breast glandular cells (e), cardiomyocytes (f), colon enterocytes (g), early spermatids (h), endothelial cells (i), enteric glia cells (j), fibroblasts (k), hepatocytes (l), macrophages (m), mesothelial cells (n), minor salivary glandular cells (o), neutrophils (p), proximal tubular cells (q), smooth muscle cells (r), and T cells (s). Thick blue lines are mean predicted ages; thin gray lines connect within-participant longitudinal visits; dotted diagonals indicate y=x (perfect tracking).

**Extended Data Fig. 8.** Distributions of predicted biological age versus chronological age across tissue clocks by visit. (a–u) Density distributions of predicted biological age (solid lines) and chronological age (dashed lines), color-matched by visit (v0, v2, v3), shown for adipose tissue (a), blood vessel (b), bone marrow (c), brain (d), choroid plexus (e), esophagus (f), heart muscle (g), intestine (h), kidney (i), liver (j), lung (k), lymphoid tissue (l), pancreas (m), parathyroid gland (n), placenta (o), retina (p), skeletal muscle (q), skin (r), testis (s), the overall proteome reference clock (t), and the overall proteome plus wearables clock (u).

**Extended Data Fig. 9.** Longitudinal effects of lisinopril exposure on overall and subsystem-specific proteomic age gaps. (a) Δage gap (visit 2 - visit 1) for the overall proteome clock comparing switch-on (blue) vs. always-off (orange) participants, stratified by age (<55, ≥55). (b) Δ age gap for the blood vessel tissue clock, stratified by age (<50, 50–60, 60+). (c) Δ age gap for the raw macrophage cell clock, stratified by age (<55, ≥55). Violins with embedded box plots; Welch p-values and significance annotations shown above each comparison.

**Extended Data Fig. 10.** Longitudinal effects of ramipril exposure on subsystem-specific proteomic age gaps. (a) Δage gap (visit 2 - visit 1) for the lymphoid tissue clock comparing switch-on (blue) vs. always-off (orange) participants, stratified by age (<50, 50–60, 60+). (b) Δage gap for the grouped reproductive-cell clock, stratified by age (<55, 55–65, ≥65). (c) Δ age gap for the raw mesothelial-cell clock, stratified by age (<50, 50–60, 60+). Violins with embedded box plots; Welch p-values and significance annotations shown above each comparison.

*Extended Data Figures*

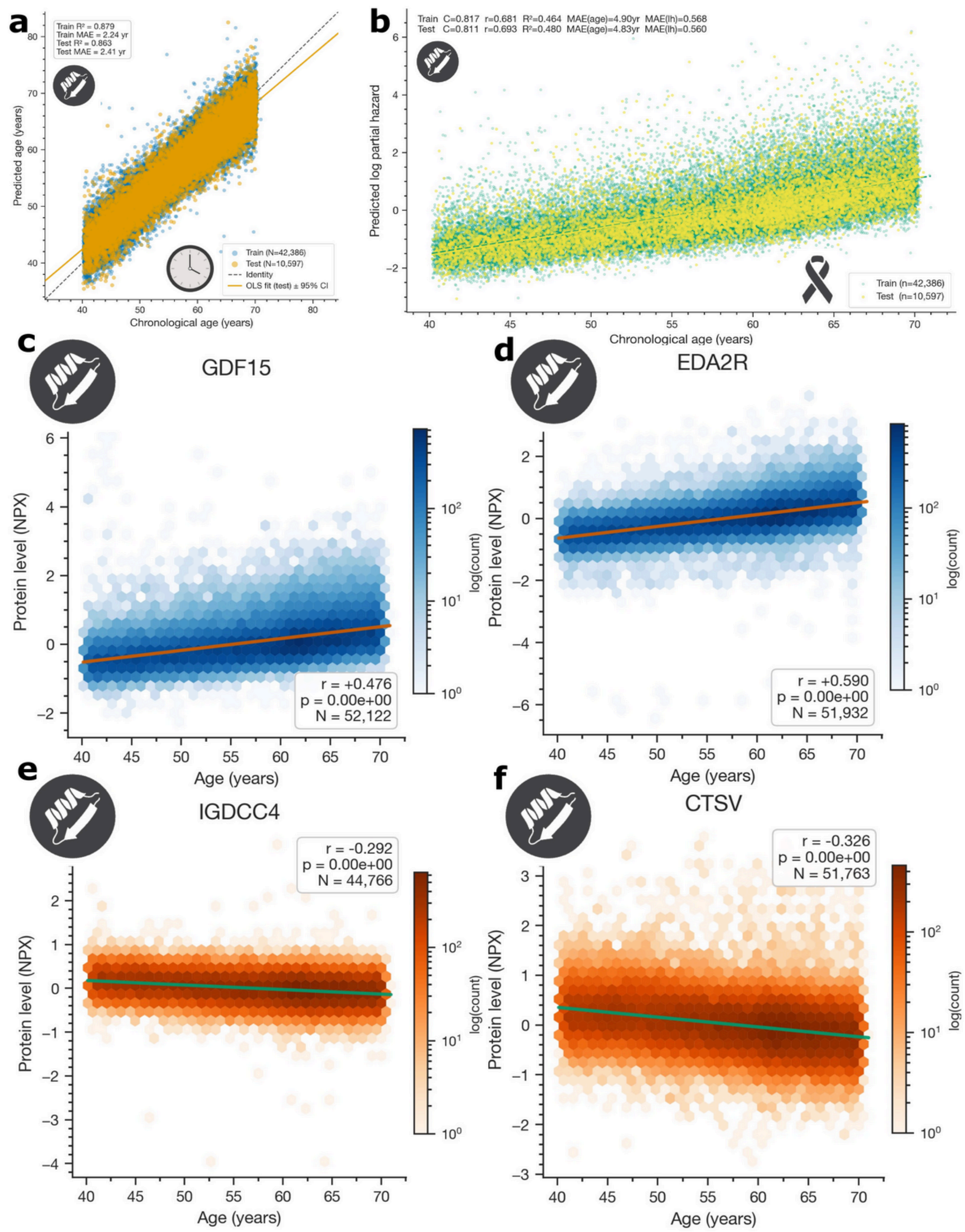


**Extended Data Fig. 1.** Performance of the unimodal proteomic aging and mortality clocks and representative protein–age associations. (a) Predicted biological age versus chronological age for the unimodal proteomic clock in train (blue) and test (orange) sets, with $R^2$ and MAE reported for each split and an OLS fit (test) ±95% CI. (b) Predicted log partial hazard versus chronological age for the unimodal proteomic mortality model in train and test sets, with C-index, correlation, $R^2$, MAE(age), and MAE(lh) reported. (c–f) Hexbin density plots of protein expression (NPX) versus age for representative age-associated proteins: GDF15 (c) and EDA2R (d) increase with age (positive Pearson r, blue), while IGDCC4 (e) and CTSV (f) decrease with age (negative Pearson r, orange).

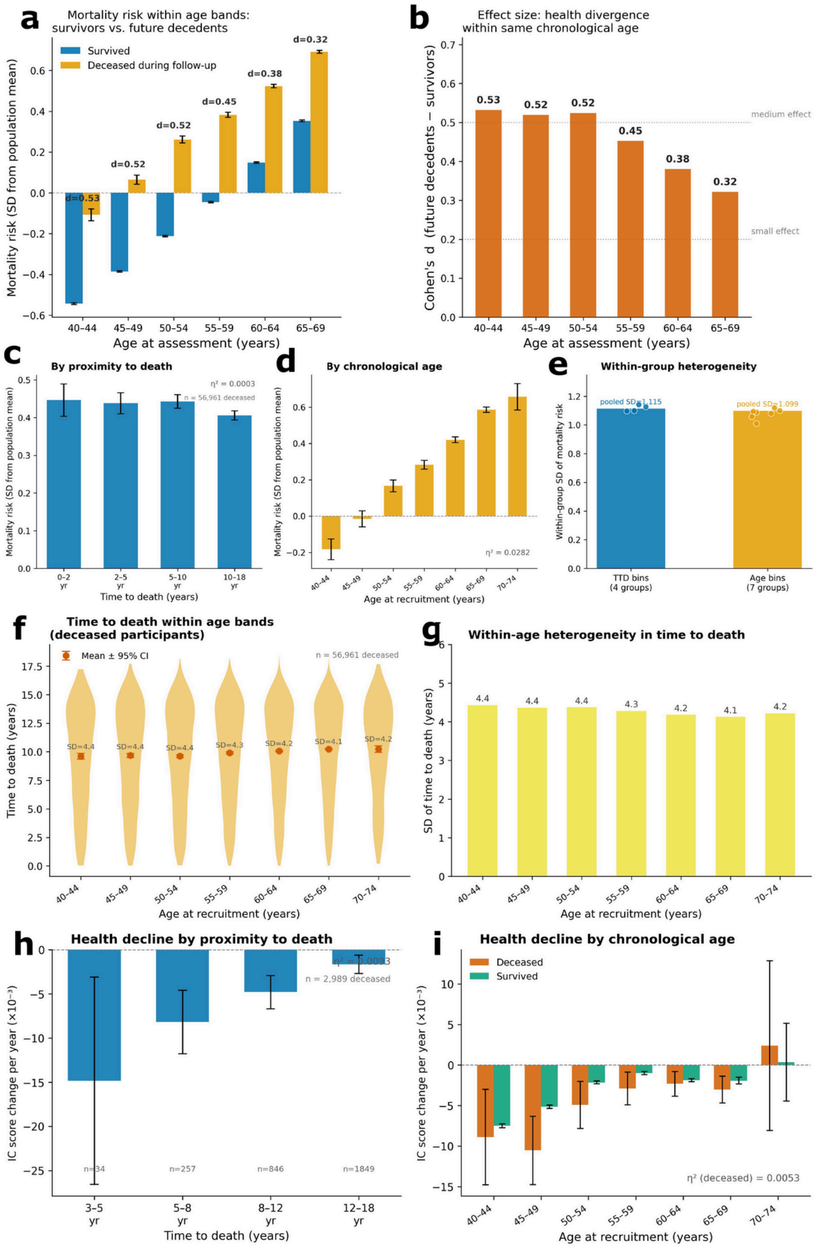


**Extended Data Fig. 2.** Relationship between proteomic mortality risk, chronological age, and time-to-death. (a) Mortality risk (SD from population mean) within age bands for survivors (blue) vs. participants deceased during follow-up (orange), with Cohen's d effect sizes annotated per band. (b) Cohen's d effect sizes (future decedents - survivors) by age at assessment, with reference lines at small (d=0.2) and medium (d=0.5) effect. (c) Mortality risk for decedents stratified by proximity to death (0–2, 2–5, 5–10, 10–18 yr; $\eta^2$=0.0003). (d) Mortality risk for decedents stratified by chronological age at recruitment ($\eta^2$=0.0282). (e) Within-group standard deviation of mortality risk comparing time-to-death bins (blue) and age bins (orange). (f) Time-to-death distribution within age bands among deceased participants (n=56,961), with mean ±95% CI overlaid. (g) Within-age standard deviation of time-to-death across age bands. (h) Annual change in IC score (health decline) by proximity to death among deceased participants ($\eta^2$=0.0003). (i) Annual change in IC score by chronological age at recruitment for deceased (orange) vs. survivors (green; $\eta^2_{deceased}$ =0.0053).

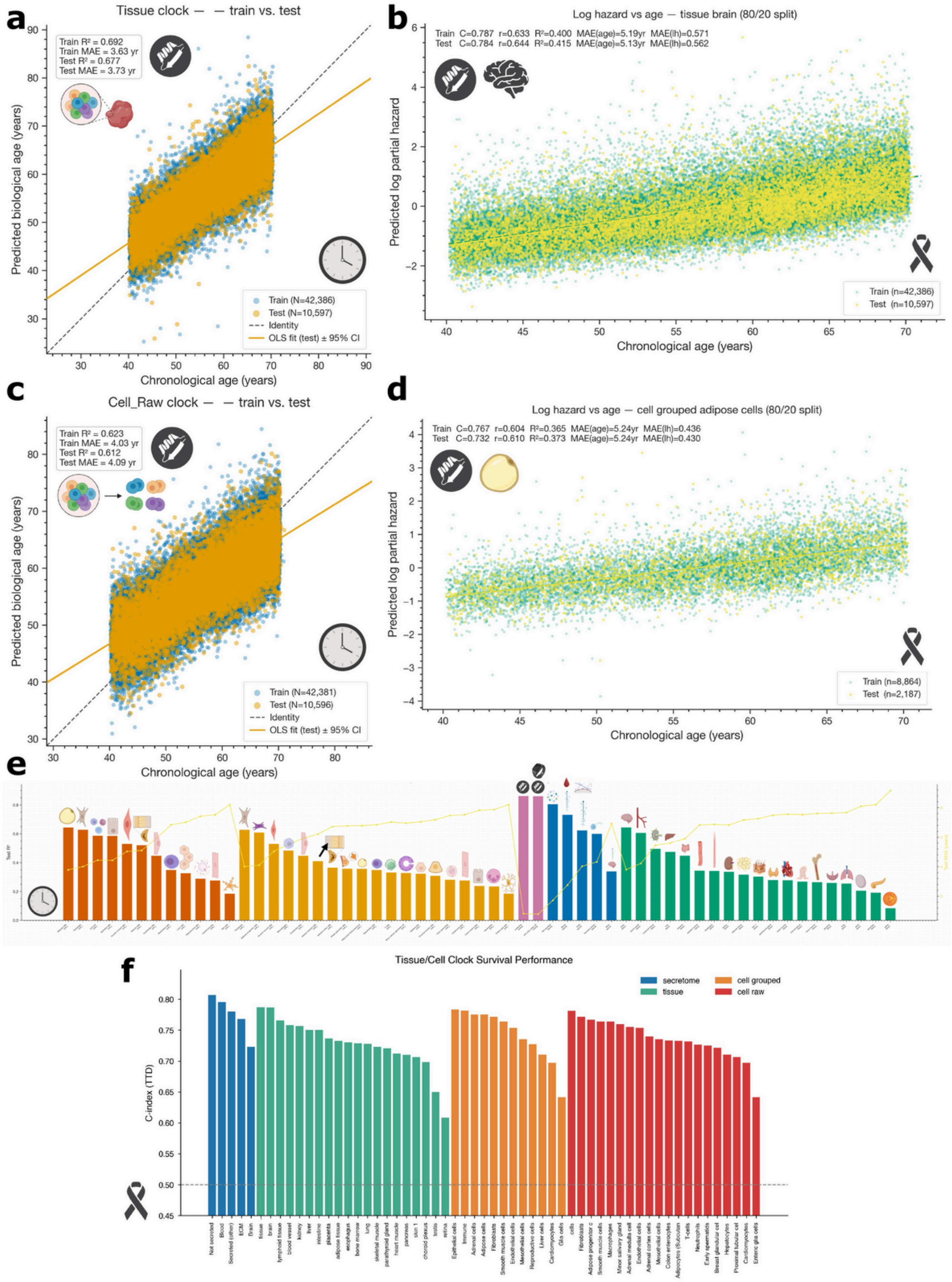


**Extended Data Fig. 3.** Performance of tissue- and cell-specific proteomic aging and mortality clocks. (a) Predicted biological age versus chronological age for the tissue-specific clock (train vs. test). (b) Predicted log partial hazard versus chronological age for the brain-tissue mortality clock (80/20 split). (c) Predicted biological age versus chronological age for the raw cell-specific clock (train vs. test). (d) Predicted log partial hazard versus chronological age for the grouped adipose-cell mortality clock (80/20 split). (e) Test $R^2$ across all aging clock variants (cell raw in orange, cell grouped in yellow, full proteome in pink, secretome in blue, tissue in green), with MAE overlaid as a yellow line. (f) Concordance index (C-index) for time-to-death across all subsystem mortality clocks, grouped by secretome (blue), tissue (green), cell grouped (orange), and cell raw (red); dashed line at C=0.5 indicates chance performance.

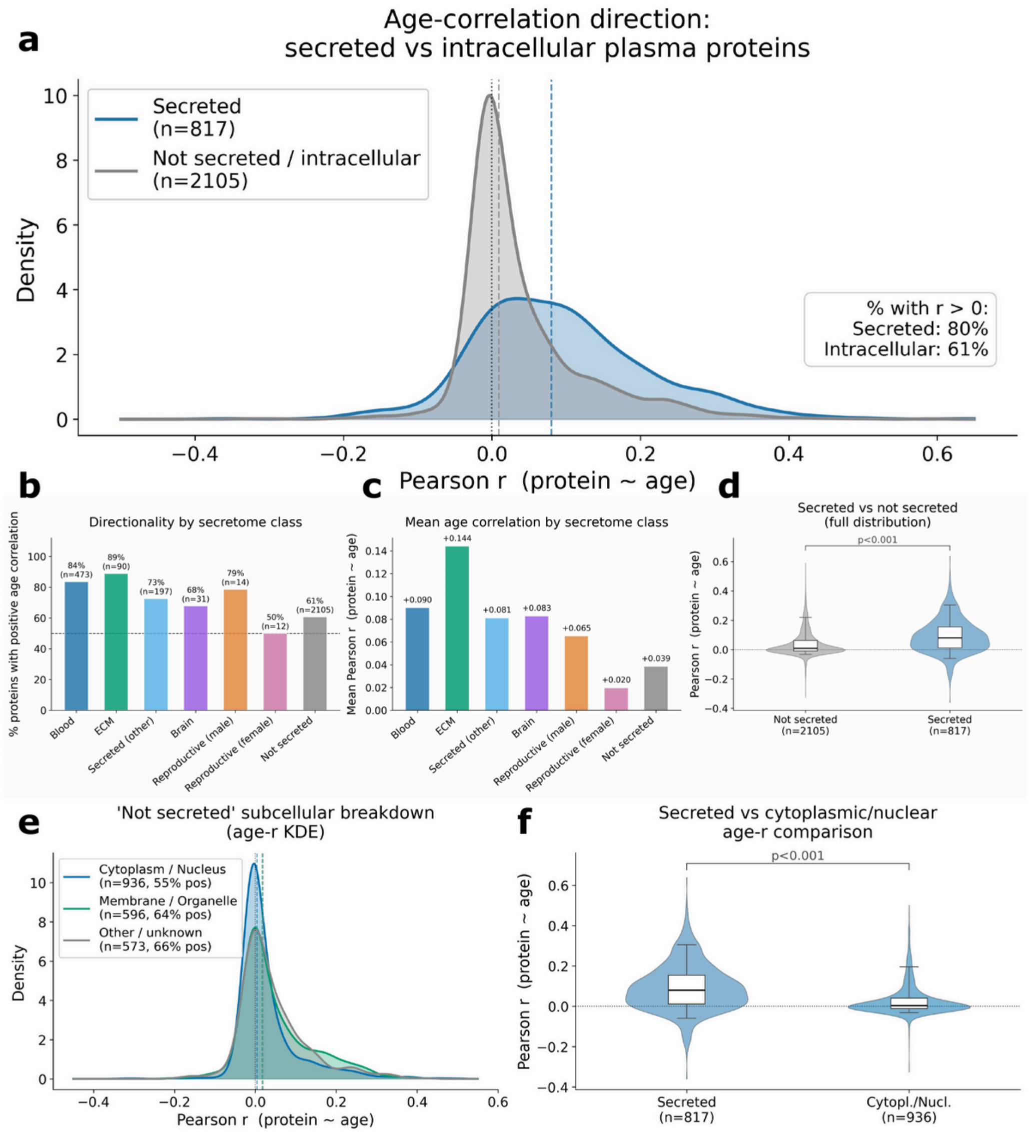


| Group | n | Mean r | Median r | % positive | Std |
|---|---|---|---|---|---|
| All secreted | 817 | +0.092 | +0.080 | 80% | 0.116 |
| Blood | 473 | +0.090 | +0.079 | 84% | 0.106 |
| ECM | 90 | +0.144 | +0.116 | 89% | 0.142 |
| Secreted (other) | 197 | +0.081 | +0.072 | 73% | 0.115 |
| Brain secreted | 31 | +0.083 | +0.062 | 68% | 0.131 |
| All not secreted | 2105 | +0.039 | +0.009 | 61% | 0.083 |
| Cytoplasm/Nucleus | 936 | +0.029 | +0.004 | 55% | 0.077 |
| Membrane/Organelle | 596 | +0.049 | +0.016 | 64% | 0.088 |
| Other/Unknown | 573 | +0.044 | +0.019 | 66% | 0.085 |

**Extended Data Fig. 4.** Age-correlation patterns of secreted versus intracellular plasma proteins. (a) Density distribution of Pearson correlations between protein level (NPX) and age for secreted (n=817, blue) and not-secreted/intracellular (n=2,105, gray) proteins; 80% of secreted vs. 61% of intracellular proteins show r>0. (b) Percentage of proteins with positive age correlation, broken out by secretome class (blood, ECM, secreted other, brain, reproductive male/female, not secreted). (c) Mean Pearson r (protein  age) by secretome class. (d) Full distribution of age correlations for secreted vs. not-secreted proteins (violin + box; p<0.001). (e) Subcellular breakdown of the not-secreted compartment (cytoplasm/nucleus, membrane/organelle, other/unknown) shown as age-r KDEs. (f) Direct comparison of age correlations between secreted (n=817) and cytoplasmic/nuclear (n=936) proteins (p<0.001). Summary statistics for all groups are tabulated below the panels.

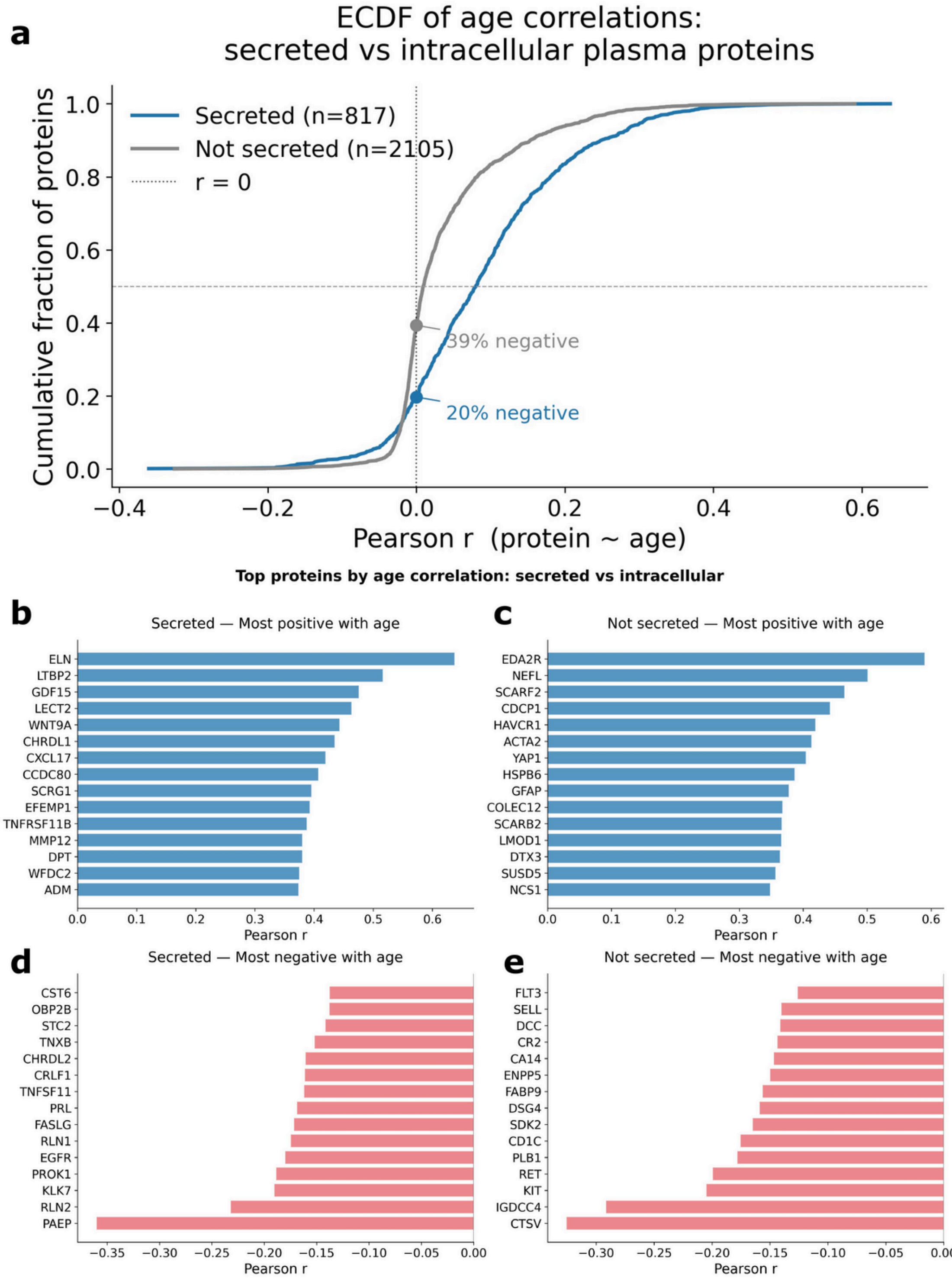


**Extended Data Fig. 5.** Top age-associated proteins among secreted and intracellular plasma proteins. (a) Empirical cumulative distribution function (ECDF) of age correlations for secreted (blue, n=817) and not-secreted (gray, n=2,105) proteins; 20% of secreted vs. 39% of intracellular proteins are negatively correlated with age. (b–c) Top 15 proteins with most positive age correlation among secreted (b) and not-secreted (c) compartments. (d–e) Top 15 proteins with most negative age correlation among secreted (d) and not-secreted (e) compartments.

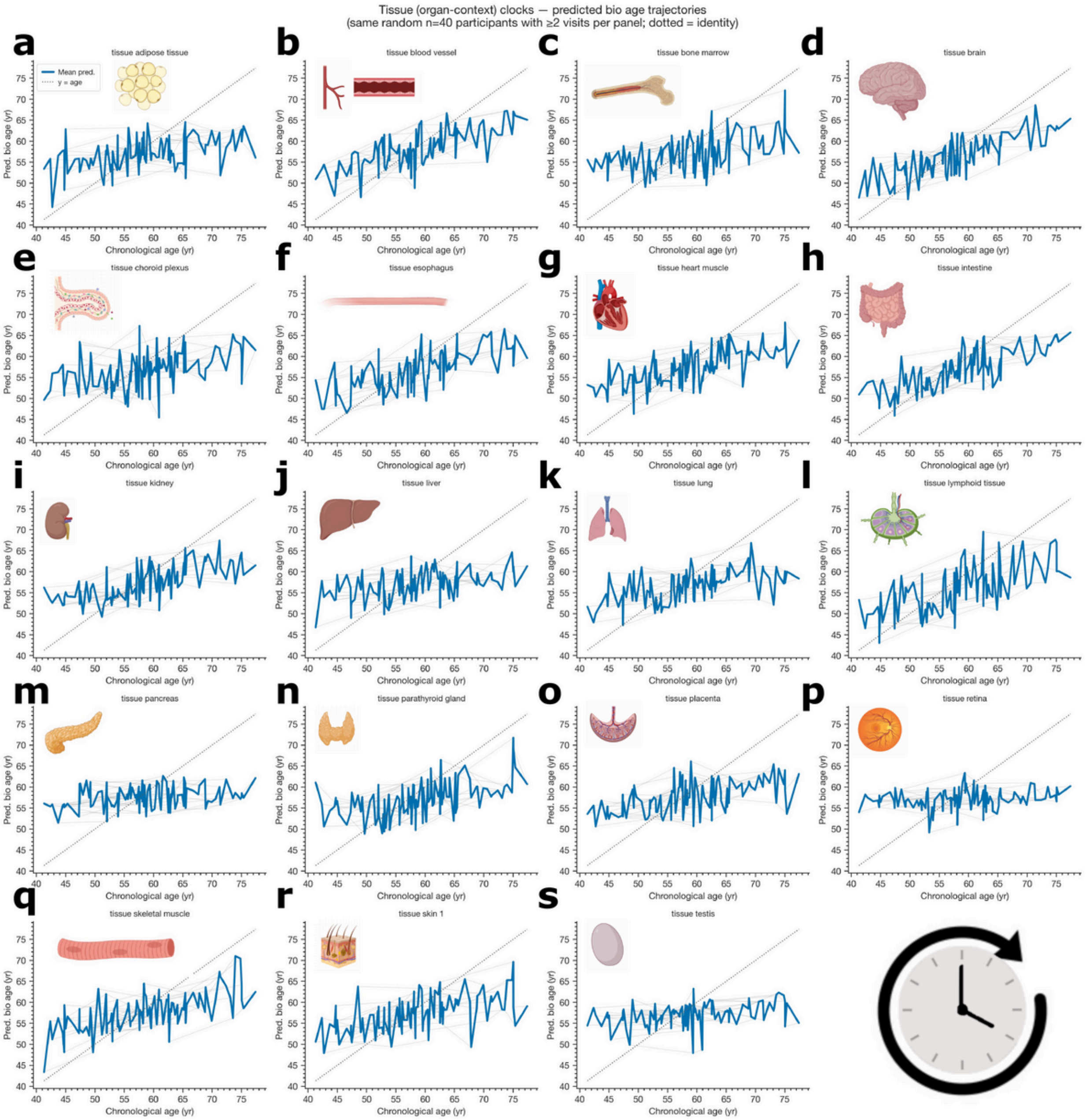


**Extended Data Fig. 6.** Longitudinal trajectories of tissue (organ-context) proteomic aging clocks. (a–s) Predicted biological age versus chronological age across repeated visits for the same random subset of n=40 participants with ≥2 proteomic visits per panel, shown for adipose tissue (a), blood vessel (b), bone marrow (c), brain (d), choroid plexus (e), esophagus (f), heart muscle (g), intestine (h), kidney (i), liver (j), lung (k), lymphoid tissue (l), pancreas (m), parathyroid gland (n), placenta (o), retina (p), skeletal muscle (q), skin (r), and testis (s). Thick blue lines are mean predicted ages; thin gray lines connect within-participant longitudinal visits; dotted diagonals indicate y=x (perfect tracking).

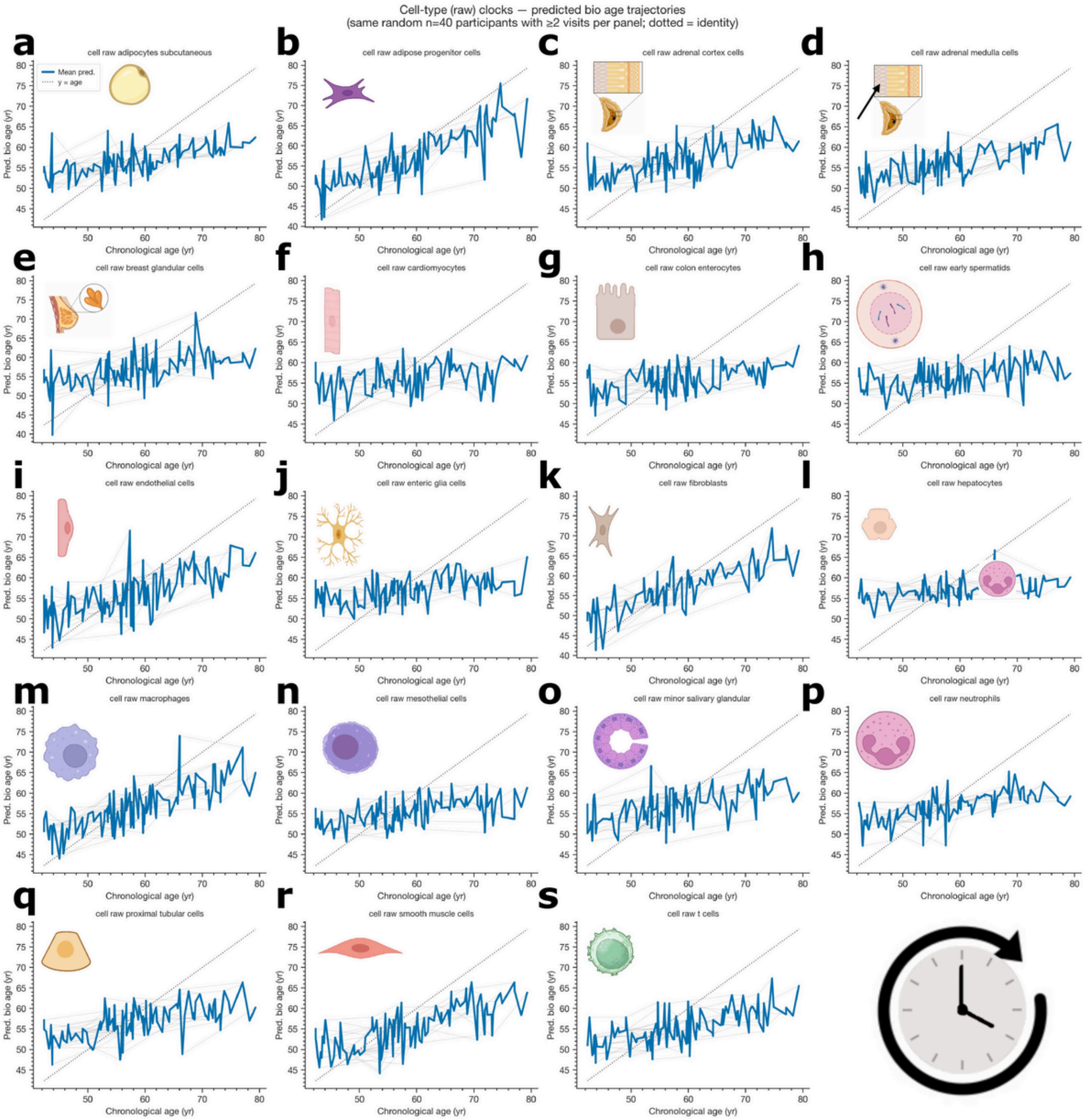


**Extended Data Fig. 7.** Longitudinal trajectories of raw cell-type-specific proteomic aging clocks. (a–s) Predicted biological age versus chronological age across repeated visits for the same random subset of n=40 participants with ≥2 proteomic visits per panel, shown for adipocytes (subcutaneous) (a), adipose progenitor cells (b), adrenal cortex cells (c), adrenal medulla cells (d), breast glandular cells (e), cardiomyocytes (f), colon enterocytes (g), early spermatids (h), endothelial cells (i), enteric glia cells (j), fibroblasts (k), hepatocytes (l), macrophages (m), mesothelial cells (n), minor salivary glandular cells (o), neutrophils (p), proximal tubular cells (q), smooth muscle cells (r), and T cells (s). Thick blue lines are mean predicted ages; thin gray lines connect within-participant longitudinal visits; dotted diagonals indicate y=x (perfect tracking).

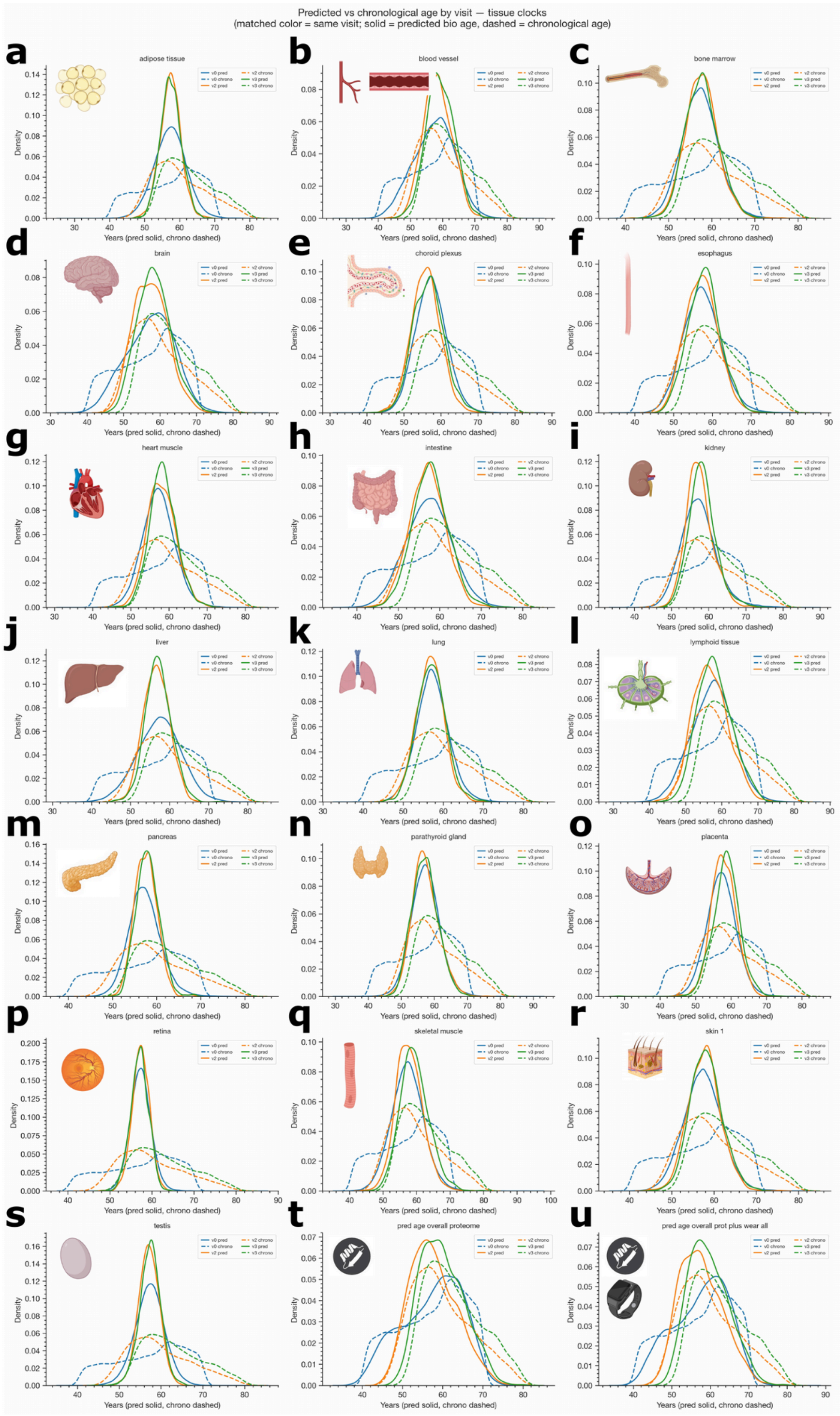


**Extended Data Fig. 8.** Distributions of predicted biological age versus chronological age across tissue clocks by visit. (a–u) Density distributions of predicted biological age (solid lines) and chronological age (dashed lines), color-matched by visit (v0, v2, v3), shown for adipose tissue (a), blood vessel (b), bone marrow (c), brain (d), choroid plexus (e), esophagus (f), heart muscle (g), intestine (h), kidney (i), liver (j), lung (k), lymphoid tissue (l), pancreas (m), parathyroid gland (n), placenta (o), retina (p), skeletal muscle (q), skin (r), testis (s), the overall proteome reference clock (t), and the overall proteome plus wearables clock (u).

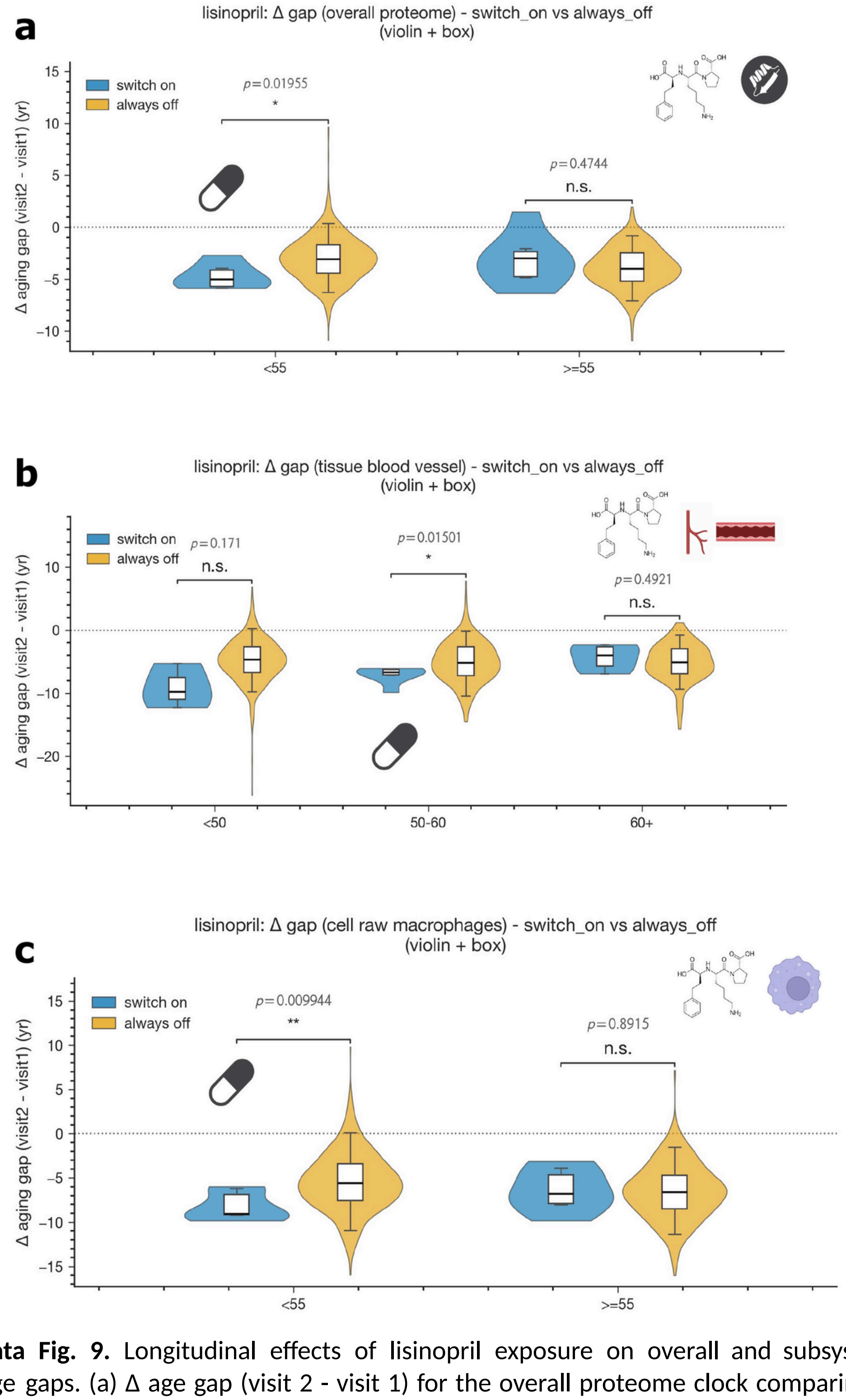


**Extended Data Fig. 9.** Longitudinal effects of lisinopril exposure on overall and subsystem-specific proteomic age gaps. (a) Δ age gap (visit 2 - visit 1) for the overall proteome clock comparing switch-on (blue) vs. always-off (orange) participants, stratified by age (<55, ≥55). (b) Δ age gap for the blood vessel tissue clock, stratified by age (<50, 50–60, 60+). (c) Δ age gap for the raw macrophage cell clock, stratified by age (<55, ≥55). Violins with embedded box plots; Welch p-values and significance annotations shown above each comparison.

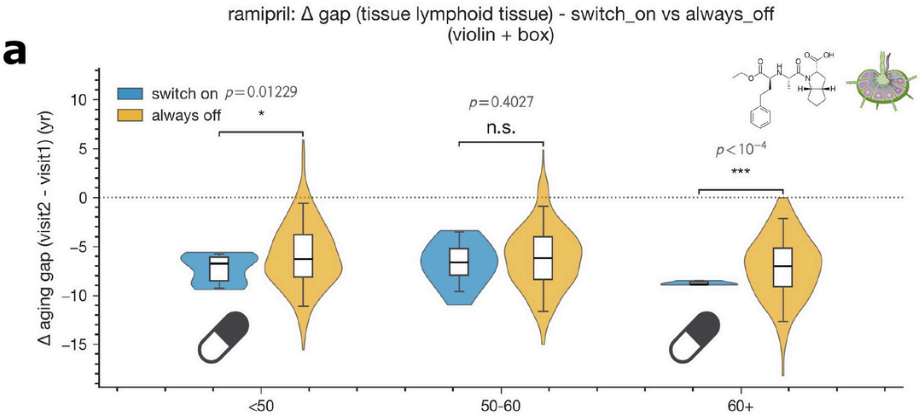


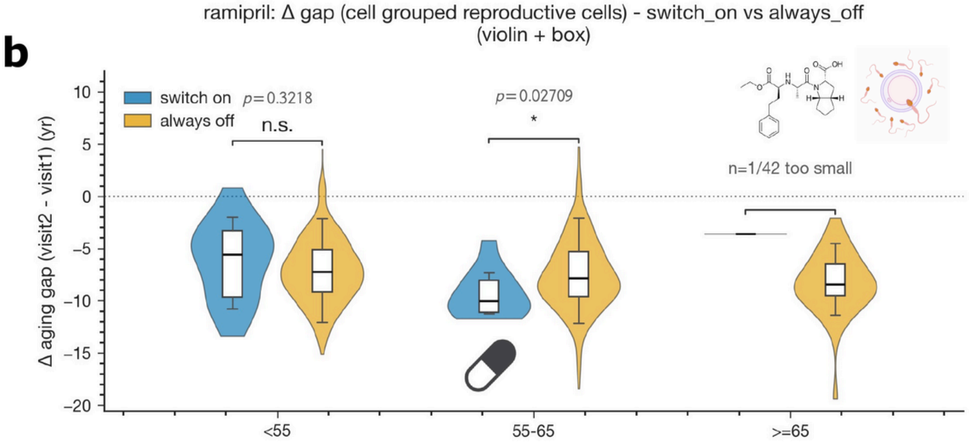


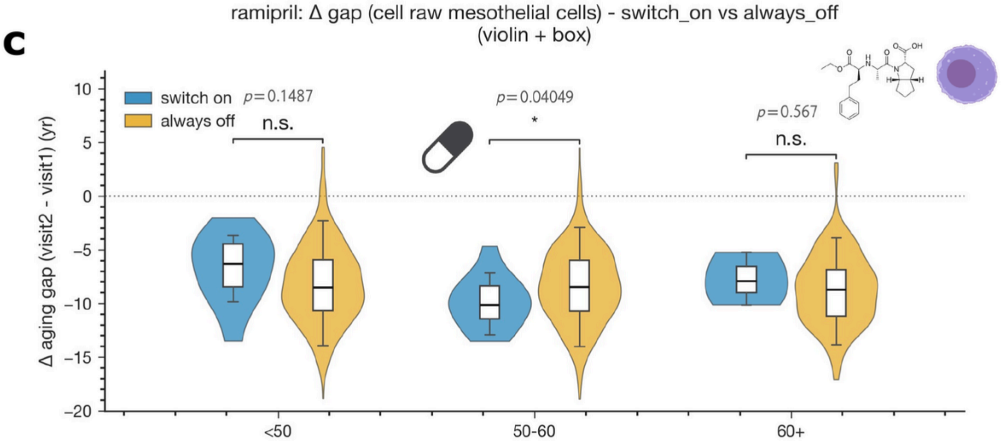


**Extended Data Fig. 10.** Longitudinal effects of ramipril exposure on subsystem-specific proteomic age gaps. (a) Δ age gap (visit 2 - visit 1) for the lymphoid tissue clock comparing switch-on (blue) vs. always-off (orange) participants, stratified by age (<50, 50–60, 60+). (b) Δ age gap for the grouped reproductive-cell clock, stratified by age (<55, 55–65, ≥65). (c) Δ age gap for the raw mesothelial-cell clock, stratified by age (<50, 50–60, 60+). Violins with embedded box plots; Welch p-values and significance annotations shown above each comparison.

### Supplementary Figure and Tables

#### *Supplementary Figure and Table Captions*

**Supplementary Fig. 1.** Olink protein-panel secretome and drug-target coverage. Horizontal bar chart of the 2,922-protein Olink panel showing, for each functional annotation, the number (and percentage of the full panel) of proteins that qualify: present in the HPA atlas (n=2,885; 98.7%), secreted (n=817; 28.0%), detected in blood (n=1,765; 60.4%), in the druggable genome (n=1,204; 41.2%), and FDA-approved drug target (n=276; 9.4%). Gray bars and labels show the number of panel proteins not meeting each criterion.

**Supplementary Fig. 2.** Flow of Olink panel proteins from tissue origin to drug status. Sankey diagram tracing all n=2,922 Olink proteins through five annotation stages: source-tissue category (gene-level: tissue enhanced, tissue enriched, group enriched, unknown), secretome location (blood, ECM, secreted other, not secreted), blood detection (detected in blood vs. not in blood), druggable target class (enzymes, transporters, CD markers, not druggable), and drug status (FDA-approved target, druggable but no drug, not targeted). Node labels include the number of proteins in each stratum.

**Supplementary Fig. 3.** Cohort overlap across UK Biobank modalities used in this study. Three-way Venn diagram of UKB participants with accelerometry (red), proteomics (green), and mortality (teal) data. Region counts: accelerometry only (n=86,941), proteomics only (n=35,582), mortality only (n=44,253), accelerometry proteomics (n=10,310), accelerometry mortality (n=5,561), proteomics mortality (n=6,387), and all three modalities (n=760). Total unique participants: 189,794.

**Supplementary Fig. 4.** UK Biobank cohort demographics, lifestyle, and covariate distributions. (a–g) Selection, geography, socioeconomic position, and ethnicity context: age at recruitment (a; N=501,936), sex (b), year of birth (c), Townsend area-deprivation index (d), ethnic background (e; log x-axis), top 12 baseline assessment centres (f), and best-endorsed education level (g). (h–n) Lifestyle, polygenic axes, substudy selection, and covariate overlap: BMI at baseline (h), smoking status (i), alcohol frequency (j), sleep duration (k), PRS genetic principal components for array 0 (l), substudy participation rates for accelerometry (CWA) and Olink proteomics (m; proteomics N=53,039), and Spearman correlation matrix among continuous covariates (age, birth year, Townsend, BMI, sleep, PRS PC) (n).

**Supplementary Fig. 5.** Sex-stratified overall proteomic aging clock performance. (a) Predicted vs. chronological age in female participants (r=0.927, MAE=2.43 yr). (b) Predicted vs. chronological age in male participants (r=0.932, MAE=2.39 yr). Dashed line in each panel is the identity (y=x).

**Supplementary Fig. 6.** Feature contributions to the multimodal chronological age clock. (a) Pie chart showing the share of total coefficients contributed by each modality: proteomics 91.0% (1,126 features), clinical 3.8% (36), wearables 3.2% (32), multimorbidities 1.6% (21), digital 0.3% (1), demographics 0.1% (1). (b) Number of non-zero features per modality (log scale). (c) Top 25 positive (ages-faster) and bottom 25 negative (ages-slower) ElasticNet coefficients, color-coded by modality (proteomics blue, clinical green, digital orange, wearables yellow).

**Supplementary Fig. 7.** Predicting time-to-death from proteomics with and without wearables. (a) Predicted time-to-death (OLS, years) versus chronological age for the proteomics + wearables model on the 80/20 train/test split (train n=8,864; test n=2,187), with C=0.814/0.782, r=-0.681/-0.707, and test MAE(ttd)=0.19 yr. (b) Predicted time-to-death versus chronological age for the proteomics-only model (train n=42,386; test n=10,597), with C=0.817/0.811, r=-0.681/-0.693, and test MAE(ttd)=0.52 yr. Restricted to the 100%-estimable subset.

**Supplementary Fig. 8.** Feature contributions to the multimodal proteomics + wearables mortality model. (a) Pie chart showing the share of total HR by modality: proteomics 90.3% (153 features), wearables 9.7% (17 features). (b) Number of non-zero features per modality (log scale). (c) Top 25 positive (higher mortality risk) and bottom 25 negative (lower mortality risk) Cox log hazard ratios, color-coded by modality (proteomics blue, wearables yellow), with HR values annotated per feature.

**Supplementary Fig. 9.** Relationship between participant age and blood sample collection time across UKB instances. (a–d) Hexbin density plots of blood draw time (UKB field 3166, hour of day) versus age at visit for instance 0 (a; n=496,822, Spearman ρ=-0.007, Pearson r=-0.003), instance 1 (b; n=20,156, ρ=0.006, r=0.009), instance 2 (c; n=97,944, ρ=0.116, r=0.119), and instance 3 (d; n=20,275, ρ=0.086, r=0.090).

**Supplementary Fig. 10.** Three-tier proteomic aging and mortality clock atlas across the body (UKB Olink, n≈53k, male + female). (a) Tier 1 — whole-body anatomical mapping of age-clock $R^2$ across tissues. (b) Tier 2 — cell-type age clock CV-$R^2$ (12 grouped cell types: adipose, fibroblasts, immune, adrenal, smooth muscle, epithelial, endothelial, mesothelial, liver, cardiomyocytes, reproductive, glia). (c) Tier 3 — organelle-level age clock $R^2$ overlaid on a stylized cell schematic. (d) Tier 1 — whole-body anatomical mapping of mortality C-index (CoxNet) across tissues. (e) Tier 2 — cell-type mortality clock C-index across the same 12 grouped cell types. (f) Tier 3 — organelle-level mortality C-index overlaid on a stylized cell schematic.

**Supplementary Fig. 11.** Tissue-resolved proteomic age clock $R^2$ rendered on male and female anatomical templates (protein subset by HPA tissue annotation; ElasticNet CV; background = overall $R^2$=0.863). (a) Male whole-body map. (b) Female whole-body map. Tissues are colored by tissue-specific clock $R^2$ (CV). The accompanying table lists per-tissue $R^2$ values, ranging from brain (0.600) and blood vessel (0.561) at the top to pancreas (0.178) and testis (0.183) at the bottom.

**Supplementary Fig. 12.** Tissue-resolved proteomic mortality clock C-index rendered on male and female anatomical templates (CoxNet, n=52,983, 7,145 deaths). (a) Male whole-body map. (b) Female whole-body map. Tissues are colored by tissue-specific Cox C-index. The accompanying table lists per-tissue C-index values, with brain (0.787), lymphoid tissue (0.766), and blood vessel (0.758) at the top and retina (0.608) and testis (0.650) at the bottom.

**Supplementary Fig. 13.** Organelle-level proteomic age clock $R^2$ rendered on a stylized cell schematic (UKB Olink, N≈53k, ElasticNet CV). Subcellular compartments are colored by their compartment-specific age-clock $R^2$ (CV). Compartments backed by fewer than five panel proteins are shown with hatched fill.

**Supplementary Fig. 14.** Organelle-level proteomic mortality clock C-index rendered on a stylized cell schematic (UKB Olink, N≈53k, CoxNet). Subcellular compartments are colored by their compartment-specific Cox C-index (CV). Compartments backed by fewer than five panel proteins are shown with hatched fill.

**Supplementary Fig. 15.** Organelle-level clock performance ranked across subcellular compartments (UKB plasma proteomics, Olink). (a) Organelle age clock ElasticNet $R^2$ (5-fold CV), led by plasma membrane (0.624), cytosol (0.593), vesicles (0.578), nucleoplasm (0.574), and Golgi apparatus (0.477). (b) Organelle mortality clock Cox C-index (CV), led by plasma membrane (0.782), nucleoplasm (0.779), vesicles (0.775), cytosol (0.770), and Golgi apparatus (0.760); dashed line at C=0.5 indicates chance performance.

**Supplementary Fig. 16.** Top age-associated plasma proteins in the UKB Olink panel. Ranked Pearson correlation between protein level and chronological age, showing the 20 most positively correlated proteins (red, top, led by ELN, EDA2R, LTBP2, NEFL, GDF15) and the 20 most negatively correlated proteins (blue, bottom, led by PAEP, CTSV, IGDCC4, RLN2, KIT).

**Supplementary Fig. 17.** Age-dependent decoupling of plasma protein–age associations and cross-modal comparison with single-cell transcriptomics. (a) Per-protein Pearson correlation with age computed in younger (x-axis) vs. older (y-axis) UKB participants; dashed diagonal indicates equal correlation across age strata. (b) Top decoupled proteins ranked by Δ r (old - young), with positive shifts (red, e.g. PAEP, PROK1, RLN2, CHRDL2) and negative shifts (blue, e.g. FSHB, CGA, ELN, DPP4) indicating proteins whose age relationship strengthens or weakens with chronological age. (c) Cross-modal scatter of UKB plasma protein–age correlations versus Tabula Sapiens (TS) gene–age correlations across matched gene–tissue pairs (Pearson r=-0.020, n=57,593), illustrating limited transfer of bulk transcriptomic age signal to plasma proteomic age signal.

**Supplementary Fig. 18.** Mean within-person slopes (Δ predicted bio age / Δ chronological age) across all proteomic aging clock variants. (a) Reference clocks built on the full proteome panel (proteome-only and proteome + wearables). (b) Tissue (organ-context) clocks ranked across organs/tissues, led by brain, blood vessel, and skeletal muscle, and trailing in retina and testis. (c) Secretome clocks (not secreted, ECM, blood, secreted other, brain). (d) HPA cell-grouped clocks across 12 grouped cell types, led by adipose cells, fibroblasts, and epithelial cells. (e) HPA cell-type (raw) clocks across 19 cell types, led by fibroblasts, adipose progenitor cells, and macrophages. Dashed line at 1.0 yr/yr indicates perfect longitudinal tracking.

**Supplementary Fig. 19.** Mapping UKB-reported medications to FDA-approved Olink targets. Three-way Venn diagram of phase-4 drug-target genes derived from UKB self-reported medications (orange, mapped via ChEMBL), GP-script medications (purple, mapped via ChEMBL), and the FDA-flagged subset of the Olink panel from HPA (green, n=276). Overlaps: 431 genes shared between UKB self-report and GP scripts, 51 across all three sources, 62 between GP scripts and FDA-Olink, and 6 unique to UKB self-report; 329 GP-script-only and 160 FDA-Olink-only.

**Supplementary Fig. 20.** Baseline characteristics and longitudinal aging-gap differences by FDA-target drug exposure (overall proteome). (a) Chronological age at first Olink visit for switch-on (blue) vs. always-off (orange) participants based on the UKB p20003 FDA-Olink target drug flag (Welch $p=1.79\times10^{-4}$). (b) Δ age gap (visit 2 - visit 1) from the overall proteome clock, switch-on vs. always-off, stratified by age group (<55, 55–65, ≥65); none of the per-stratum comparisons reach significance.

**Supplementary Fig. 21.** Lifestyle/exposure-stratified within-person slopes and baseline age gaps, FDA Olink target-drug exposure (UKB/GP, ChEMBL ph.4). (a–d) Proteomics + accelerometry clock (baseline wearers, all participants), comparing participants with no matched FDA-Olink drug vs. ≥1 matched drug: within-person Δgap slope (a), within-person predicted-age slope (b), baseline (visit 0) age gap (c), and baseline predicted bio age (d). (e–h) Overall proteome clock (full NPX panel), with the same four readouts under the same drug exposure stratification.

**Supplementary Fig. 22.** Diabetic-cohort second-omics aging-gap analysis using cell-specific clocks: switch-on vs. always-off at outcome visit, stratified by age (<55, 55–65, ≥65). (a) Age gap from the cell-grouped epithelial-cell clock at the outcome Olink visit; no significant differences across age strata. (b) Age gap from the raw minor salivary glandular cell clock at the outcome Olink visit; no significant differences across age strata.

**Supplementary Fig. 23.** Diabetic-cohort second-omics predicted-age analysis using cell-specific clocks: switch-on vs. always-off at outcome visit, stratified by age (<55, 55–65, ≥65). (a) Predicted age from the cell-grouped epithelial-cell clock; switch-on participants show significantly lower predicted age in the 55–65 stratum (p=0.0074). (b) Predicted age from the raw minor salivary glandular cell clock; switch-on participants show significantly lower predicted age in the 55–65 stratum (p=0.0040).

**Supplementary Fig. 24.** Diabetic-cohort second-omics mortality analysis using cell-specific clocks: median predicted time-to-death (TTD from fitted Cox survival) at outcome visit, switch-on vs. always-off, stratified by age (<55, 55–65, ≥65). (a) Cell-grouped epithelial-cell clock; switch-on participants show significantly higher predicted TTD across all three age strata (p=0.027, $p<10^{-4}$, p=0.024). (b) Raw minor salivary glandular cell clock; switch-on participants show significantly higher predicted TTD across all three age strata (p=0.016, $p<10^{-4}$, p=0.018).

**Supplementary Fig. 25.** Diabetic-cohort second-omics mortality analysis using cell-specific clocks: log partial hazard (from fitted Cox) at outcome visit, switch-on vs. always-off, stratified by age (<55, 55–65, ≥65). (a) Cell-grouped epithelial-cell clock; switch-on participants show significantly lower log partial hazard in the 55–65 ($p<10^{-4}$) and ≥65 (p=0.042) strata. (b) Raw minor salivary glandular cell clock; switch-on participants show significantly lower log partial hazard in the 55–65 stratum ($p<10^{-4}$).

**Supplementary Fig. 26.** Overlap between NIA ITP interventions, UKB medications, and FDA-flagged Olink-panel targeting drugs. Three-way Venn diagram of NIA ITP intervention drugs (green), UKB self-reported/GP-script medication names (orange), and ChEMBL phase-4 drugs targeting FDA-flagged Olink panel proteins (teal, the “bridge bundle”). 442 drugs are shared between UKB medications and the FDA-Olink targeting set, 3 are shared between ITP and the FDA-Olink targeting set, and 20 ITP interventions overlap with UKB medication names but not the FDA-Olink set. UKB-only (n=1,973) and FDA-Olink-targeting-only (n=337) regions are also shown.

**Supplementary Fig. 27.** Within-person slopes and baseline age gaps stratified by alcohol frequency. (a–d) Proteomics + accelerometry clock (baseline wearers, all participants) by alcohol frequency category (daily or almost daily, three or four times a week, once or twice a week, one to three times a month, special occasions only, never): within-person Δgap slope (a), within-person predicted-age slope (b), baseline age gap (c), and baseline predicted bio age (d). (e–h) Overall proteome clock (full NPX panel) under the same alcohol-frequency stratification, with the additional “prefer not to answer” category included in the baseline panels.

**Supplementary Fig. 28.** Within-person slopes and baseline age gaps stratified by smoking status. (a–d) Proteomics + accelerometry clock (baseline wearers, all participants) by smoking status (never smoked, previous smoker, current smoker): within-person Δgap slope (a), within-person predicted-age slope (b), baseline age gap (c), and baseline predicted bio age (d). Current smokers show the largest baseline age gap. (e–h) Overall proteome clock (full NPX panel) under the same smoking stratification.

**Supplementary Fig. 29.** Wearable accelerometry features in the trimodal Cox mortality model, ranked by hazard ratio. Lollipop plot of all 17 wearable features retained by the multimodal (proteomics + wearables) Cox proportional-hazards mortality model, color-coded as protective (HR<1, n=15, green) or risk (HR>1, n=2, orange). The 15 protective wearable features—led by no-wear-bias-adjusted average acceleration (HR=0.959), afternoon acceleration 17:00–18:00 (HR=0.964), overall daily average acceleration (HR=0.964), and late-afternoon acceleration 16:00–17:00 (HR=0.970)—are dominated by daytime moderate-intensity activity signals consistent with prior accelerometer mortality studies[28–31]. Only two wearable features show HR>1: midnight acceleration 00:00–01:00 (HR=1.006) and early-morning acceleration 04:00–05:00 (HR=1.001), both far below the protein-level top-25 risk-feature threshold (HR1.024) and likely reflecting nocturnal activity patterns associated with sleep disruption rather than independent biological risk. The dashed vertical line at HR=1 marks the neutral risk reference. This figure complements Fig. 1e by showing the full wearable-feature hazard-ratio spectrum that is otherwise truncated by the protein-dominated top-25 ranking.

**Supplementary Fig. 30.** Intrinsic capacity domain scores across age in UK Biobank participants, visualized as Hexbin density plots showing z-normalized intrinsic capacity (IC) domain scores as a function of age at visit for (a) vitality, (b) locomotion, (c) cognition, (d) psychological well-being, (e) sensory function, and (f) total intrinsic capacity. Color intensity represents log-scaled participant density (log(count)). Solid lines indicate linear regression fits across age, with corresponding Pearson correlation coefficients (r), p-values, and per-decade score changes (Δ/decade) shown in each panel. Most IC domains exhibit modest declines with increasing age, particularly sensory and locomotion function, whereas psychological scores show a slight positive association with age. Sample sizes for each domain are indicated in the lower-left corner of each panel.

**Table S1.** Intrinsic capacity (IC) domain items used in this study. Each item is direction-harmonized so that higher values indicate better capacity, z-scored, and averaged within domain. The domain score is the mean of all available standardized items (≥1 item required). Items marked ↓ were negated prior to z-scoring (higher raw value = worse capacity).

**Table S2.** Comparison of intrinsic capacity (IC) operationalization approaches in UK Biobank. This study implements IC as a continuous functional-reserve construct across 137 items; published UK Biobank studies use a deficit-accumulation approach that dichotomizes each domain into impaired/not-impaired and sums across domains. Both approaches share the same five WHO ICOPE domains (Beard et al. 2016; Cesari et al. 2018) but differ in indicator depth, scaling, and analytic purpose.

**Table S3.** Definition and inventory of the 49 mAge biological subsystems. Each row lists one mAge subsystem with its category (organ/tissue, secretome class, grouped cell type, raw cell type, or organelle), within-category index, the assignment rule used to map plasma proteins to that subsystem, whether assignment is exclusive or overlapping, the minimum number of panel proteins used to train the corresponding clock, and whether a per-subsystem age and/or mortality clock was trained. Because most plasma proteins map to multiple compartments under HPA annotation rules, the subsystems are non-exclusive by construction: this table makes those overlaps and minimum-feature thresholds explicit so that subsystem-specific performance differences are not over-interpreted as fully independent estimates of compartment-specific aging.
**Table S4.** ChEMBL drugs targeting cardiomyocyte-clock genes.
**Table S5.** Drug targets and effects on aging-related outcomes in diabetic participants**Table S6.** *Subsystem-specific clock proteins, performance, and optimization parameters.*

***Supplementary Figures***

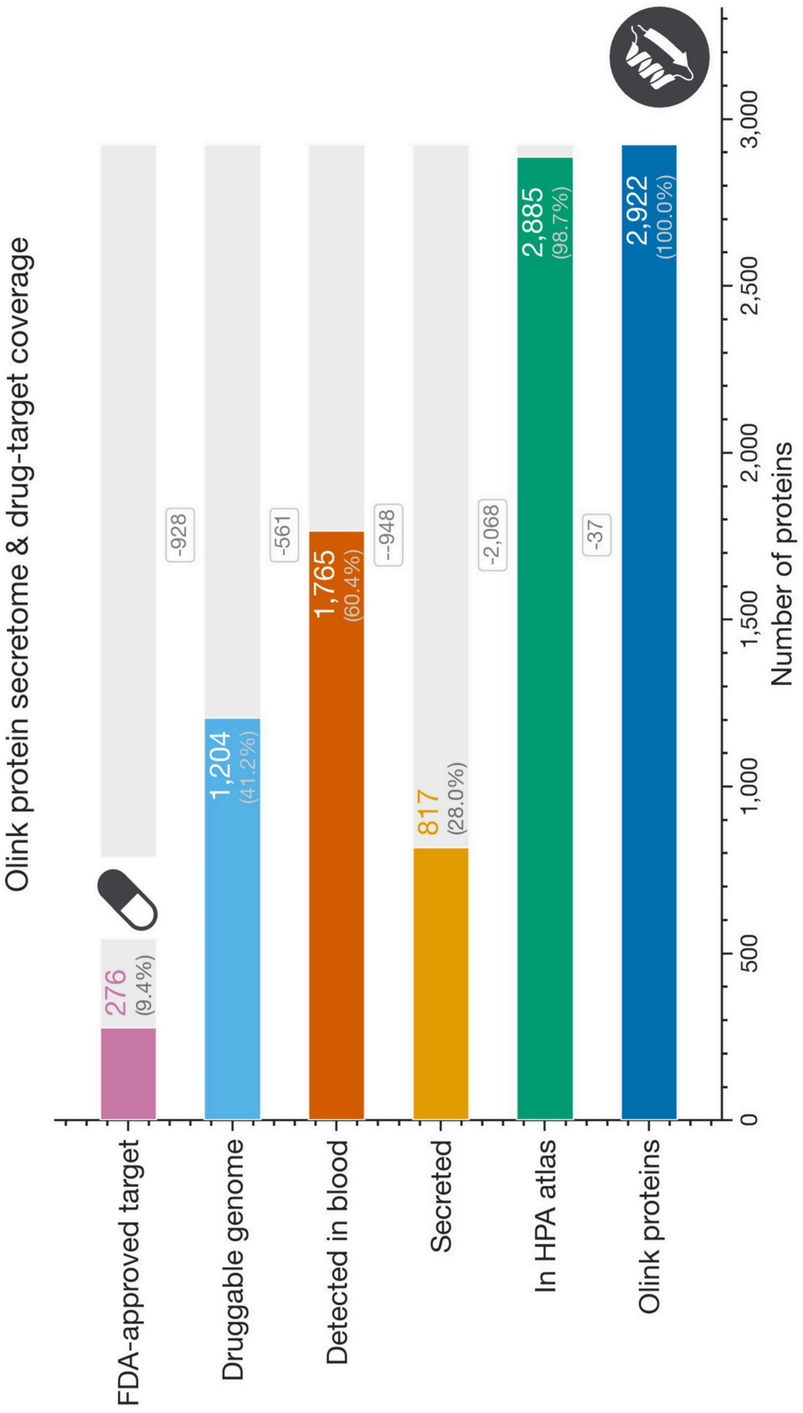


**Supplementary Fig. 1.** Olink protein-panel secretome and drug-target coverage. Horizontal bar chart of the 2,922-protein Olink panel showing, for each functional annotation, the number (and percentage of the full panel) of proteins that qualify: present in the HPA atlas (n=2,885; 98.7%), secreted (n=817; 28.0%), detected in blood (n=1,765; 60.4%), in the druggable genome (n=1,204; 41.2%), and FDA-approved drug target (n=276; 9.4%). Gray bars and labels show the number of panel proteins not meeting each criterion.

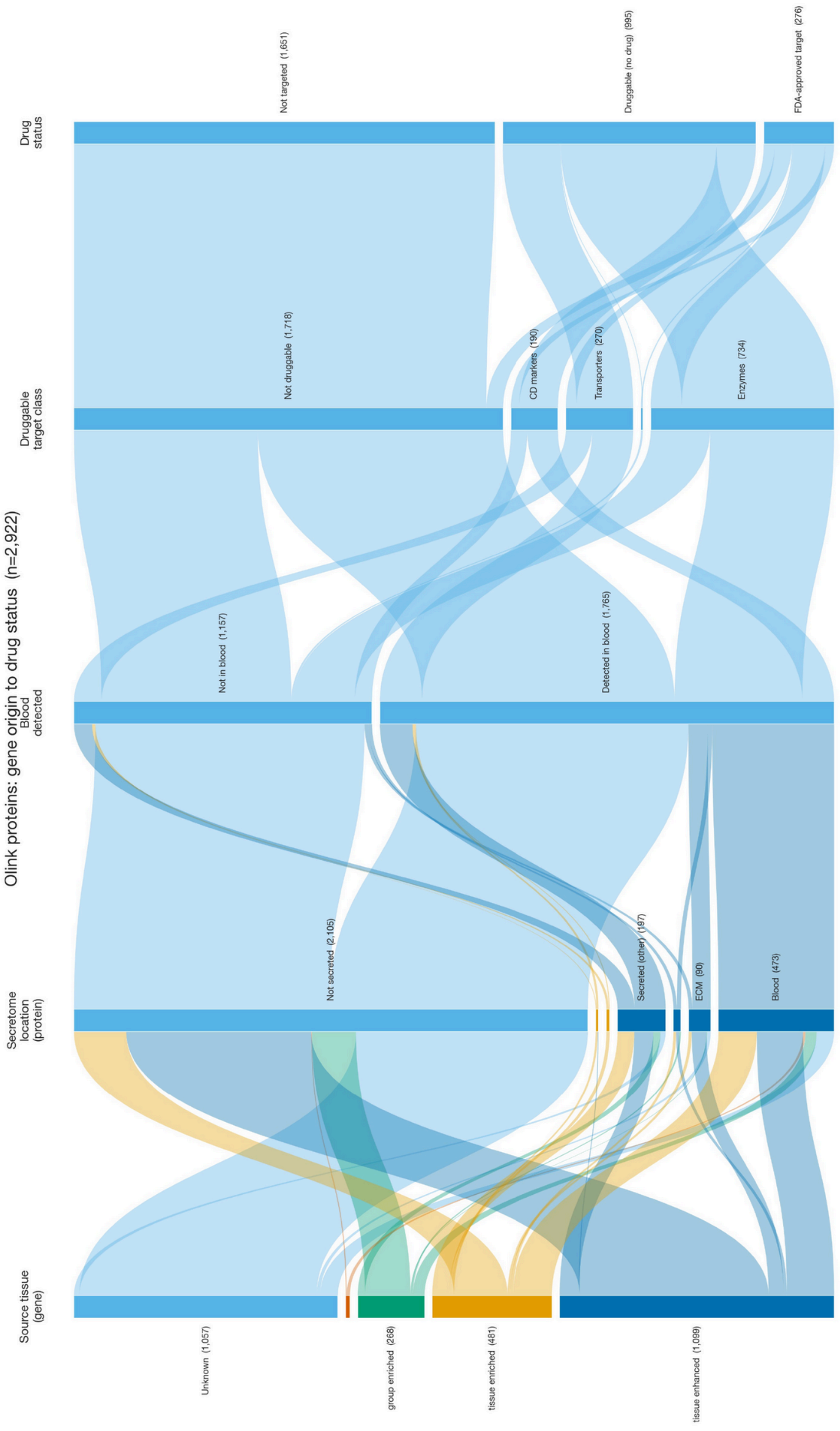


**Supplementary Fig. 2.** Flow of Olink panel proteins from tissue origin to drug status. Sankey diagram tracing all n=2,922 Olink proteins through five annotation stages: source-tissue category (gene-level: tissue enhanced, tissue enriched, group enriched, unknown), secretome location (blood, ECM, secreted other, not secreted), blood detection (detected in blood vs. not in blood), druggable target class (enzymes, transporters, CD markers, not druggable), and drug status (FDA-approved target, druggable but no drug, not targeted). Node labels include the number of proteins in each stratum.

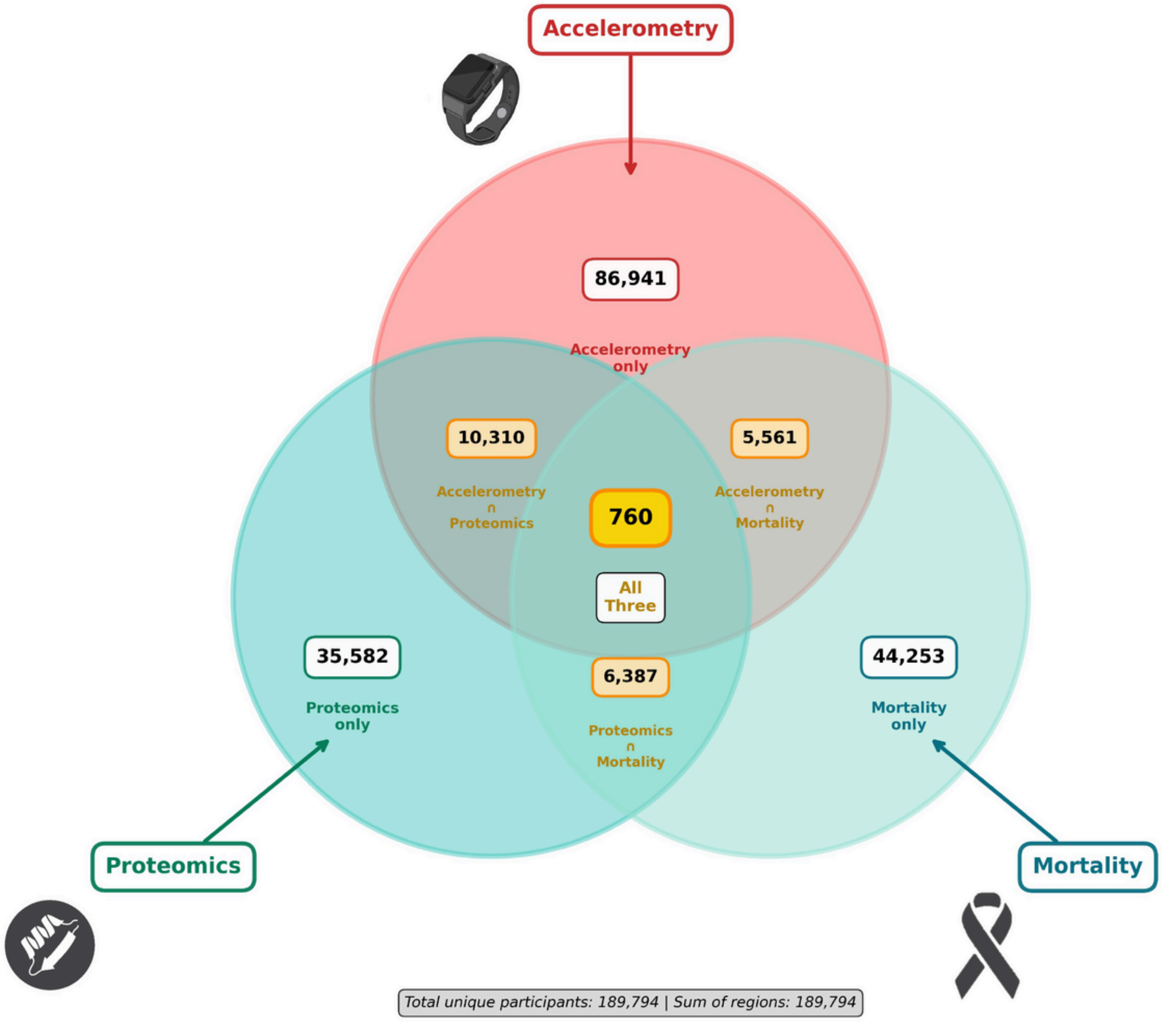


**Supplementary Fig. 3.** Cohort overlap across UK Biobank modalities used in this study. Three-way Venn diagram of UKB participants with accelerometry (red), proteomics (green), and mortality (teal) data. Region counts: accelerometry only (n=86,941), proteomics only (n=35,582), mortality only (n=44,253), accelerometry proteomics (n=10,310), accelerometry mortality (n=5,561), proteomics mortality (n=6,387), and all three modalities (n=760). Total unique participants: 189,794.

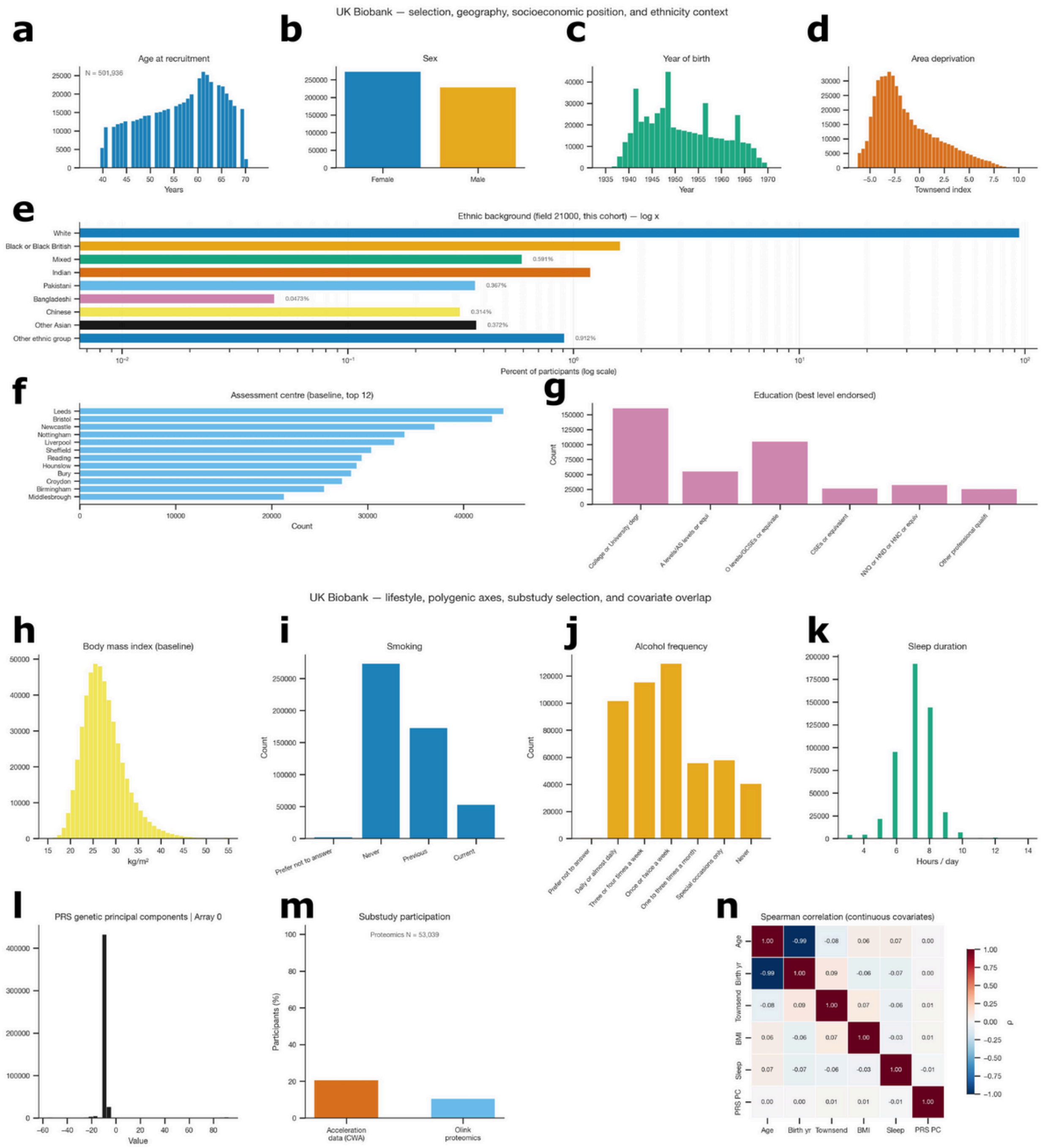


**Supplementary Fig. 4.** UK Biobank cohort demographics, lifestyle, and covariate distributions. (a–g) Selection, geography, socioeconomic position, and ethnicity context: age at recruitment (a; N=501,936), sex (b), year of birth (c), Townsend area-deprivation index (d), ethnic background (e; log x-axis), top 12 baseline assessment centres (f), and best-endorsed education level (g). (h–n) Lifestyle, polygenic axes, substudy selection, and covariate overlap: BMI at baseline (h), smoking status (i), alcohol frequency (j), sleep duration (k), PRS genetic principal components for array 0 (l), substudy participation rates for accelerometry (CWA) and Olink proteomics (m; proteomics N=53,039), and Spearman correlation matrix among continuous covariates (age, birth year, Townsend, BMI, sleep, PRS PC) (n).

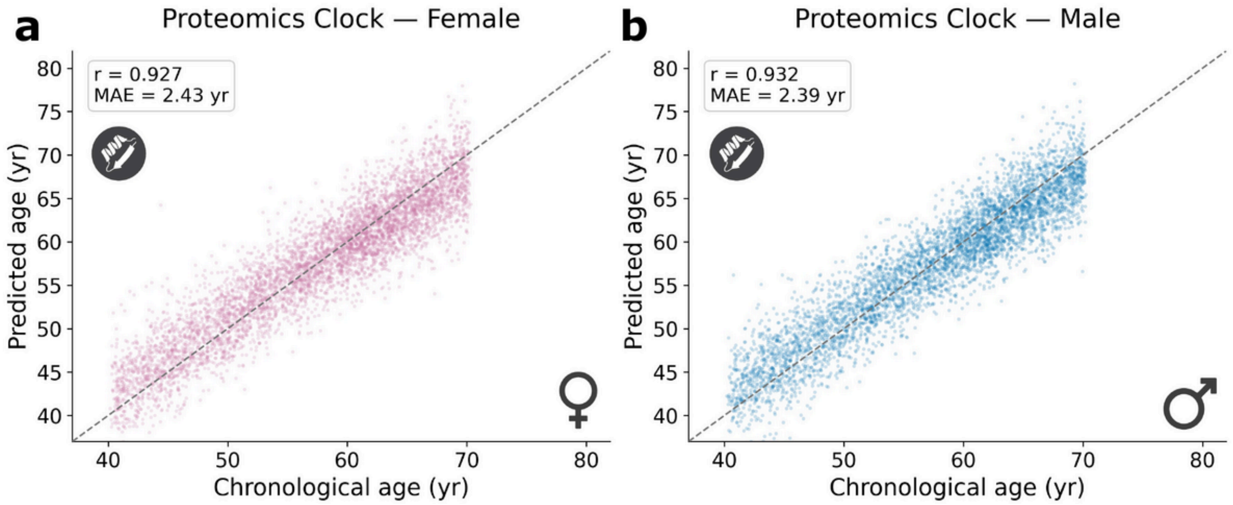


**Supplementary Fig. 5.** Sex-stratified overall proteomic aging clock performance. (a) Predicted vs. chronological age in female participants (r=0.927, MAE=2.43 yr). (b) Predicted vs. chronological age in male participants (r=0.932, MAE=2.39 yr). Dashed line in each panel is the identity (y=x).

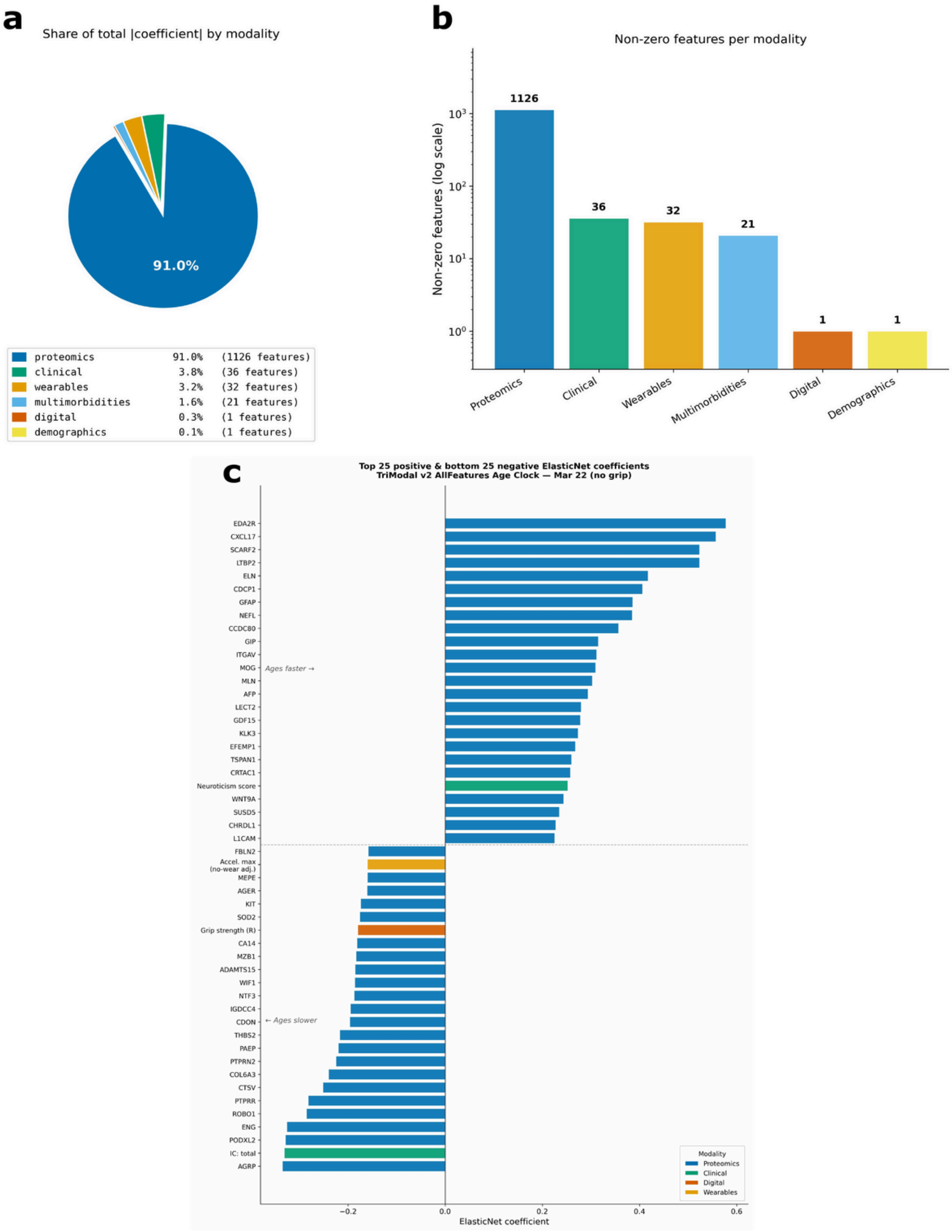


**Supplementary Fig. 6.** Feature contributions to the multimodal chronological age clock. (a) Pie chart showing the share of total coefficients contributed by each modality: proteomics 91.0% (1,126 features), clinical 3.8% (36), wearables 3.2% (32), multimorbidities 1.6% (21), digital 0.3% (1), demographics 0.1% (1). (b) Number of non-zero features per modality (log scale). (c) Top 25 positive (ages-faster) and bottom 25 negative (ages-slower) ElasticNet coefficients, color-coded by modality (proteomics blue, clinical green, digital orange, wearables yellow).

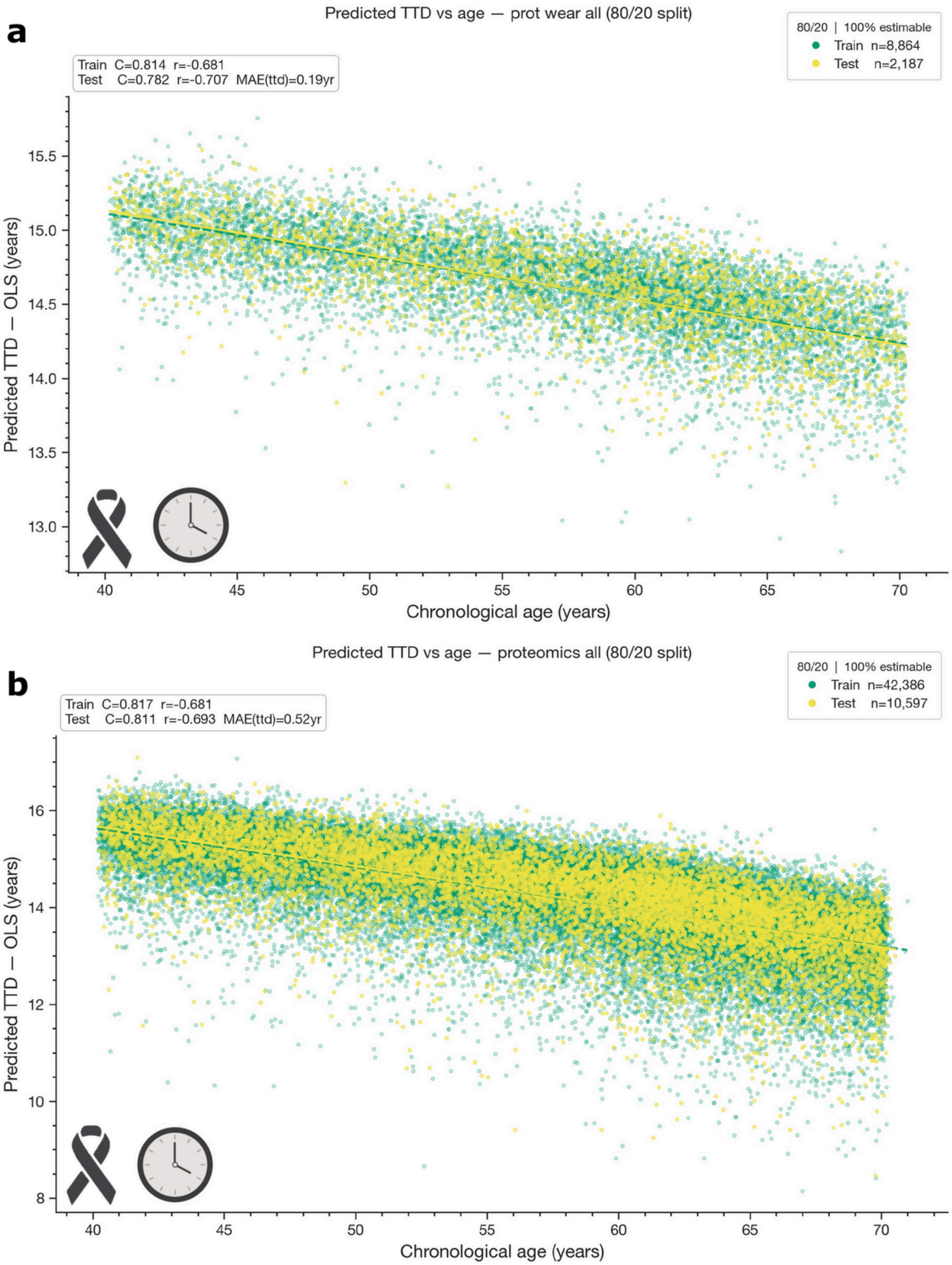


**Supplementary Fig. 7.** Predicting time-to-death from proteomics with and without wearables. (a) Predicted time-to-death (OLS, years) versus chronological age for the proteomics + wearables model on the 80/20 train/test split (train n=8,864; test n=2,187), with C=0.814/0.782, r=-0.681/-0.707, and test MAE(ttd)=0.19 yr. (b) Predicted time-to-death versus chronological age for the proteomics-only model (train n=42,386; test n=10,597), with C=0.817/0.811, r=-0.681/-0.693, and test MAE(ttd)=0.52 yr. Restricted to the 100%-estimable subset.

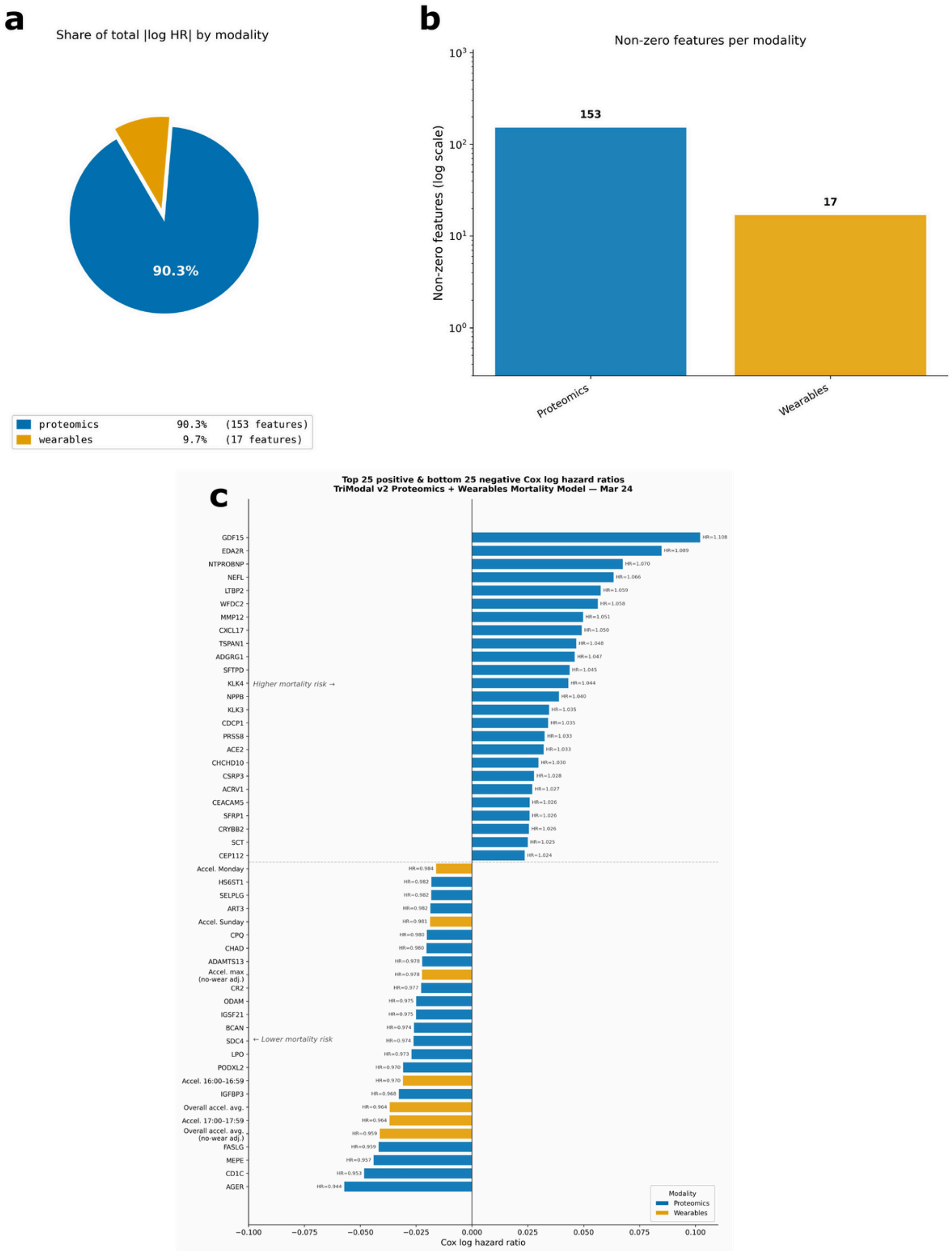


**Supplementary Fig. 8.** Feature contributions to the multimodal proteomics + wearables mortality model. (a) Pie chart showing the share of total HR by modality: proteomics 90.3% (153 features), wearables 9.7% (17 features). (b) Number of non-zero features per modality (log scale). (c) Top 25 positive (higher mortality risk) and bottom 25 negative (lower mortality risk) Cox log hazard ratios, color-coded by modality (proteomics blue, wearables yellow), with HR values annotated per feature.

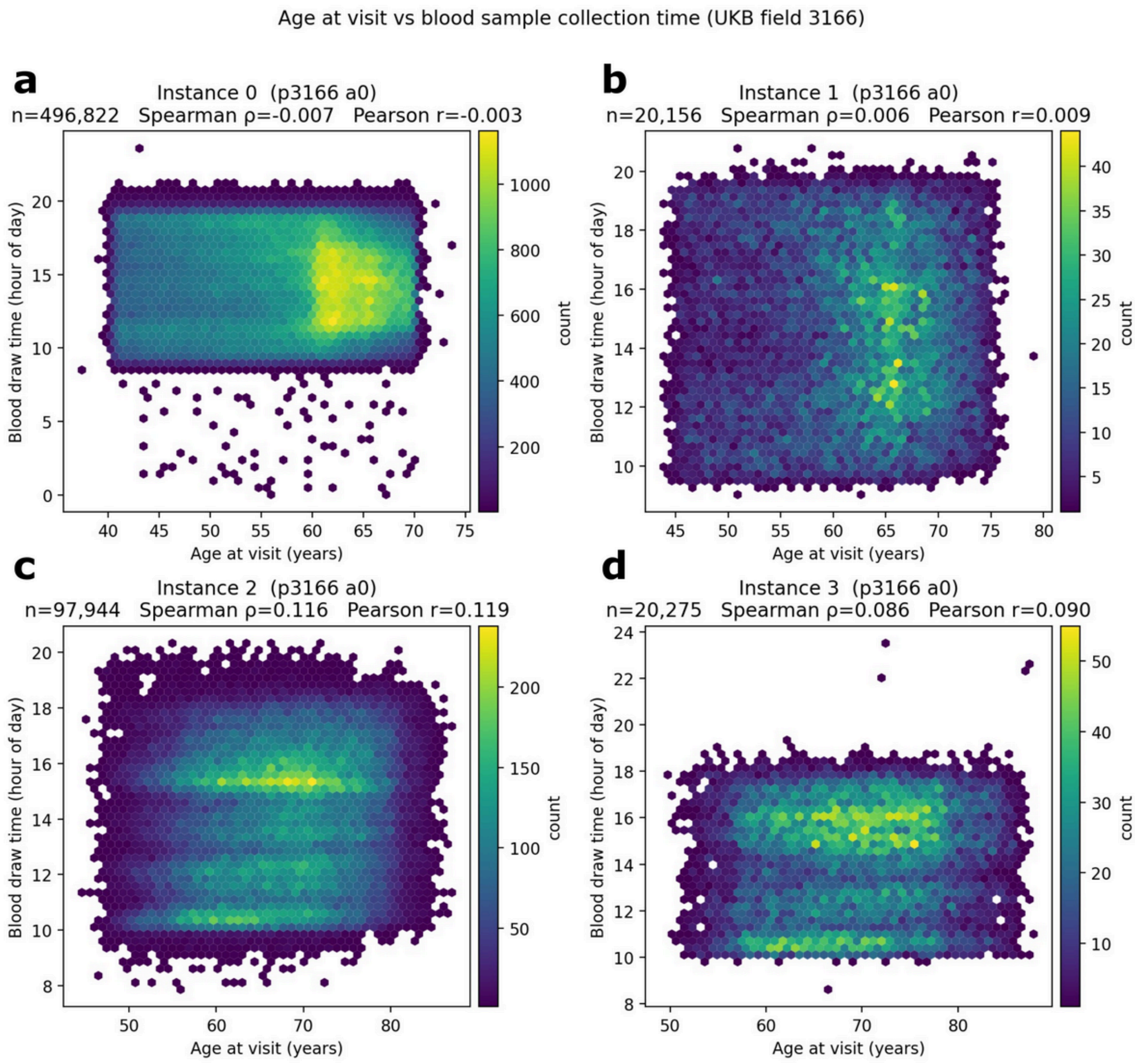


**Supplementary Fig. 9.** Relationship between participant age and blood sample collection time across UKB instances. (a–d) Hexbin density plots of blood draw time (UKB field 3166, hour of day) versus age at visit for instance 0 (a; n=496,822, Spearman ρ=-0.007, Pearson r=-0.003), instance 1 (b; n=20,156, ρ=0.006, r=0.009), instance 2 (c; n=97,944, ρ=0.116, r=0.119), and instance 3 (d; n=20,275, ρ=0.086, r=0.090).

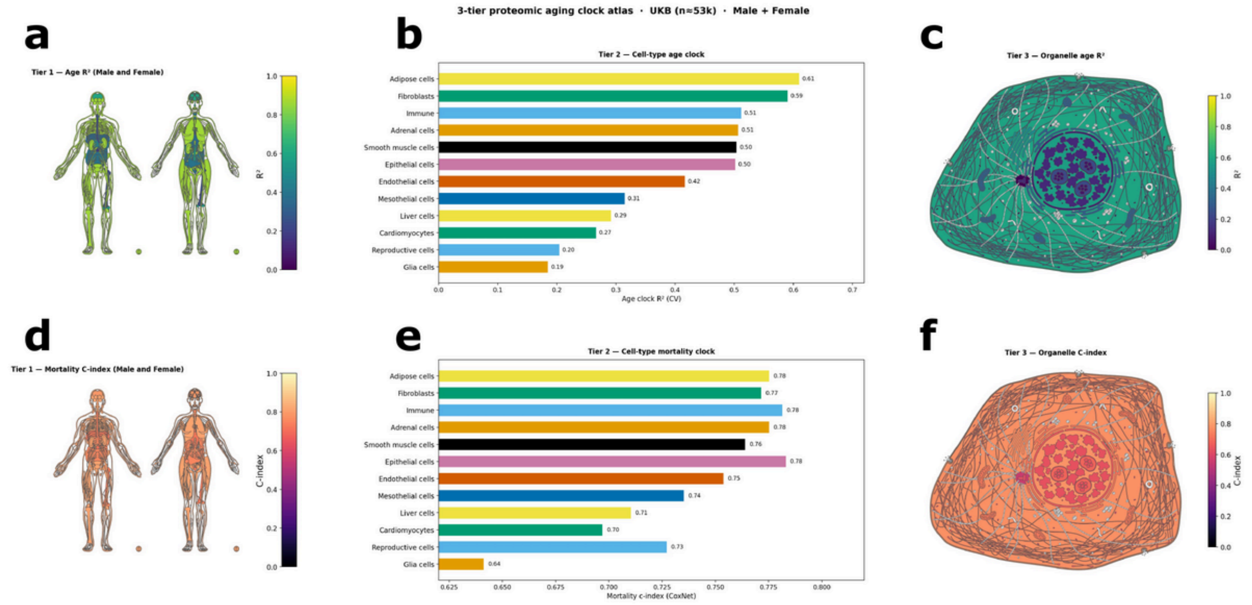


**Supplementary Fig. 10.** Three-tier proteomic aging and mortality clock atlas across the body (UKB Olink, n≈53k, male + female). (a) Tier 1 — whole-body anatomical mapping of age-clock $R^2$ across tissues. (b) Tier 2 — cell-type age clock CV-$R^2$ (12 grouped cell types: adipose, fibroblasts, immune, adrenal, smooth muscle, epithelial, endothelial, mesothelial, liver, cardiomyocytes, reproductive, glia). (c) Tier 3 — organelle-level age clock $R^2$ overlaid on a stylized cell schematic. (d) Tier 1 — whole-body anatomical mapping of mortality C-index (CoxNet) across tissues. (e) Tier 2 — cell-type mortality clock C-index across the same 12 grouped cell types. (f) Tier 3 — organelle-level mortality C-index overlaid on a stylized cell schematic.

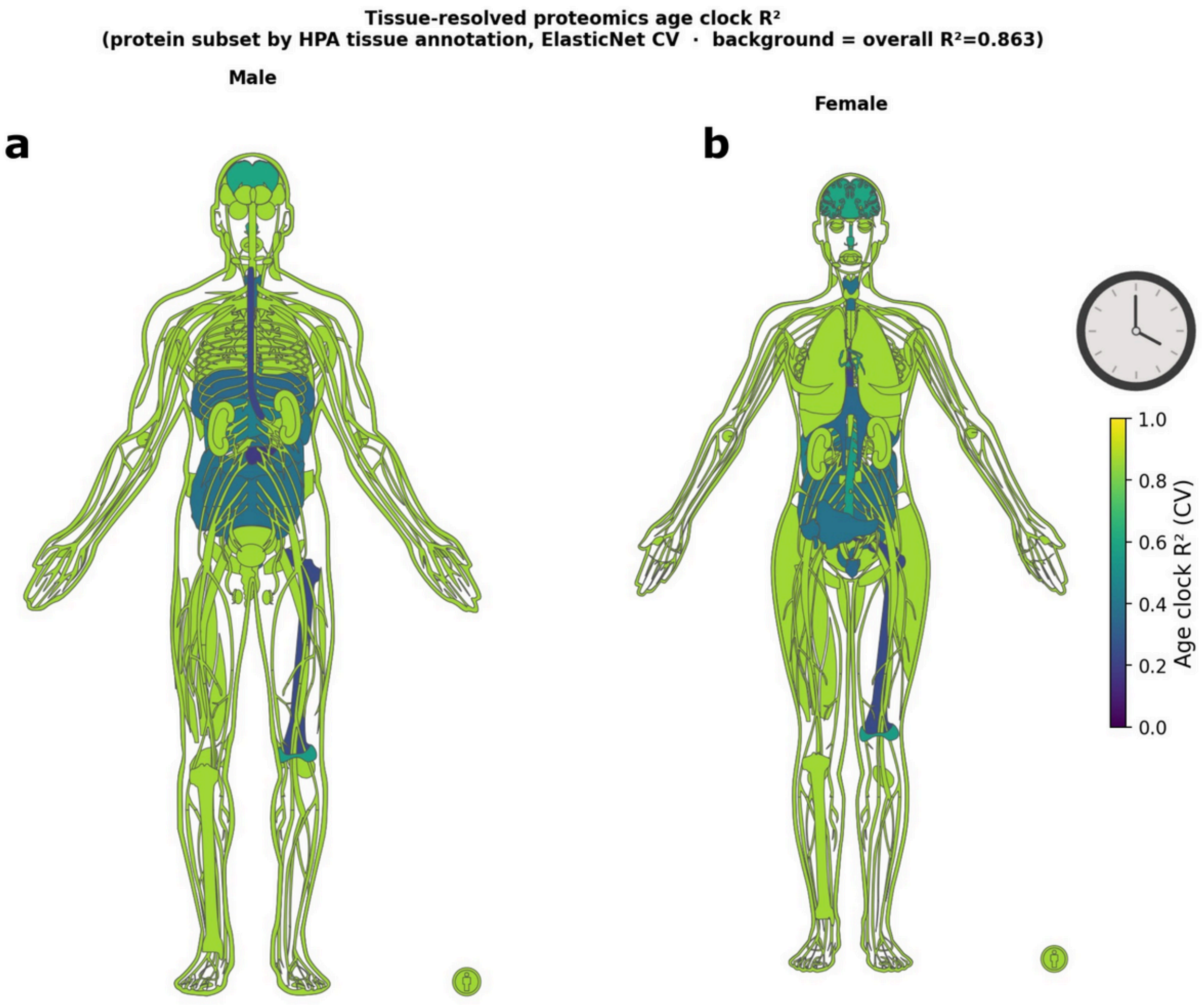


| Tissue | $R^2$ (CV) | Tissue | $R^2$ (CV) |
|---|---|---|---|
| Brain | 0.600 | Skin 1 | 0.261 |
| Blood Vessel | 0.561 | Heart Muscle | 0.244 |
| Liver | 0.439 | Bone Marrow | 0.234 |
| Lymphoid Tissue | 0.390 | Esophagus | 0.224 |
| Intestine | 0.383 | Placenta | 0.213 |
| Skeletal Muscle | 0.345 | Testis | 0.183 |
| Adipose Tissue | 0.297 | Pancreas | 0.178 |

**Supplementary Fig. 11.** Tissue-resolved proteomic age clock $R^2$ rendered on male and female anatomical templates (protein subset by HPA tissue annotation; ElasticNet CV; background = overall $R^2$=0.863). (a) Male whole-body map. (b) Female whole-body map. Tissues are colored by tissue-specific clock $R^2$ (CV). The accompanying table lists per-tissue $R^2$ values, ranging from brain (0.600) and blood vessel (0.561) at the top to pancreas (0.178) and testis (0.183) at the bottom.

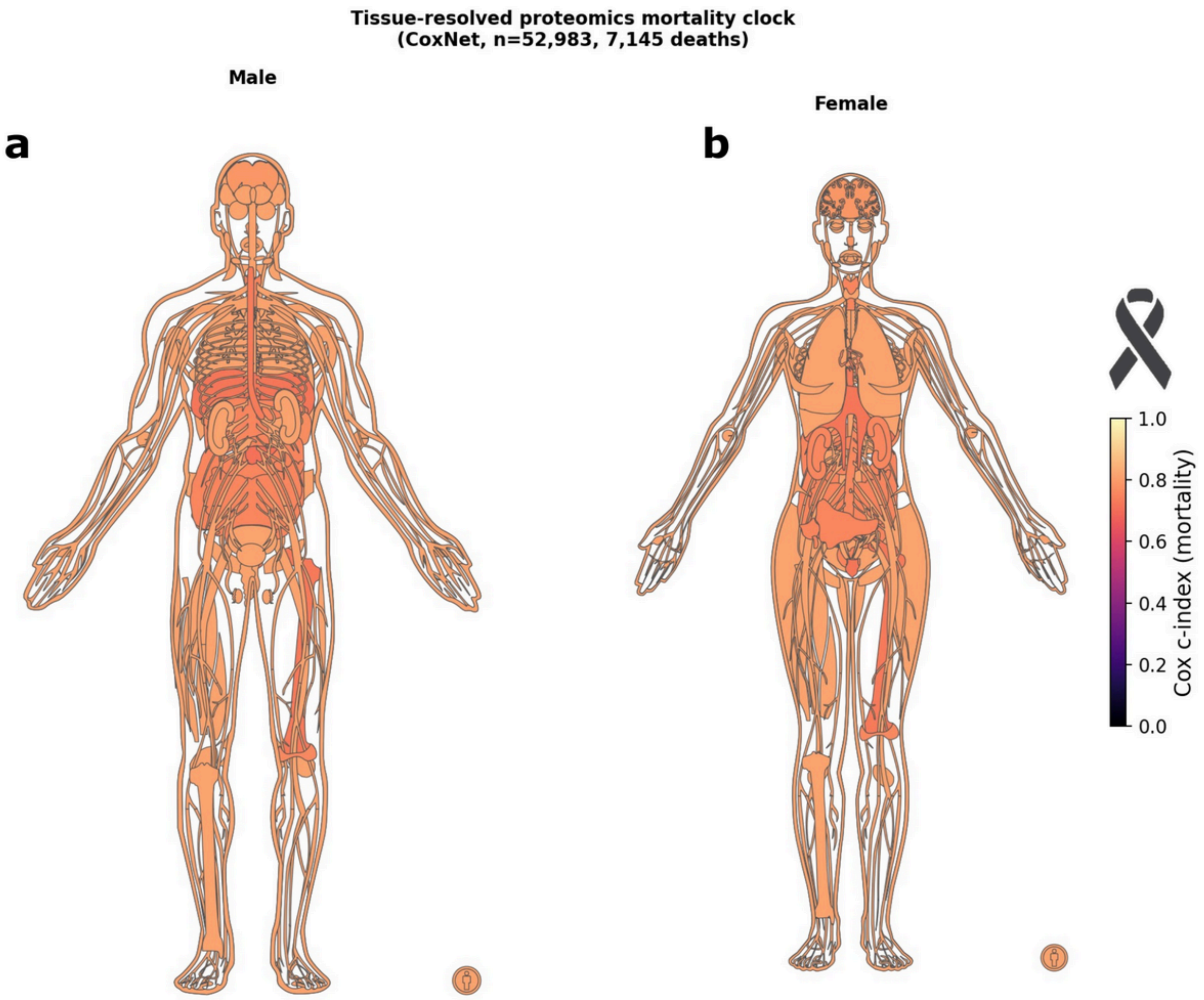


| Tissue | C-index | Tissue | C-index |
|---|---|---|---|
| Brain | 0.787 | Lung | 0.728 |
| Lymphoid Tissue | 0.766 | Skeletal Muscle | 0.723 |
| Blood Vessel | 0.758 | Parathyroid Gland | 0.720 |
| Kidney | 0.756 | Heart Muscle | 0.712 |
| Liver | 0.750 | Pancreas | 0.710 |
| Intestine | 0.750 | Skin 1 | 0.707 |
| Placenta | 0.736 | Choroid Plexus | 0.698 |
| Adipose Tissue | 0.733 | Testis | 0.650 |
| Esophagus | 0.730 | Retina | 0.608 |
| Bone Marrow | 0.729 | | |

**Supplementary Fig. 12.** Tissue-resolved proteomic mortality clock C-index rendered on male and female anatomical templates (CoxNet, n=52,983, 7,145 deaths). (a) Male whole-body map. (b) Female whole-body map. Tissues are colored by tissue-specific Cox C-index. The accompanying table lists per-tissue C-index values, with brain (0.787), lymphoid tissue (0.766), and blood vessel (0.758) at the top and retina (0.608) and testis (0.650) at the bottom.

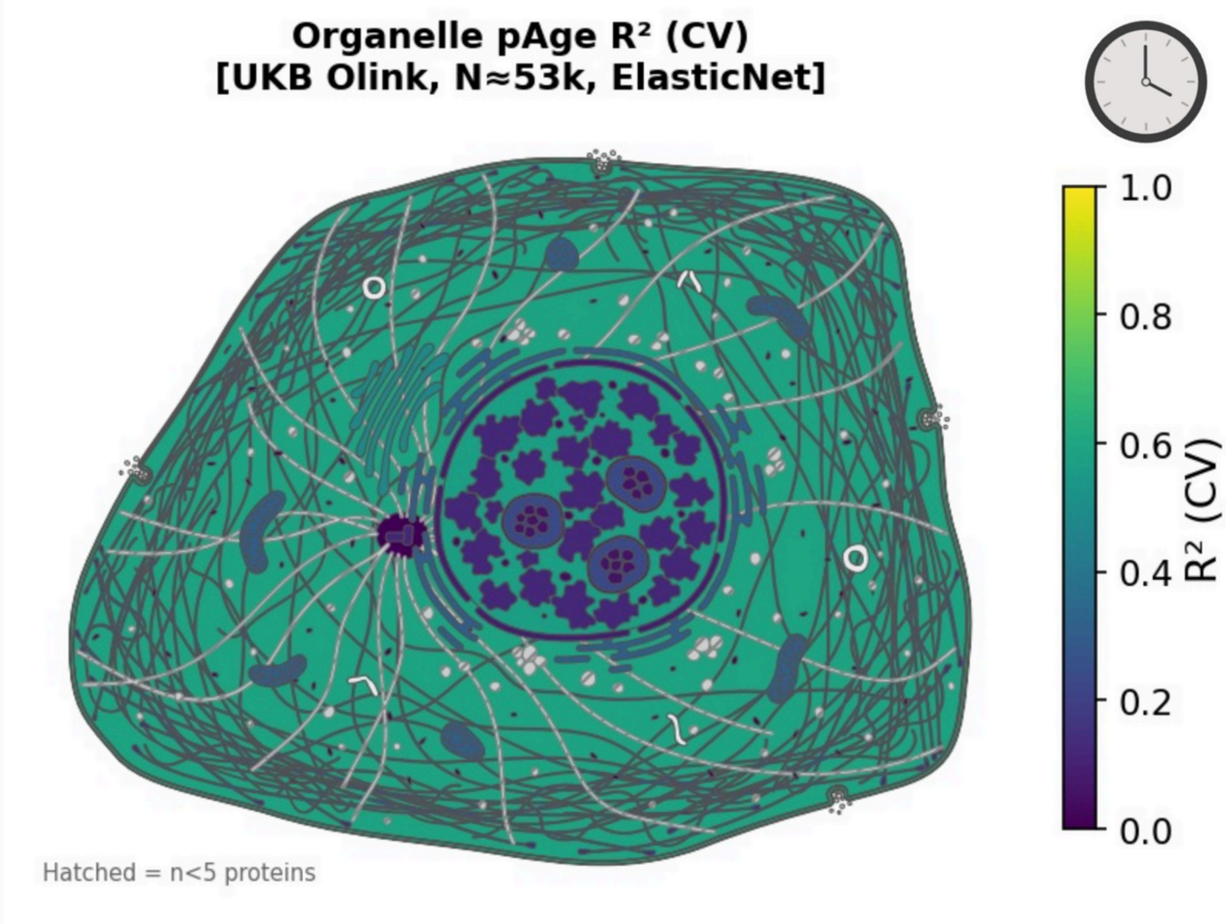


**Supplementary Fig. 13.** Organelle-level proteomic age clock $R^2$ rendered on a stylized cell schematic (UKB Olink, N≈53k, ElasticNet CV). Subcellular compartments are colored by their compartment-specific age-clock $R^2$ (CV). Compartments backed by fewer than five panel proteins are shown with hatched fill.

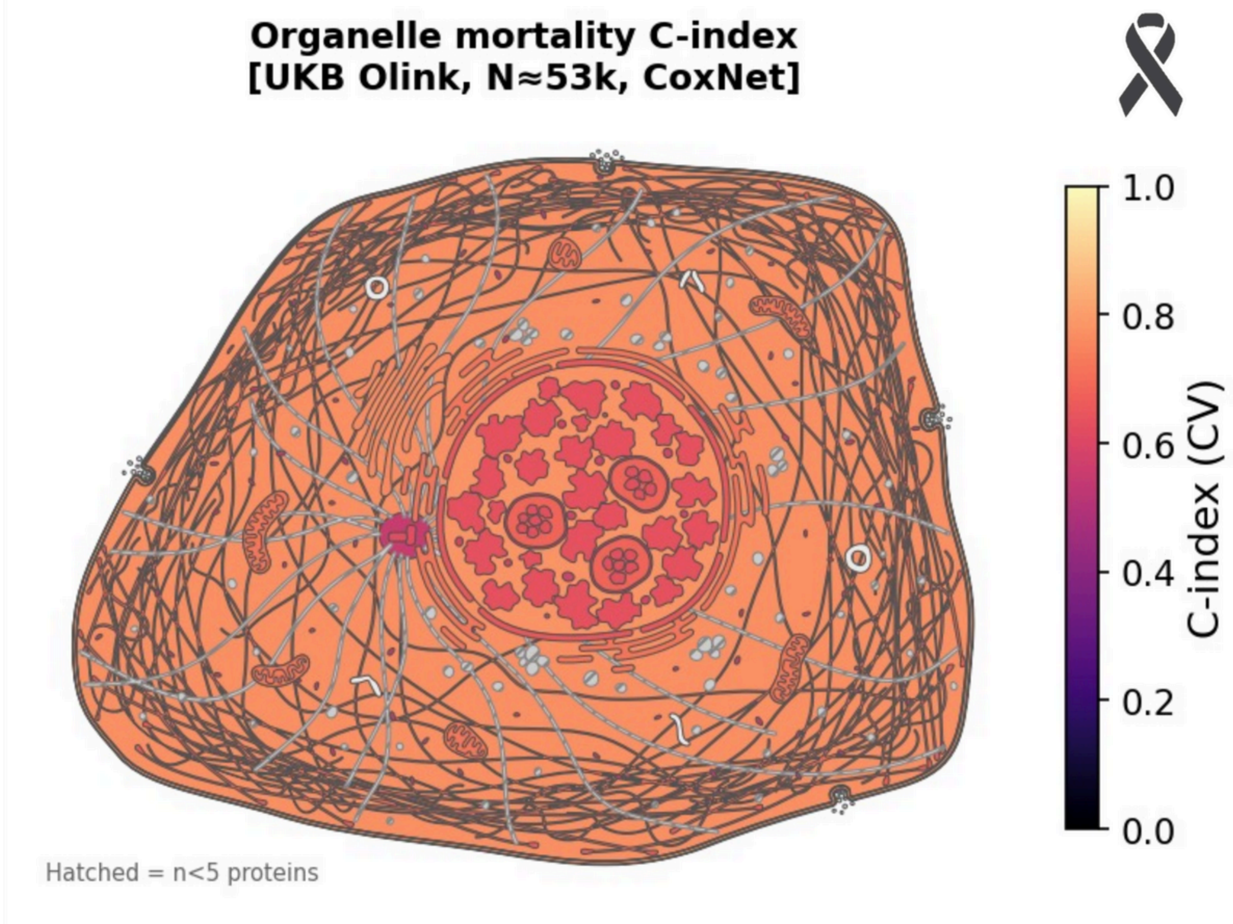


**Supplementary Fig. 14.** Organelle-level proteomic mortality clock C-index rendered on a stylized cell schematic (UKB Olink, N≈53k, CoxNet). Subcellular compartments are colored by their compartment-specific Cox C-index (CV). Compartments backed by fewer than five panel proteins are shown with hatched fill.

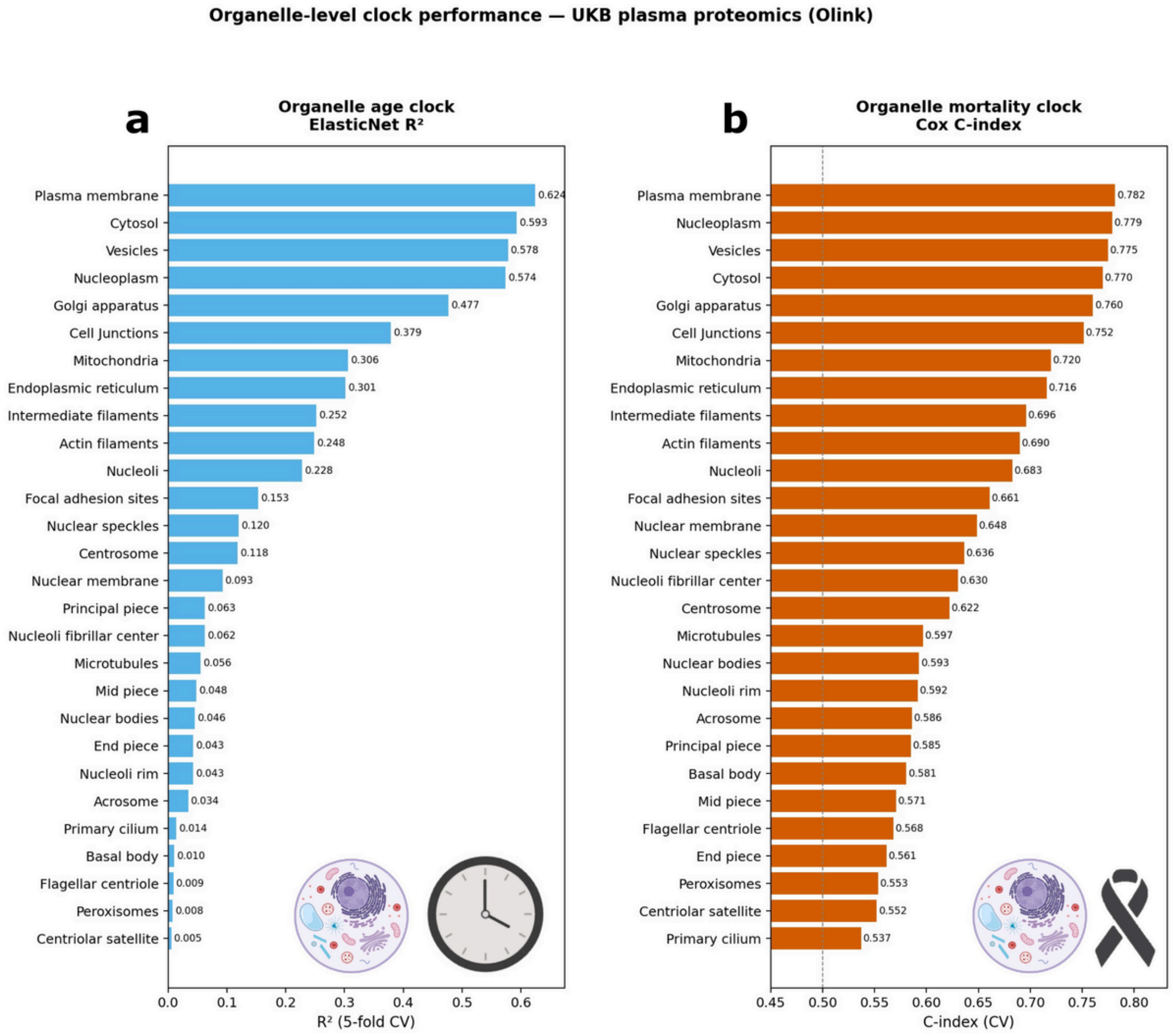


**Supplementary Fig. 15.** Organelle-level clock performance ranked across subcellular compartments (UKB plasma proteomics, Olink). (a) Organelle age clock ElasticNet $R^2$ (5-fold CV), led by plasma membrane (0.624), cytosol (0.593), vesicles (0.578), nucleoplasm (0.574), and Golgi apparatus (0.477). (b) Organelle mortality clock Cox C-index (CV), led by plasma membrane (0.782), nucleoplasm (0.779), vesicles (0.775), cytosol (0.770), and Golgi apparatus (0.760); dashed line at C=0.5 indicates chance performance.

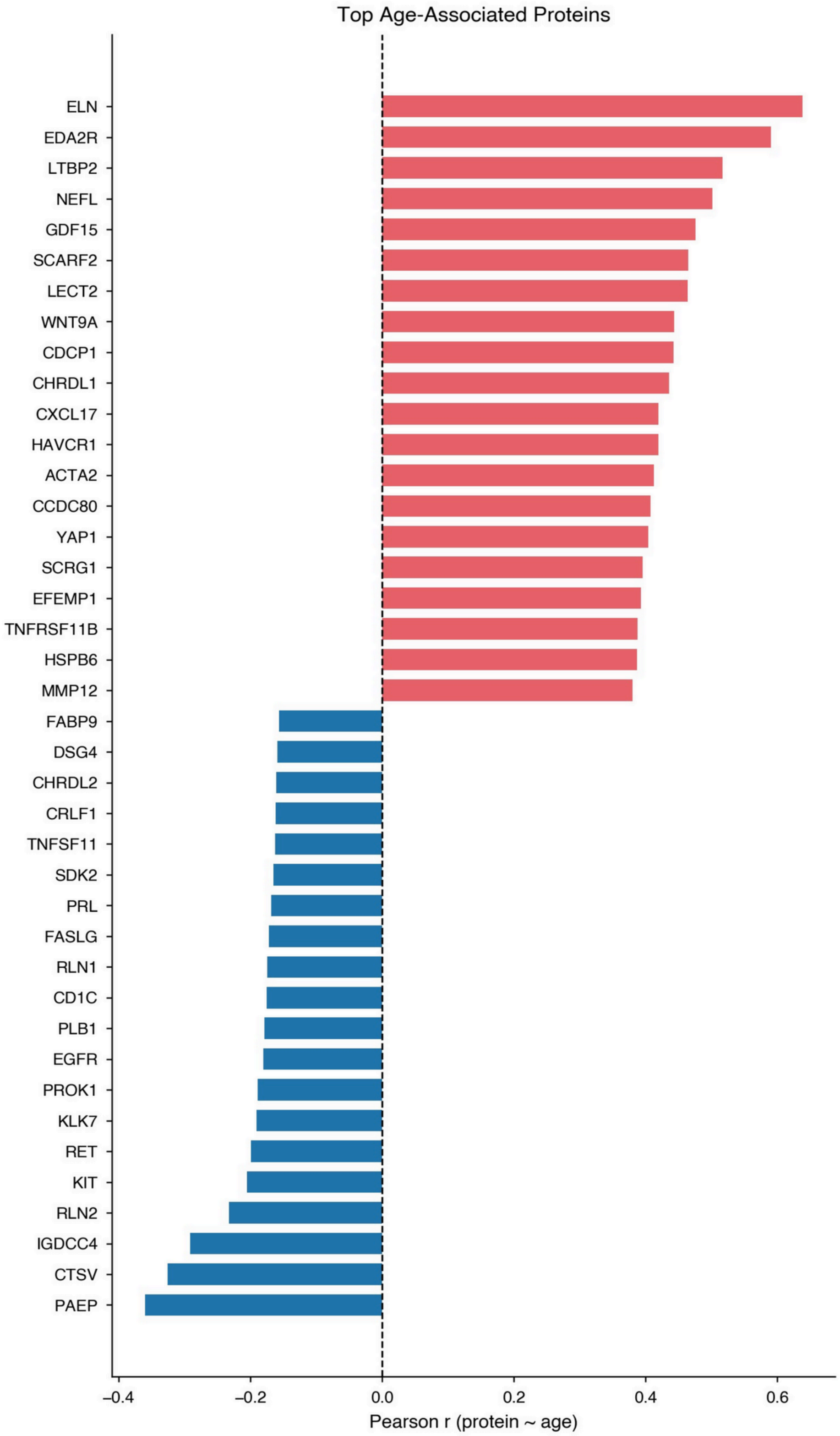


**Supplementary Fig. 16.** Top age-associated plasma proteins in the UKB Olink panel. Ranked Pearson correlation between protein level and chronological age, showing the 20 most positively correlated proteins (red, top, led by ELN, EDA2R, LTBP2, NEFL, GDF15) and the 20 most negatively correlated proteins (blue, bottom, led by PAEP, CTSV, IGDCC4, RLN2, KIT).

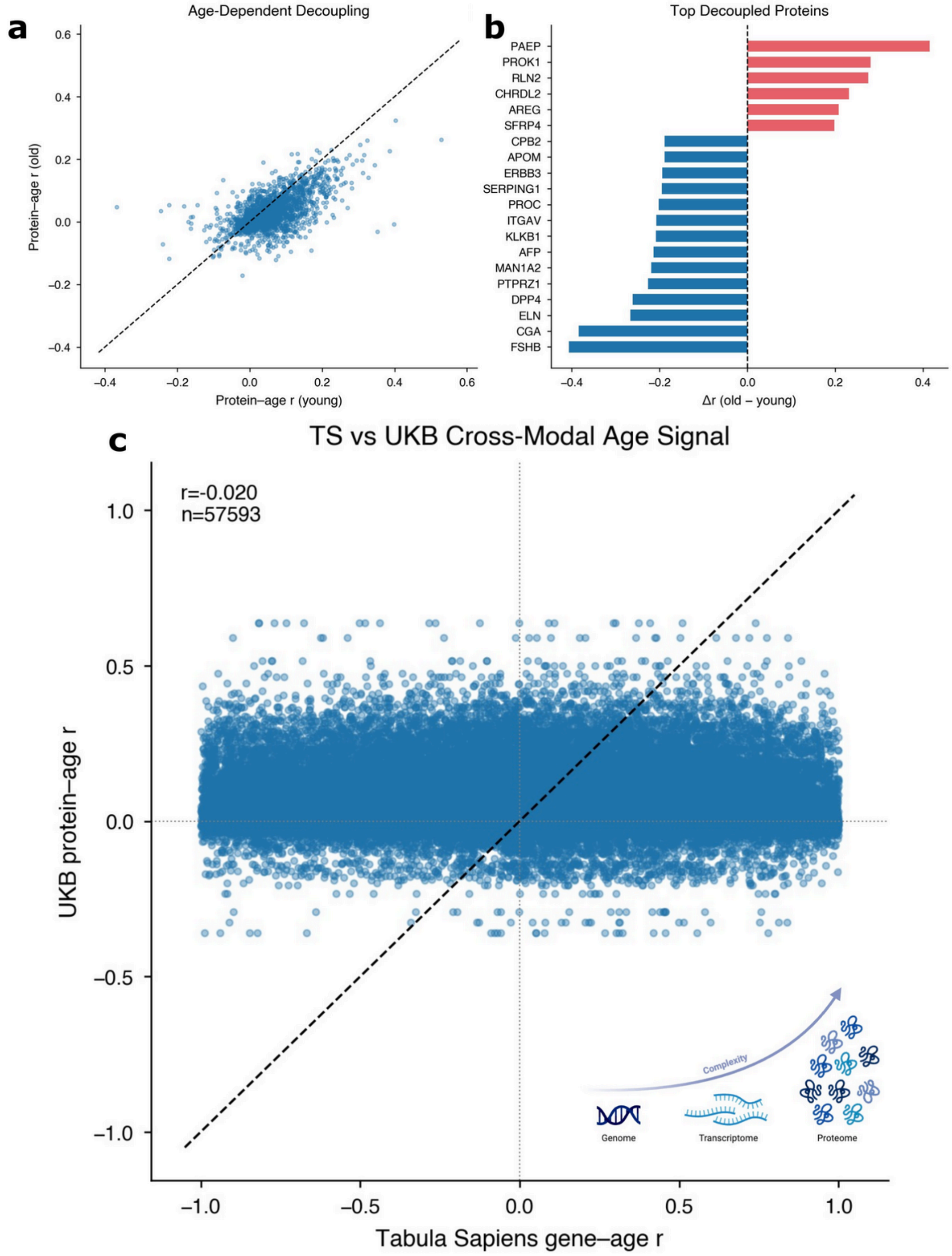


**Supplementary Fig. 17.** Age-dependent decoupling of plasma protein–age associations and cross-modal comparison with single-cell transcriptomics. (a) Per-protein Pearson correlation with age computed in younger (x-axis) vs. older (y-axis) UKB participants; dashed diagonal indicates equal correlation across age strata. (b) Top decoupled proteins ranked by Δ r (old - young), with positive shifts (red, e.g. PAEP, PROK1, RLN2, CHRDL2) and negative shifts (blue, e.g. FSHB, CGA, ELN, DPP4) indicating proteins whose age relationship strengthens or weakens with chronological age. (c) Cross-modal scatter of UKB plasma protein–age correlations versus Tabula Sapiens (TS) gene–age correlations across matched gene–tissue pairs (Pearson r=-0.020, n=57,593), illustrating limited transfer of bulk transcriptomic age signal to plasma proteomic age signal.

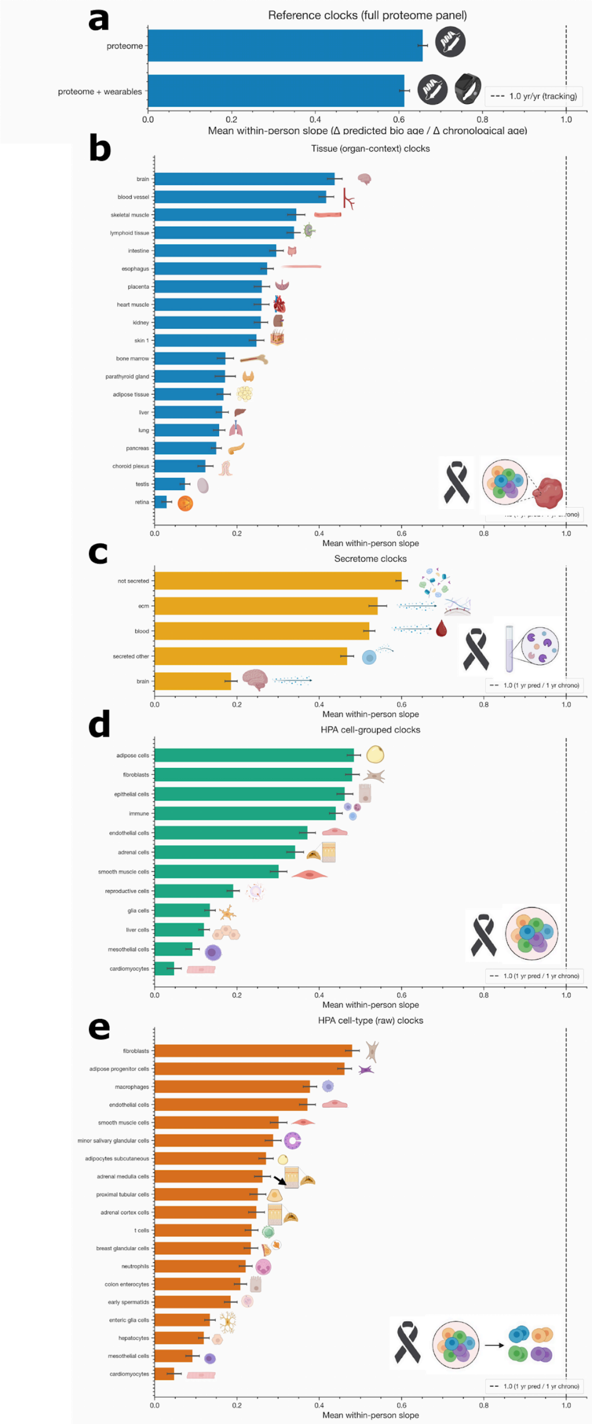


**Supplementary Fig. 18.** Mean within-person slopes (Δ predicted bio age / Δ chronological age) across all proteomic aging clock variants. (a) Reference clocks built on the full proteome panel (proteome-only and proteome + wearables). (b) Tissue (organ-context) clocks ranked across organs/tissues, led by brain, blood vessel, and skeletal muscle, and trailing in retina and testis. (c) Secretome clocks (not secreted,

ECM, blood, secreted other, brain). (d) HPA cell-grouped clocks across 12 grouped cell types, led by adipose cells, fibroblasts, and epithelial cells. (e) HPA cell-type (raw) clocks across 19 cell types, led by fibroblasts, adipose progenitor cells, and macrophages. Dashed line at 1.0 yr/yr indicates perfect longitudinal tracking.

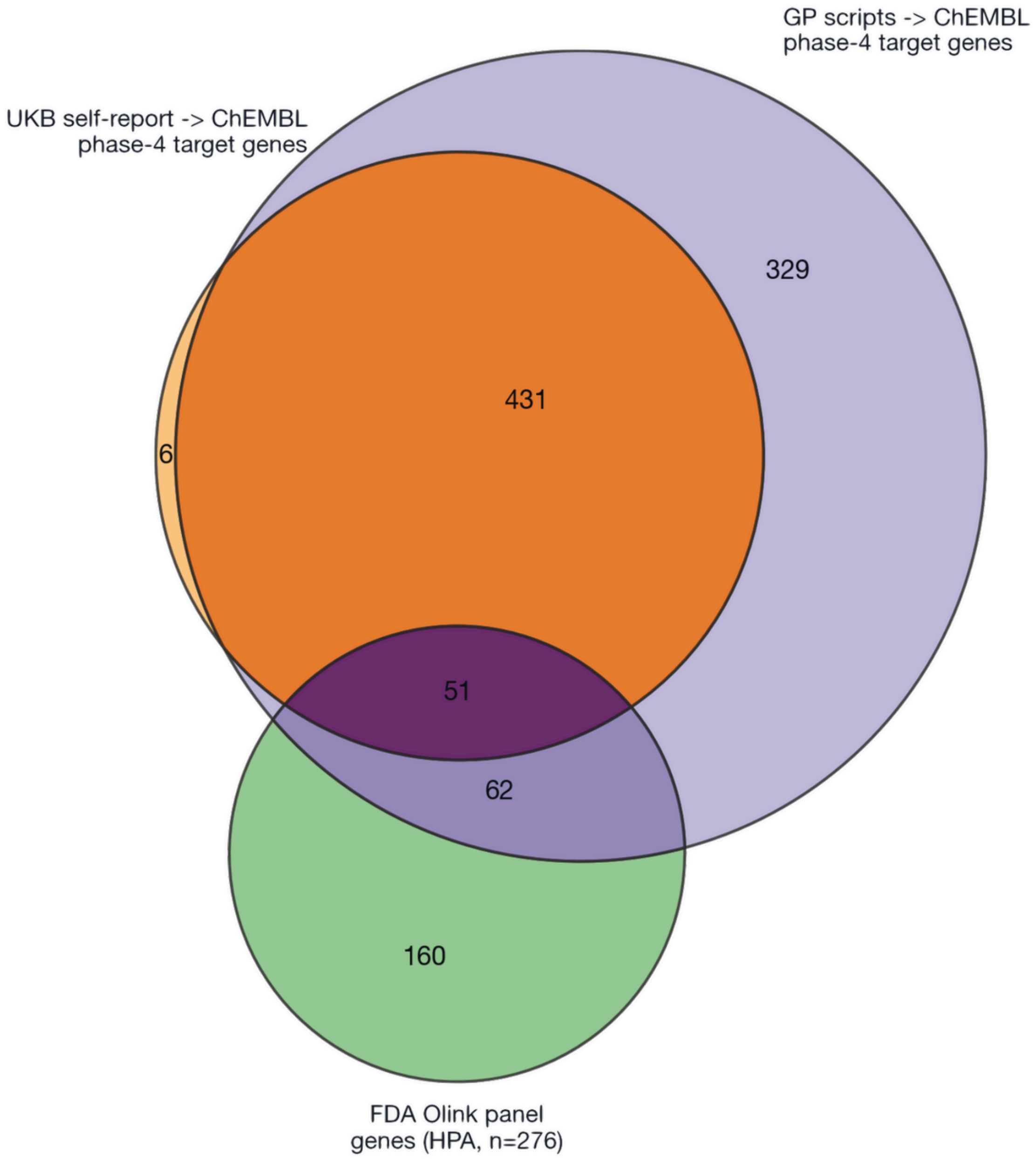


**Supplementary Fig. 19.** Mapping UKB-reported medications to FDA-approved Olink targets. Three-way Venn diagram of phase-4 drug-target genes derived from UKB self-reported medications (orange, mapped via ChEMBL), GP-script medications (purple, mapped via ChEMBL), and the FDA-flagged subset of the Olink panel from HPA (green, n=276). Overlaps: 431 genes shared between UKB self-report and GP scripts, 51 across all three sources, 62 between GP scripts and FDA-Olink, and 6 unique to UKB self-report; 329 GP-script-only and 160 FDA-Olink-only.

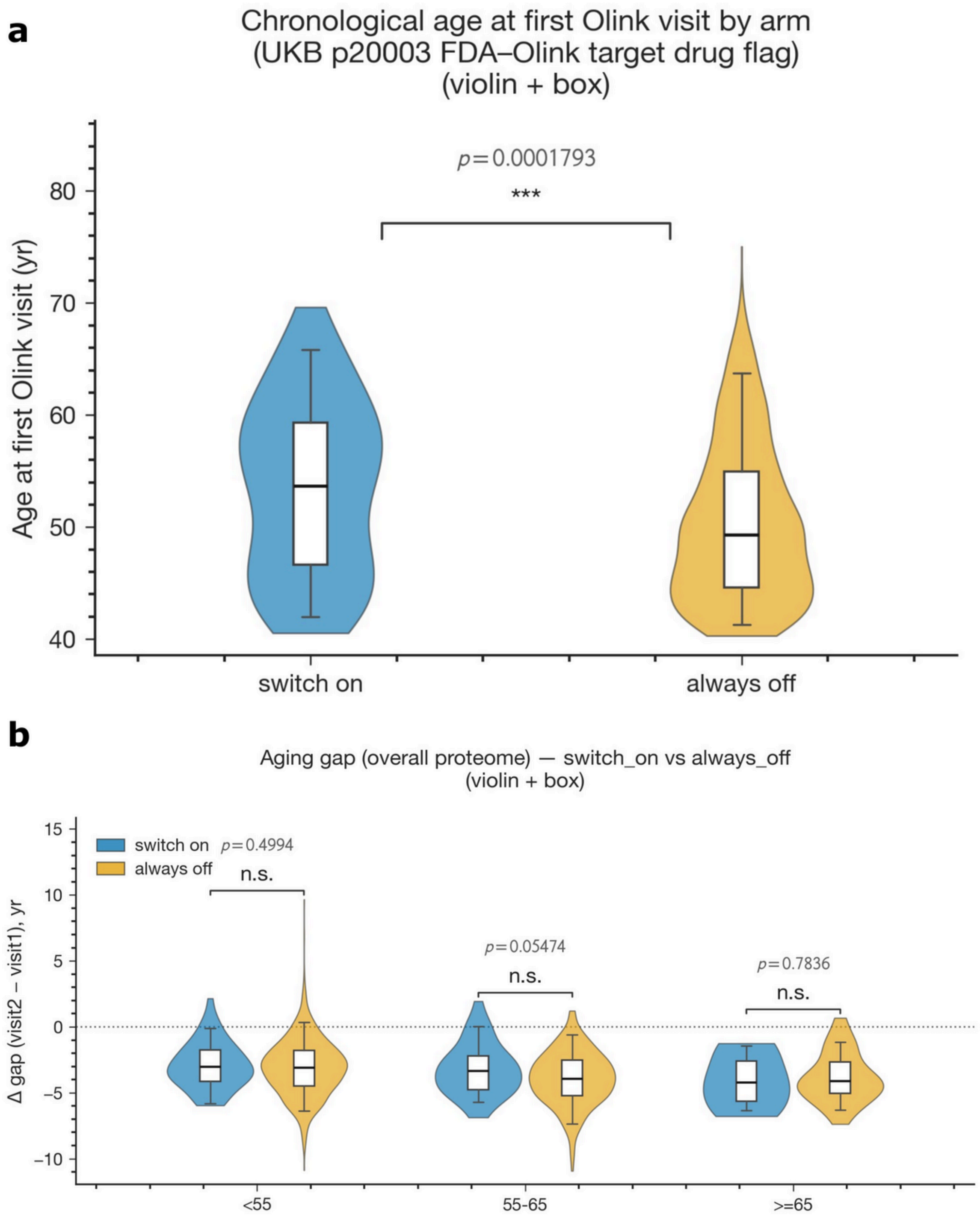


**Supplementary Fig. 20.** Baseline characteristics and longitudinal aging-gap differences by FDA-target drug exposure (overall proteome). (a) Chronological age at first Olink visit for switch-on (blue) vs. always-off (orange) participants based on the UKB p20003 FDA-Olink target drug flag (Welch $p=1.79\times10^{-4}$). (b) Δ age gap (visit 2 - visit 1) from the overall proteome clock, switch-on vs. always-off, stratified by age group (<55, 55–65, ≥65); none of the per-stratum comparisons reach significance.

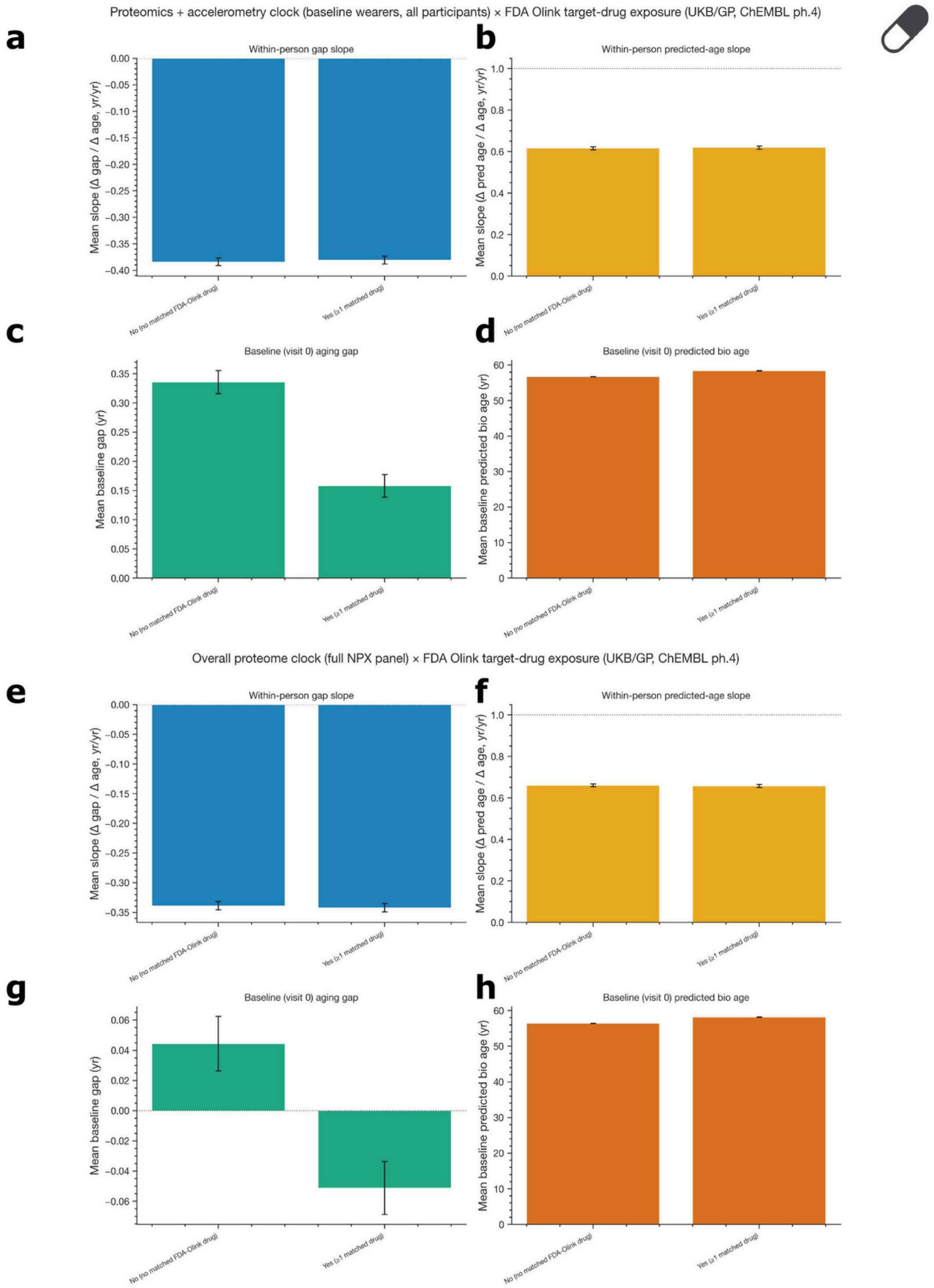


**Supplementary Fig. 21.** Lifestyle/exposure-stratified within-person slopes and baseline age gaps, FDA Olink target-drug exposure (UKB/GP, ChEMBL ph.4). (a–d) Proteomics + accelerometry clock (baseline wearers, all participants), comparing participants with no matched FDA-Olink drug vs. ≥1 matched drug: within-person Δgap slope (a), within-person predicted-age slope (b), baseline (visit 0) age gap (c), and baseline predicted bio age (d). (e–h) Overall proteome clock (full NPX panel), with the same four readouts under the same drug exposure stratification.

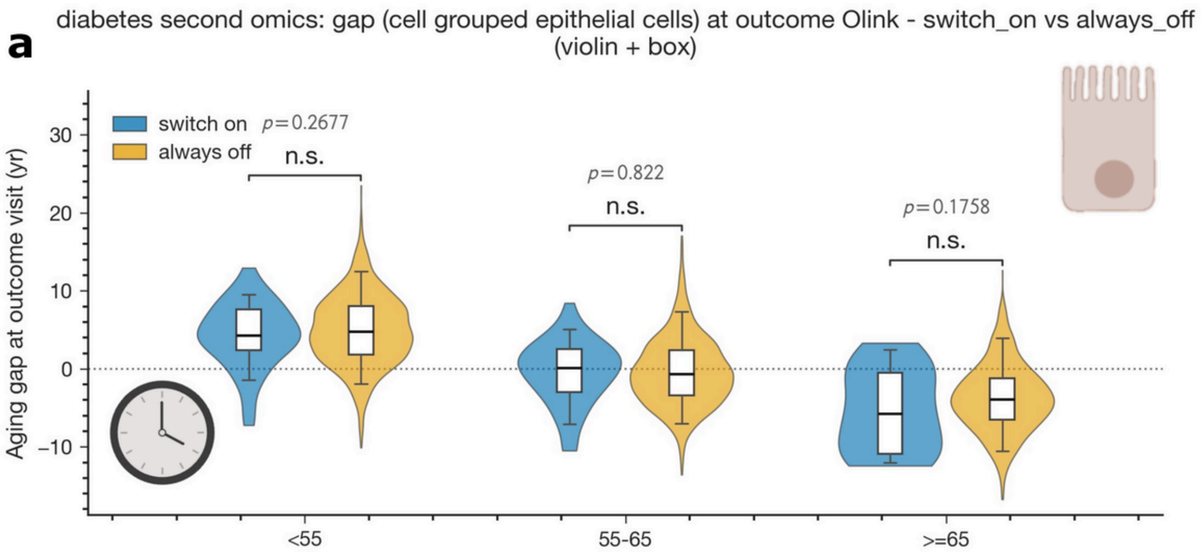


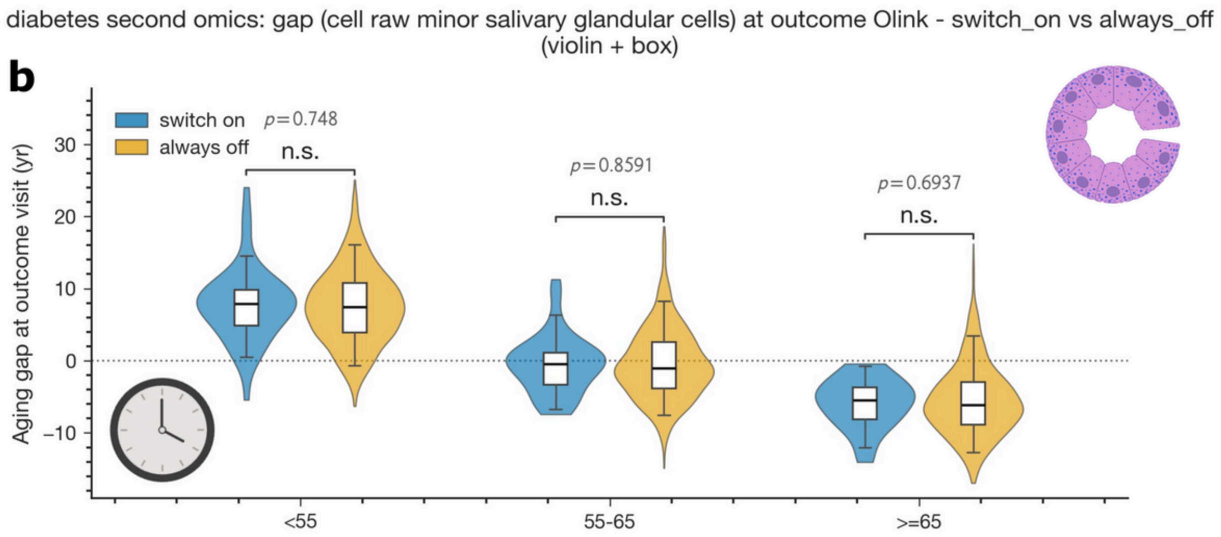


**Supplementary Fig. 22.** Diabetic-cohort second-omics aging-gap analysis using cell-specific clocks: switch-on vs. always-off at outcome visit, stratified by age (<55, 55–65, ≥65). (a) Age gap from the cell-grouped epithelial-cell clock at the outcome Olink visit; no significant differences across age strata. (b) Age gap from the raw minor salivary glandular cell clock at the outcome Olink visit; no significant differences across age strata.

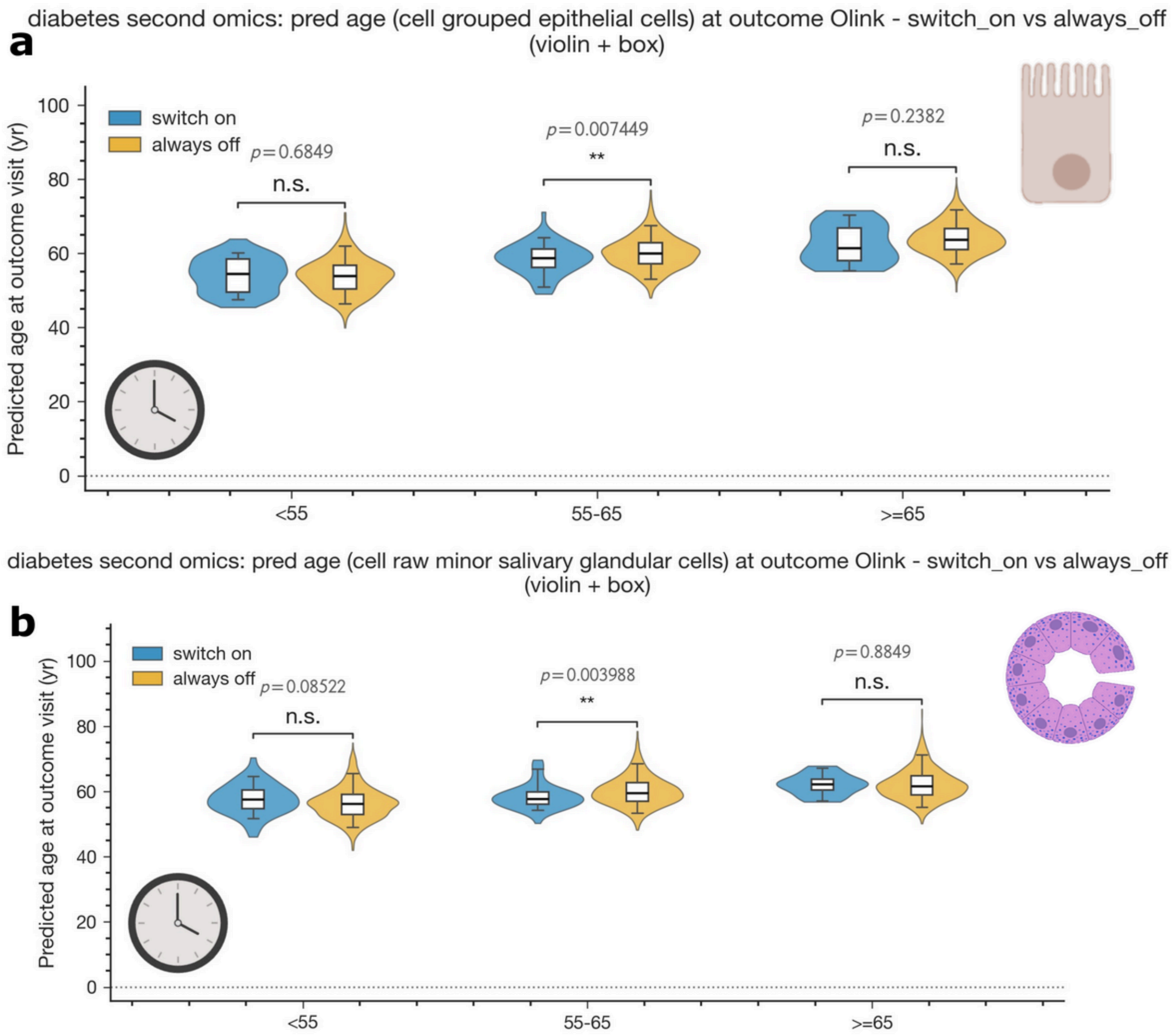


**Supplementary Fig. 23.** Diabetic-cohort second-omics predicted-age analysis using cell-specific clocks: switch-on vs. always-off at outcome visit, stratified by age (<55, 55–65, ≥65). (a) Predicted age from the cell-grouped epithelial-cell clock; switch-on participants show significantly lower predicted age in the 55–65 stratum (p=0.0074). (b) Predicted age from the raw minor salivary glandular cell clock; switch-on participants show significantly lower predicted age in the 55–65 stratum (p=0.0040).

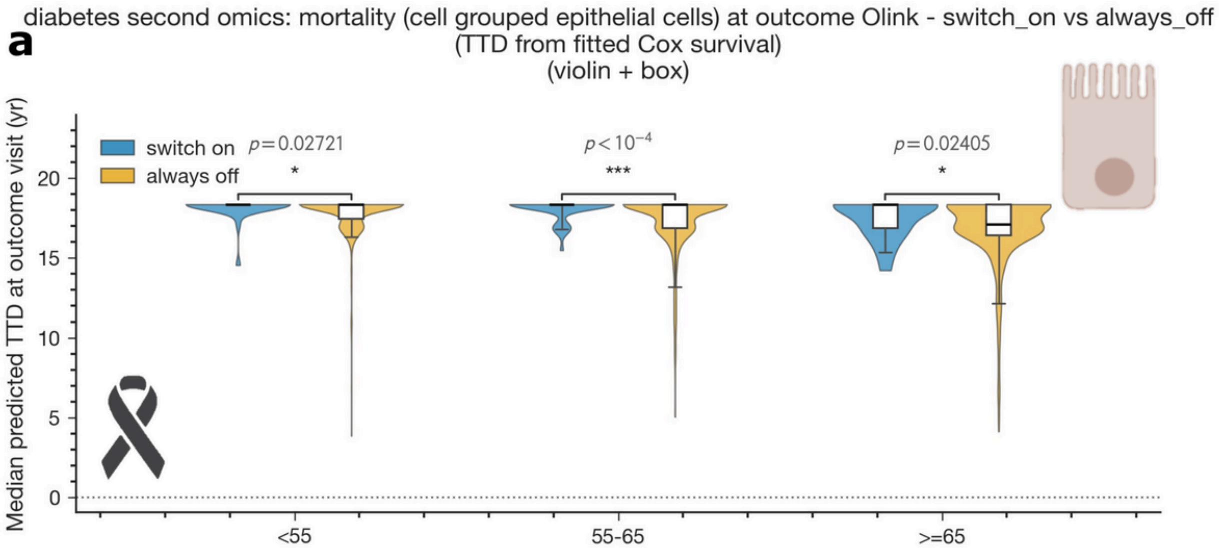


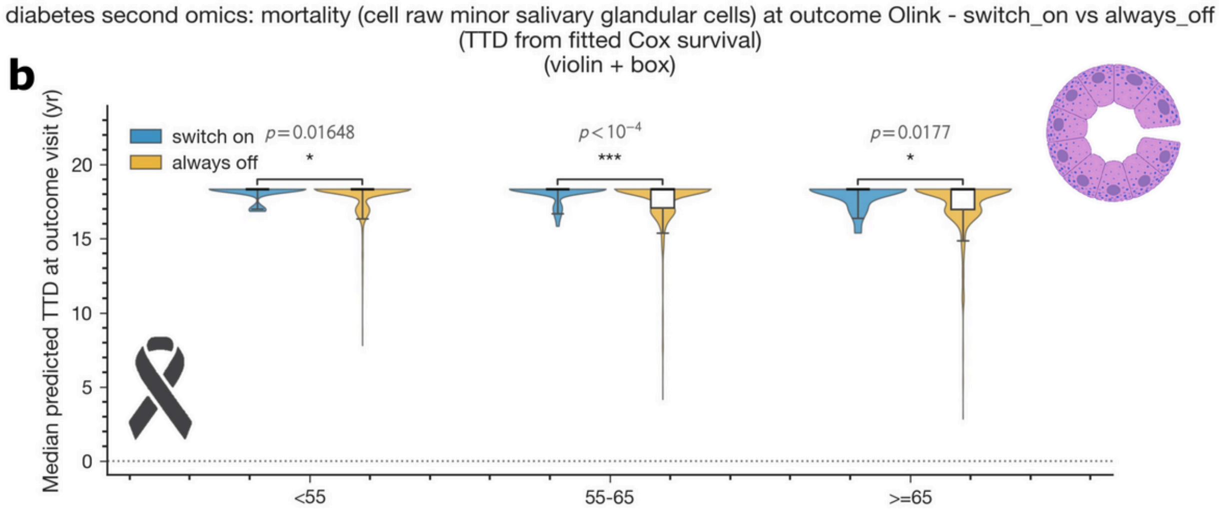


**Supplementary Fig. 24.** Diabetic-cohort second-omics mortality analysis using cell-specific clocks: median predicted time-to-death (TTD from fitted Cox survival) at outcome visit, switch-on vs. always-off, stratified by age (<55, 55–65, ≥65). (a) Cell-grouped epithelial-cell clock; switch-on participants show significantly higher predicted TTD across all three age strata ($p=0.027$, $p<10^{-4}$, $p=0.024$). (b) Raw minor salivary glandular cell clock; switch-on participants show significantly higher predicted TTD across all three age strata ($p=0.016$, $p<10^{-4}$, $p=0.018$).

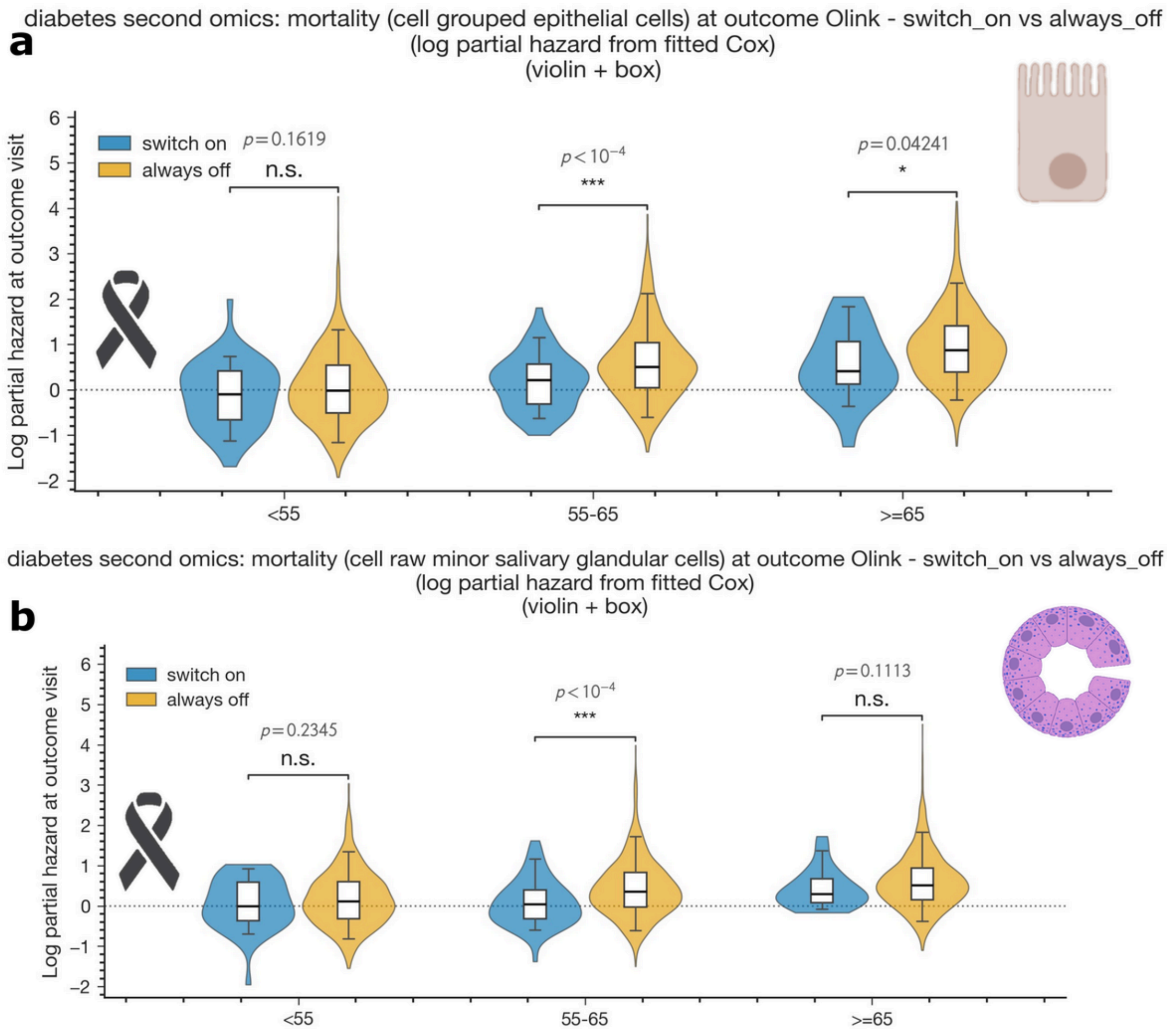


**Supplementary Fig. 25.** Diabetic-cohort second-omics mortality analysis using cell-specific clocks: log partial hazard (from fitted Cox) at outcome visit, switch-on vs. always-off, stratified by age (<55, 55–65, ≥65). (a) Cell-grouped epithelial-cell clock; switch-on participants show significantly lower log partial hazard in the 55–65 ($p<10^{-4}$) and ≥65 (p=0.042) strata. (b) Raw minor salivary glandular cell clock; switch-on participants show significantly lower log partial hazard in the 55–65 stratum ($p<10^{-4}$).

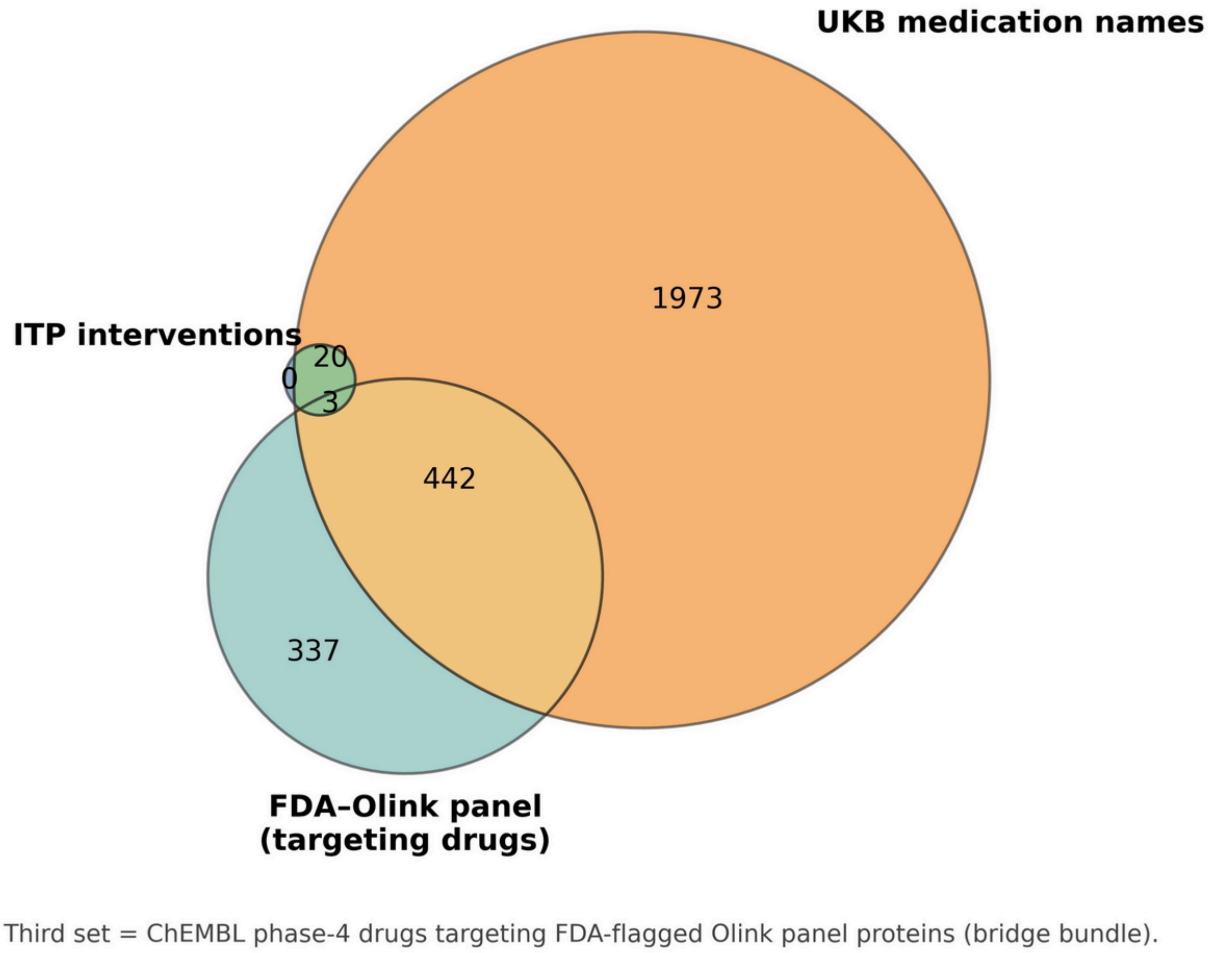


**Supplementary Fig. 26.** Overlap between NIA ITP interventions, UKB medications, and FDA-flagged Olink-panel targeting drugs. Three-way Venn diagram of NIA ITP intervention drugs (green), UKB self-reported/GP-script medication names (orange), and ChEMBL phase-4 drugs targeting FDA-flagged Olink panel proteins (teal, the “bridge bundle”). 442 drugs are shared between UKB medications and the FDA-Olink targeting set, 3 are shared between ITP and the FDA-Olink targeting set, and 20 ITP interventions overlap with UKB medication names but not the FDA-Olink set. UKB-only (n=1,973) and FDA-Olink-targeting-only (n=337) regions are also shown.

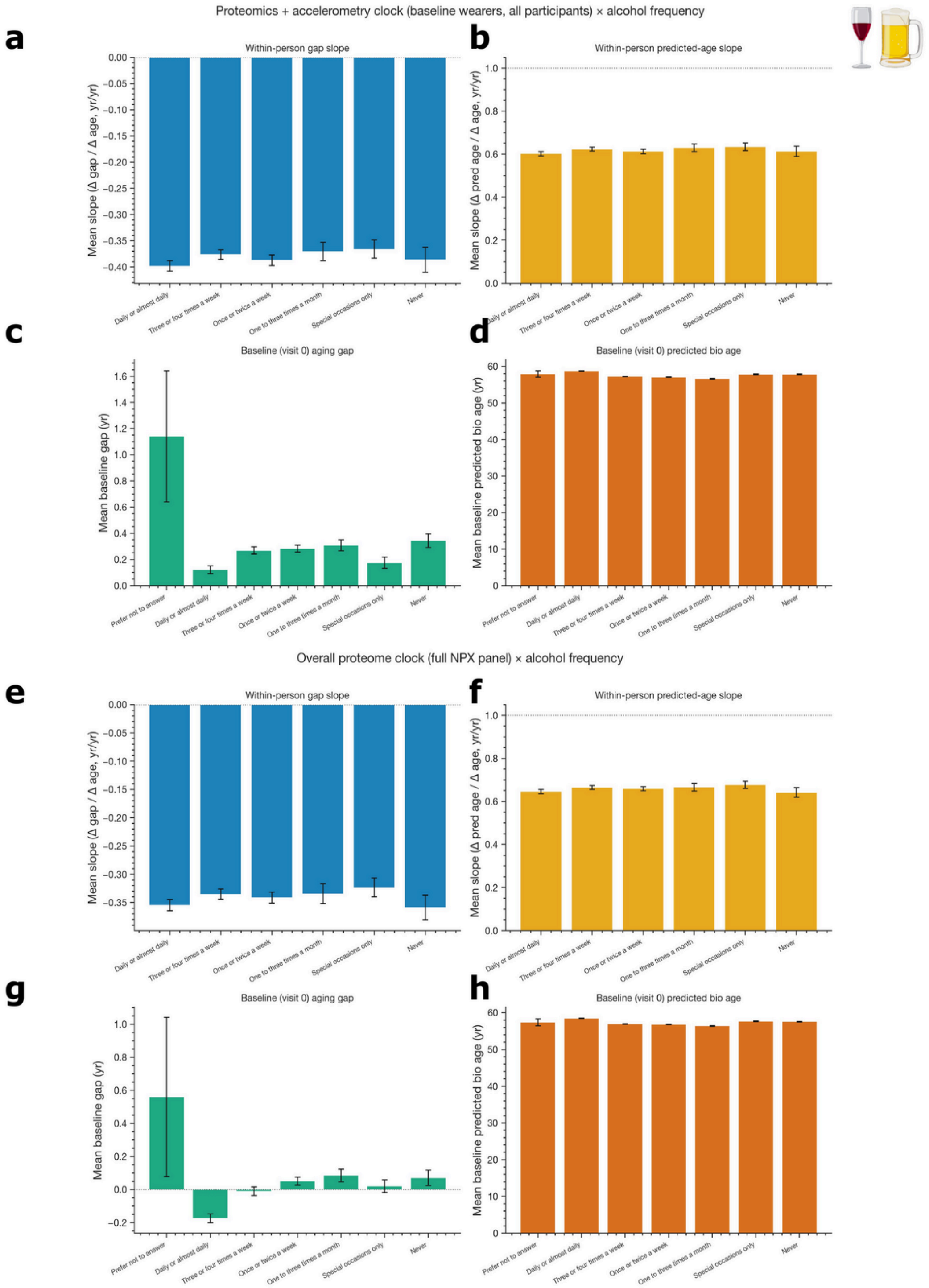


**Supplementary Fig. 27.** Within-person slopes and baseline age gaps stratified by alcohol frequency. (a–d) Proteomics + accelerometry clock (baseline wearers, all participants) by alcohol frequency category (daily or almost daily, three or four times a week, once or twice a week, one to three times a month, special occasions only, never): within-person Δgap slope (a), within-person predicted-age slope (b), baseline age gap (c), and baseline predicted bio age (d). (e–h) Overall proteome clock (full NPX panel) under the same alcohol-frequency stratification, with the additional "prefer not to answer" category included in the baseline panels.

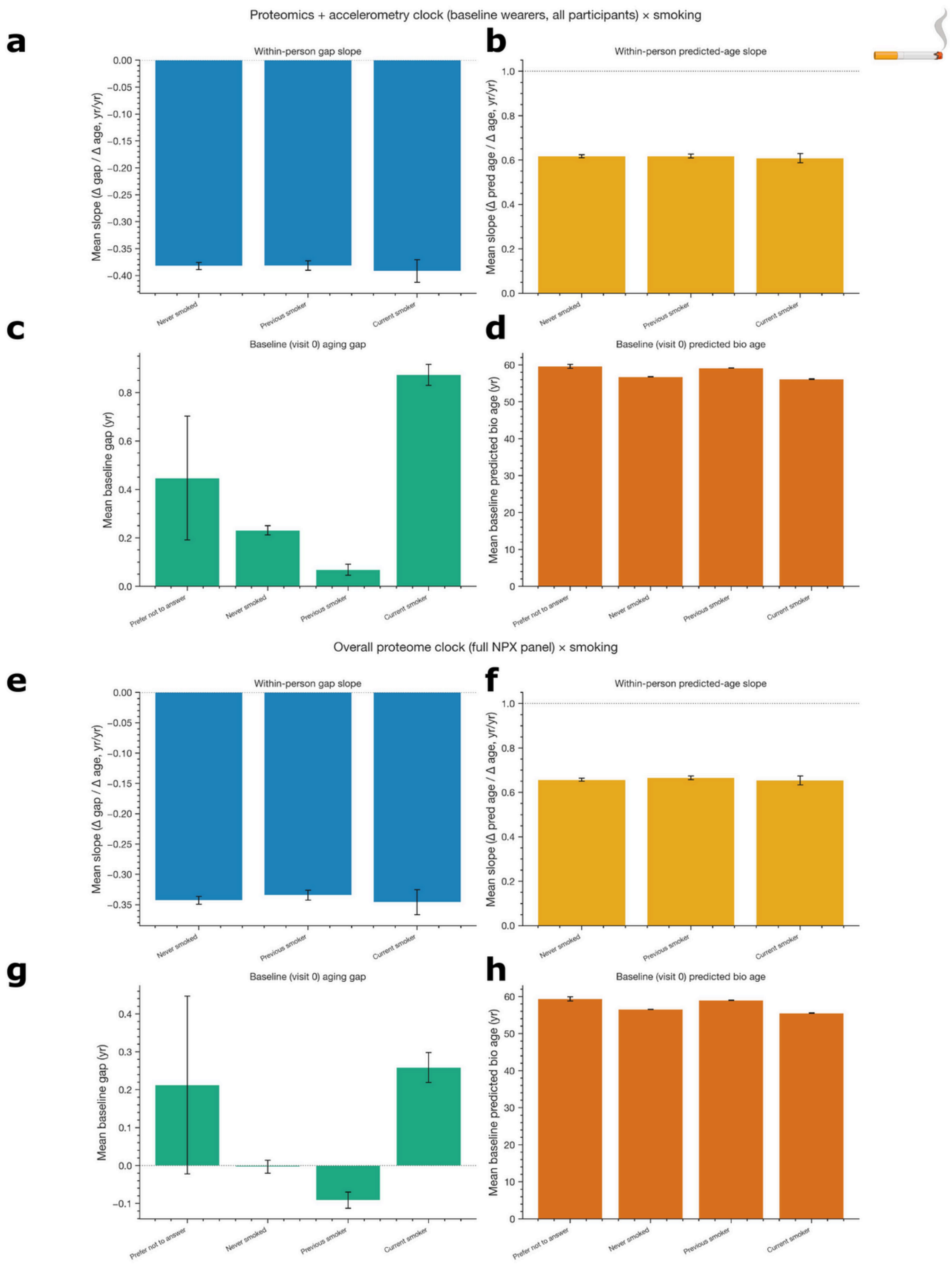


**Supplementary Fig. 28.** Within-person slopes and baseline age gaps stratified by smoking status. (a–d) Proteomics + accelerometry clock (baseline wearers, all participants) by smoking status (never smoked, previous smoker, current smoker): within-person Δgap slope (a), within-person predicted-age slope (b), baseline age gap (c), and baseline predicted bio age (d). Current smokers show the largest baseline age gap. (e–h) Overall proteome clock (full NPX panel) under the same smoking stratification, with "prefer not to answer" included in the baseline panels.

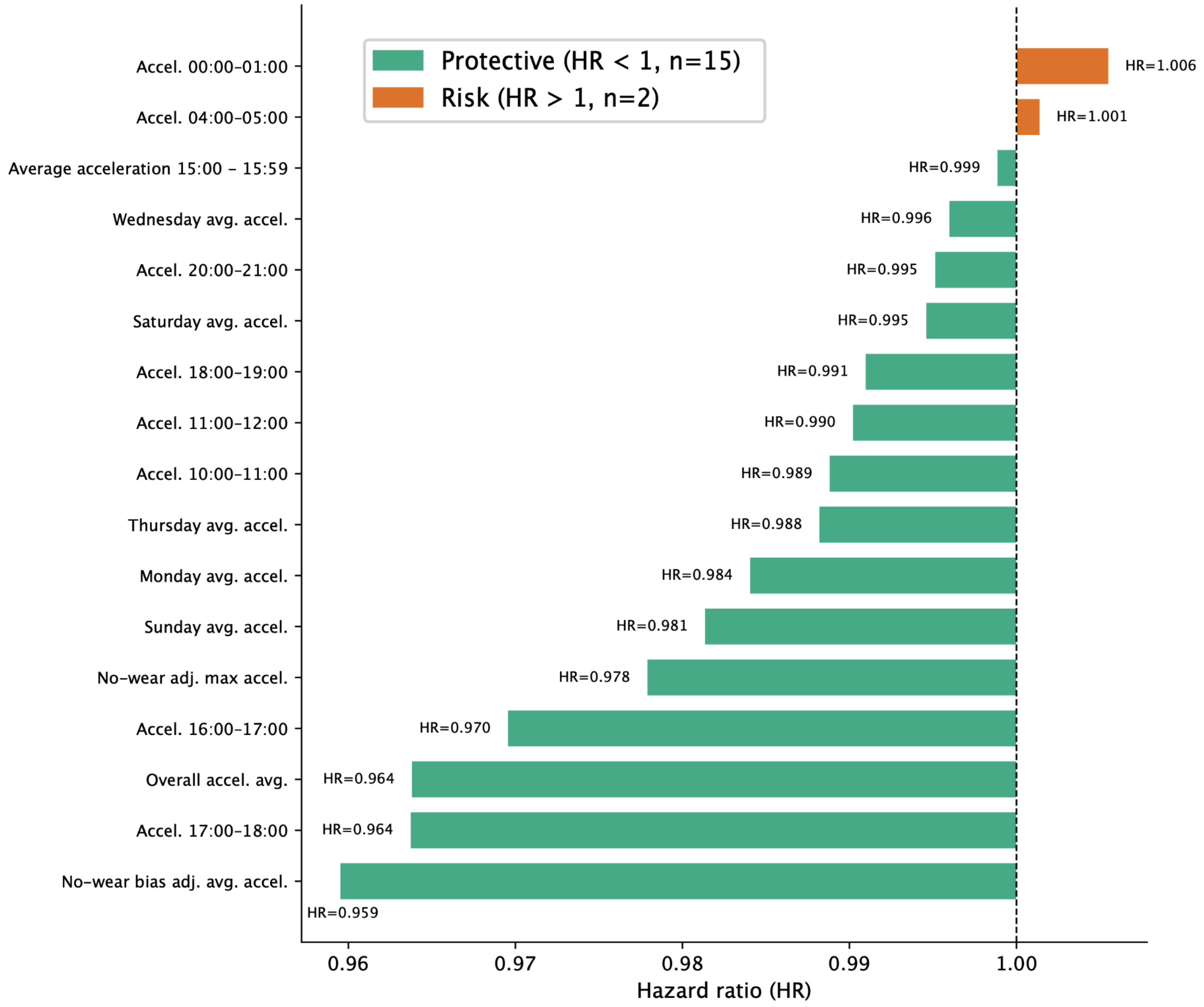


**Supplementary Fig. 29.** Wearable accelerometry features in the trimodal Cox mortality model, ranked by hazard ratio. Lollipop plot of all 17 wearable features retained by the multimodal (proteomics + wearables) Cox proportional-hazards mortality model, color-coded as protective (HR<1, n=15, green) or risk (HR>1, n=2, orange). The 15 protective wearable features—led by no-wear-bias-adjusted average acceleration (HR=0.959), afternoon acceleration 17:00–18:00 (HR=0.964), overall daily average acceleration (HR=0.964), and late-afternoon acceleration 16:00–17:00 (HR=0.970)—are dominated by daytime moderate-intensity activity signals consistent with prior accelerometer mortality studies[28–31]. Only two wearable features show HR>1: midnight acceleration 00:00–01:00 (HR=1.006) and early-morning acceleration 04:00–05:00 (HR=1.001), both far below the protein-level top-25 risk-feature threshold (HR1.024) and likely reflecting nocturnal activity patterns associated with sleep disruption rather than independent biological risk. The dashed vertical line at HR=1 marks the neutral risk reference. This figure complements Fig. 1e by showing the full wearable-feature hazard-ratio spectrum that is otherwise truncated by the protein-dominated top-25 ranking.

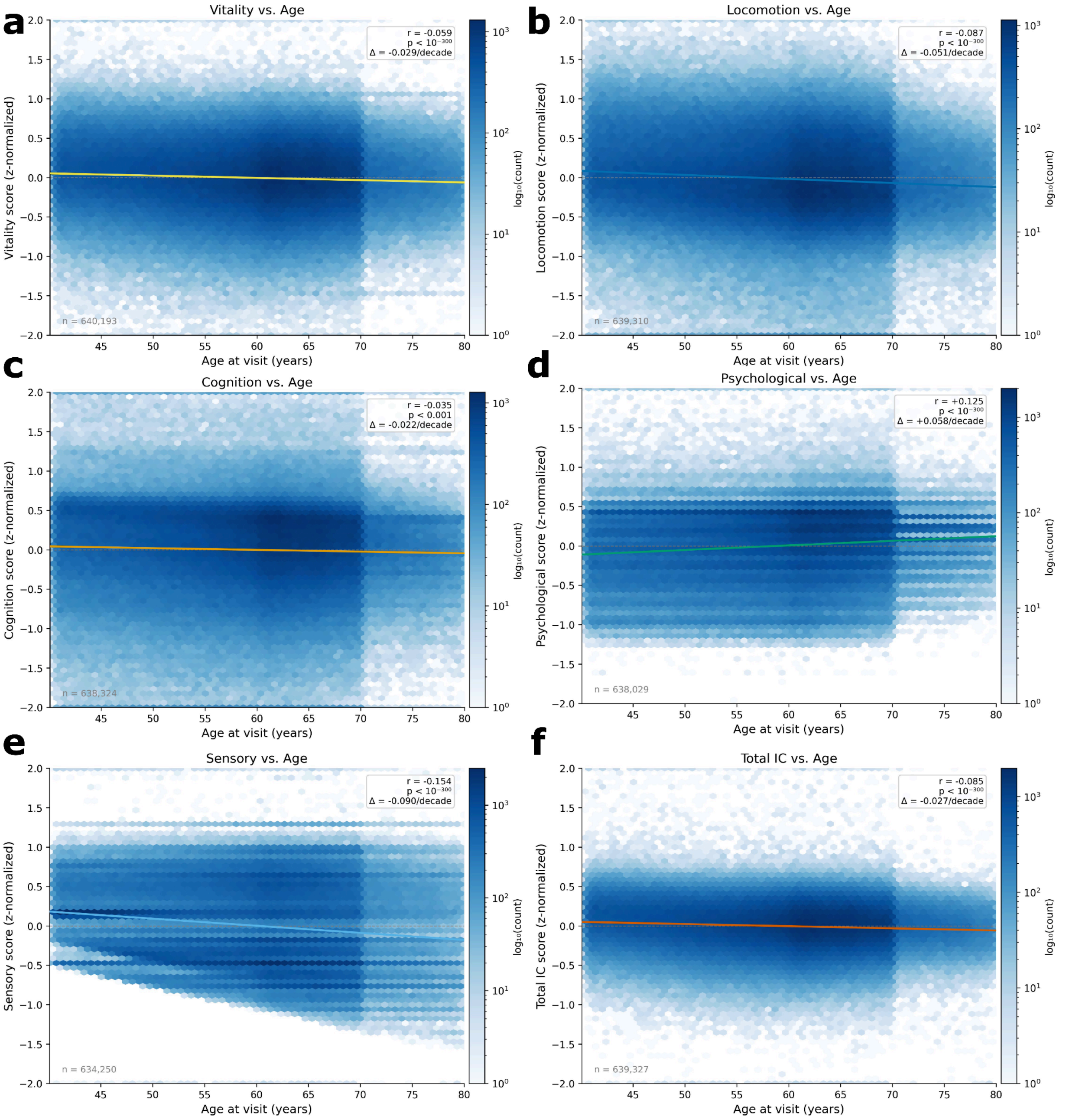


**Supplementary Fig. 30.** Intrinsic capacity domain scores across age in UK Biobank participants, visualized as Hexbin density plots showing z-normalized intrinsic capacity (IC) domain scores as a function of age at visit for (a) vitality, (b) locomotion, (c) cognition, (d) psychological well-being, (e) sensory function, and (f) total intrinsic capacity. Color intensity represents log-scaled participant density (log(count)). Solid lines indicate linear regression fits across age, with corresponding Pearson correlation coefficients (r), p-values, and per-decade score changes (Δ/decade) shown in each panel. Most IC domains exhibit modest declines with increasing age, particularly sensory and locomotion function, whereas psychological scores show a slight positive association with age. Sample sizes for each domain are indicated in the lower-left corner of each panel.

***Supplementary Tables***

**Table S1.** *Intrinsic capacity (IC) domain items used in this study. Each item is direction-harmonized so that higher values indicate better capacity, z-scored, and averaged within domain. The domain score is the mean of all available standardized items (≥1 item required). Items marked ↓ were negated prior to z-scoring (higher raw value = worse capacity).*

| Domain | UKB Field | Description | Dir. |
|---|---|---|---|
| ***Vitality (26 items)*** | | | |
| | p21001 | Body mass index (kg/m²) | ↓ |
| | p48 | Waist circumference (cm) | ↓ |
| | p49 | Hip circumference (cm) | ↓ |
| | p46 | Left-hand grip strength (kg) | ↑ |
| | p47 | Right-hand grip strength (kg) | ↑ |
| | p1160 | Self-reported sleep duration (h) | ↑ |
| | p90041 | Wearable-measured sleep duration (h) | ↑ |
| | p2100 | Frequency of sleeplessness/insomnia | ↓ |
| | p2110 | Frequency of daytime dozing | ↓ |
| | p20517 | Trouble falling or staying asleep | ↓ |
| | p2080 | Frequency of tiredness/lethargy | ↓ |
| | p20077 | Days of unusual fatigue | ↓ |
| | p29011 | Appetite change (poor appetite/overeating) | ↓ |
| | p30740 | Fasting plasma glucose (mmol/L) | ↓ |
| | p30750 | Glycated haemoglobin HbA1c (mmol/mol) | ↓ |
| | p30870 | Plasma triglycerides (mmol/L) | ↓ |
| | p30780 | LDL cholesterol (mmol/L) | ↓ |
| | p30760 | HDL cholesterol (mmol/L) | ↑ |
| | p30690 | Total cholesterol (mmol/L) | ↓ |
| | p30710 | C-reactive protein (mg/L) | ↓ |
| | p30660 | Serum albumin (g/L) | ↑ |
| | p30020 | Haemoglobin concentration (g/dL) | ↑ |
| | p30900 | 25-hydroxyvitamin D (nmol/L) | ↑ |

| Domain | UKB Field | Description | Dir. |
|---|---|---|---|
| | p30700 | Serum creatinine (µmol/L) | ↓ |
| | p30720 | Cystatin C (mg/L) | ↓ |
| | p30840 | Testosterone (nmol/L) | ↑ |
| ***Locomotion (20 items)*** | | | |
| | p864 | Days/week walking ≥10 min | ↑ |
| | p874 | Usual walking pace | ↓ |
| | p884 | Days/week moderate physical activity | ↑ |
| | p894 | Duration of moderate activity (min/day) | ↑ |
| | p904 | Days/week vigorous physical activity | ↑ |
| | p914 | Duration of vigorous activity (min/day) | ↑ |
| | p1070 | TV viewing hours/day | ↓ |
| | p1080 | Computer use hours/day | ↓ |
| | p1090 | Driving hours/day | ↓ |
| | p2296 | Number of falls in past year | ↓ |
| | p3414 | Hip pain lasting ≥3 months | ↓ |
| | p3571 | Back pain lasting ≥3 months | ↓ |
| | p48 | Waist circumference (cm) | ↓ |
| | p90012 | Mean wrist acceleration (mg) | ↑ |
| | p90013 | SD of wrist acceleration | ↑ |
| | p90020 | Sedentary time fraction | ↓ |
| | p90021 | Light physical activity time fraction | ↑ |
| | p90022 | Moderate physical activity time fraction | ↑ |
| | p90023 | Vigorous physical activity time fraction | ↑ |
| | p90029 | MVPA fraction (accelerometry) | ↑ |
| ***Cognition (24 items)*** | | | |
| | p20016 | Fluid intelligence score | ↑ |
| | p20023 | Mean reaction time (ms) | ↓ |
| | p2060 | Frequency of unenthusiasm/disinterest | ↓ |

| Domain | UKB Field | Description | Dir. |
|---|---|---|---|
| | p2070 | Frequency of tenseness/restlessness | ↓ |
| | p2090 | Difficulty concentrating | ↓ |
| | p29002 | Frequency of lack of interest/pleasure | ↓ |
| | p20089 | Self-rated cognitive decline vs. 10 years ago | ↓ |
| | p20088 | Dementia ever diagnosed | ↓ |
| | p5699 | FI10 arithmetic sequence score | ↑ |
| | p20183 | FI10 arithmetic sequence score (alt form) | ↑ |
| | p5790 | FI12 matrix pattern sequence score | ↑ |
| | p20187 | FI12 matrix pattern score (alt form) | ↑ |
| | p26302 | Picture vocabulary (cognitive ability) | ↑ |
| | p26306 | Response delay interval (processing speed) | ↓ |
| | p21625 | Time spent on touchscreen cognitive tasks | ↑ |
| | p62 | Willingness to attempt cognitive tests | ↑ |
| | p10241 | Touchscreen duration (pilot) | ↑ |
| | p120042 | Cognitive symptom severity (composite) | ↓ |
| | p32104 | Forgetfulness/memory problems (past 7 days) | ↓ |
| | p32105 | Poor concentration (past 7 days) | ↓ |
| | p32106 | Trouble expressing thoughts (past 7 days) | ↓ |
| | p32107 | Trouble finding words (past 7 days) | ↓ |
| | p32108 | Slow thinking (past 7 days) | ↓ |
| | p32109 | Trouble solving problems (past 7 days) | ↓ |
| ***Psychological (33 items)*** | | | |
| | p20127 | Neuroticism score (EPQ-R) | ↓ |
| | p1920 | Mood swings | ↓ |
| | p1930 | Irritability | ↓ |
| | p1940 | Outgoingness/sociability | ↑ |
| | p1950 | Miserableness | ↓ |
| | p1960 | Nervousness | ↓ |

| Domain | UKB Field | Description | Dir. |
|---|---|---|---|
| | p1970 | Emotional sensitivity | ↓ |
| | p1980 | Has someone to confide in | ↑ |
| | p1990 | Job stress | ↓ |
| | p2000 | Family relationship stress | ↓ |
| | p2010 | Financial stress | ↓ |
| | p4598 | Never clinically depressed (reverse-coded) | ↑ |
| | p20123 | Probable single-episode major depression | ↓ |
| | p20124 | Probable recurrent MDD (moderate) | ↓ |
| | p20125 | Probable recurrent MDD (severe) | ↓ |
| | p4653 | Frequency of anxiety/worry | ↓ |
| | p29059 | Cannot stop worrying (GAD item) | ↓ |
| | p29060 | Worries too much (GAD item) | ↓ |
| | p29061 | Hard to relax (GAD item) | ↓ |
| | p29062 | Restlessness (GAD item) | ↓ |
| | p2020 | Loneliness/isolation (self-report) | ↓ |
| | p29172 | Lacks companionship | ↓ |
| | p29173 | Feels left out | ↓ |
| | p29174 | Feels isolated from others | ↓ |
| | p29164 | Frequency of video calls | ↑ |
| | p29165 | Frequency of voice calls | ↑ |
| | p29166 | Confides in someone | ↑ |
| | p29175 | Resilience: bounces back from hard times | ↑ |
| | p29176 | Resilience: gets through stressful events | ↑ |
| | p29181 | General happiness | ↑ |
| | p29182 | Life feels meaningful | ↑ |
| | p20501 | Manic episode ever | ↓ |
| | p2030 | Guilty feelings | ↓ |
| ***Sensory (34 items)*** | | | |

| Domain | UKB Field | Description | Dir. |
|---|---|---|---|
| | p2247 | Hearing difficulty (self-report) | ↓ |
| | p10793 | Hearing difficulty (pilot assessment) | ↓ |
| | p2257 | Difficulty hearing in background noise | ↓ |
| | p3393 | Hearing aid use | ↓ |
| | p28627 | Current hearing loss (static) | ↓ |
| | p28628 | Duration of hearing loss | ↓ |
| | p28629 | Extent of hearing loss | ↓ |
| | p28630 | Other current hearing issues | ↓ |
| | p28631 | Duration of other hearing issues | ↓ |
| | p28632 | Extent of other hearing issues | ↓ |
| | p4849 | Hearing test completion | ↑ |
| | p4268 | Hearing test completion: left ear | ↑ |
| | p4272 | Hearing test duration: left ear (ms) | ↑ |
| | p4275 | Hearing test completion: right ear | ↑ |
| | p4279 | Hearing test duration: right ear (ms) | ↑ |
| | p131258 | Date of first H90 (conductive HL) ICD record | ↑ |
| | p131259 | Source of H90 ICD record | ↑ |
| | p131260 | Date of first H91 (sensorineural HL) ICD record | ↑ |
| | p131261 | Source of H91 ICD record | ↑ |
| | p132460 | Date of first Q16 (ear anomaly) ICD record | ↑ |
| | p132461 | Source of Q16 ICD record | ↑ |
| | p2217 | Vision difficulty (self-report) | ↓ |
| | p2207 | Wears glasses or contact lenses | ↓ |
| | p2227 | Other eye problems | ↓ |
| | p28609 | Current vision problems (static) | ↓ |
| | p28610 | Duration of vision problems | ↓ |
| | p28611 | Extent of vision problems | ↓ |
| | p20261 | Age-related macular degeneration (self-report) | ↓ |

| Domain | UKB Field | Description | Dir. |
|---|---|---|---|
| | p20262 | Myopia/short-sightedness diagnosis | ↓ |
| | p5419 | Eye affected by injury or trauma | ↓ |
| | p5430 | Age at vision loss diagnosis | ↓ |
| | p6147 | Reason for wearing glasses/contacts | ↓ |
| | p131212 | Date of first H54 (blindness/low vision) ICD record | ↑ |
| | p131213 | Source of H54 ICD record | ↑ |

**Table S2.** *Comparison of intrinsic capacity (IC) operationalization approaches in UK Biobank. This study implements IC as a continuous functional-reserve construct across 137 items; published UK Biobank studies use a deficit-accumulation approach that dichotomizes each domain into impaired/not-impaired and sums across domains. Both approaches share the same five WHO ICOPE domains (Beard et al. 2016; Cesari et al. 2018) but differ in indicator depth, scaling, and analytic purpose.*

| Feature | This Study | Beyene et al. Maturitas 2024 | Ramírez-Vélez et al. J Cachexia 2023 | Ramírez-Vélez et al. J Affect Disord 2025 |
|---|---|---|---|---|
| **Cohort** | UK Biobank (n = 501,936) | UK Biobank (n = 157,457) | UK Biobank (n = 443,059) | UK Biobank (n = 156,076) |
| **Conceptual framing** | Continuous latent capacity / functional reserve | Deficit accumulation / risk index | Deficit accumulation / risk index | Deficit accumulation / risk index |
| **Theoretical basis** | WHO ICOPE (Beard et al. 2016; Cesari et al. 2018) | WHO ICOPE | WHO ICOPE | WHO ICOPE |
| **Domains** | Locomotion, cognition, psychological, sensory, vitality | Locomotion, cognition, psychological, sensory, vitality | Locomotion, cognition, psychological, sensory, vitality | Locomotion, cognition, psychological, sensory, vitality |
| **Indicators per domain** | 20–40 items per domain (137 total) | 1–2 items per domain (7 total) | 1 item per domain (5 total) | 1–2 items per domain (6 total) |
| **Indicator sources** | Grip strength, accelerometry, gait pace, activity duration; fluid intelligence, reaction time; neuroticism, mental-health items; BMI, sleep, fatigue, blood biomarkers; audiometry, vision self-report | Grip strength; cognitive function; depressive symptoms (PHQ-9); visual acuity; walking pace | Grip strength; fluid intelligence; depressive symptoms; vision difficulty; $FEV_1$ | Grip strength; fluid intelligence; depressive symptoms; vision/hearing difficulty |
| **Indicator scaling** | Direction-harmonized (higher = better capacity), z-scored within domain | Binary threshold: impaired / not impaired | Binary threshold: impaired / not impaired | Binary threshold: impaired / not impaired |
| **Domain score** | Mean of standardized indicators (≥1 item required) | Dichotomous: impaired / not impaired | Dichotomous: impaired / not impaired | Dichotomous: impaired / not impaired |
| **Total IC score** | Mean of five continuous domain scores | Count of impaired domains (ordinal, 0–5) | Count of impaired domains (ordinal, 0–5) | Count of impaired domains (ordinal, 0–5) |
| **Missing data** | Graceful skipping; domain computed if ≥1 item non-null | Complete-case or exclusion | Complete-case | Complete-case |
| **Output type** | Continuous, symmetric, approximately normal | Ordinal integer (0–5) | Ordinal integer (0–5) | Ordinal integer (0–5) |
| **Primary outcome** | Biological age, all-cause mortality (Cox), incident disease (logistic), IC trajectories | All-cause mortality, hospitalization | Incident CVD, CVD mortality | Incident depression |
| **Primary use case** | Biological aging, longitudinal trajectories, genomics | Clinical risk stratification | Clinical risk stratification | Clinical risk stratification |

**Table S3.** *Definition and inventory of the 49 mAge biological subsystems.*

| Subsystem | Category | # | Assignment rule | Overlap | Min. proteins | Clock |
|---|---|---|---|---|---|---|
| Brain | Organ/tissue | 1 | HPA top-expressing tissue (max nTPM); proteins permitted to also map to other tissues | Overlap | 50 | Age + Mortality |
| Blood vessel/Artery | Organ/tissue | 2 | HPA top-expressing tissue; vascular smooth muscle and endothelial enrichment | Overlap | 50 | Age + Mortality |
| Liver | Organ/tissue | 3 | HPA top-expressing tissue; hepatocyte-enriched proteins | Overlap | 50 | Age + Mortality |
| Lymphoid tissue | Organ/tissue | 4 | HPA top-expressing tissue; spleen/lymph node enrichment | Overlap | 50 | Age + Mortality |
| Intestine | Organ/tissue | 5 | HPA top-expressing tissue; small/large intestine enrichment | Overlap | 50 | Age + Mortality |
| Kidney | Organ/tissue | 6 | HPA top-expressing tissue (max nTPM) | Overlap | 50 | Age + Mortality |
| Heart muscle | Organ/tissue | 7 | HPA top-expressing tissue; cardiomyocyte-enriched | Overlap | 50 | Age + Mortality |
| Skeletal muscle | Organ/tissue | 8 | HPA top-expressing tissue | Overlap | 50 | Age + Mortality |
| Lung | Organ/tissue | 9 | HPA top-expressing tissue | Overlap | 50 | Age + Mortality |
| Pancreas | Organ/tissue | 10 | HPA top-expressing tissue | Overlap | 30 | Age + Mortality |
| Adipose tissue | Organ/tissue | 11 | HPA top-expressing tissue | Overlap | 30 | Age + Mortality |
| Bone marrow | Organ/tissue | 12 | HPA top-expressing tissue; haematopoietic enrichment | Overlap | 30 | Age + Mortality |
| Adrenal gland | Organ/tissue | 13 | HPA top-expressing tissue | Overlap | 30 | Age + Mortality |
| Reproductive tissue | Organ/tissue | 14 | HPA top-expressing tissue (testis/ovary/prostate/uterus pooled) | Overlap | 30 | Age + Mortality |
| Retina | Organ/tissue | 15 | HPA top-expressing tissue | Overlap | 20 | Age + Mortality |
| Stomach/Esophagus | Organ/tissue | 16 | HPA top-expressing tissue | Overlap | 20 | Age + Mortality |
| Skin | Organ/tissue | 17 | HPA top-expressing tissue | Overlap | 30 | Age + Mortality |
| Salivary/Endocrine | Organ/tissue | 18 | HPA top-expressing tissue (minor salivary, parathyroid, thyroid pooled) | Overlap | 20 | Age + Mortality |
| Testis | Organ/tissue | 19 | HPA top-expressing tissue (male reproductive only) | Overlap | 20 | Age + Mortality |
| Secreted to blood | Secretome | 20 | HPA secretome class "secreted to blood" | Overlap (secretome) | 50 | Age + Mortality |
| Secreted in brain | Secretome | 21 | HPA secretome class "secreted in brain" | Overlap (secretome) | 30 | Age + Mortality |
| Secreted to ECM | Secretome | 22 | HPA secretome class "secreted to ECM" | Overlap (secretome) | 30 | Age + Mortality |
| Secreted other | Secretome | 23 | HPA secretome class "secreted other" (non-blood, non-ECM, non-brain) | Overlap (secretome) | 30 | Age + Mortality |
| Not secreted (intracellular) | Secretome | 24 | HPA secretome class "intracellular"/"not secreted" | Overlap (secretome) | 50 | Age + Mortality |
| Adipose cells (grouped) | Grouped cell | 25 | HPA cell-type pooled enrichment (adipocytes + progenitors) | Overlap | 30 | Age + Mortality |
| Fibroblasts (grouped) | Grouped cell | 26 | HPA cell-type pooled (raw fibroblasts + stromal subsets) | Overlap | 30 | Age + Mortality |

| Subsystem | Category | # | Assignment rule | Overlap | Min. proteins | Clock |
|---|---|---|---|---|---|---|
| Immune cells (grouped) | Grouped cell | 27 | HPA cell-type pooled (B, T, NK, monocytes, macrophages, neutrophils) | Overlap | 50 | Age + Mortality |
| Endothelial cells (grouped) | Grouped cell | 28 | HPA cell-type pooled (vascular, lymphatic, capillary endothelial) | Overlap | 30 | Age + Mortality |
| Epithelial cells (grouped) | Grouped cell | 29 | HPA cell-type pooled (squamous, glandular, ciliated, secretory) | Overlap | 30 | Age + Mortality |
| Smooth muscle cells (grouped) | Grouped cell | 30 | HPA cell-type pooled (vascular and visceral smooth muscle) | Overlap | 30 | Age + Mortality |
| Mesothelial cells (grouped) | Grouped cell | 31 | HPA cell-type pooled (peritoneal, pleural mesothelium) | Overlap | 20 | Age + Mortality |
| Liver/hepatocytes (grouped) | Grouped cell | 32 | HPA cell-type pooled (hepatocytes + cholangiocytes) | Overlap | 30 | Age + Mortality |
| Cardiomyocytes (grouped) | Grouped cell | 33 | HPA cell-type pooled (atrial + ventricular cardiomyocytes) | Overlap | 20 | Age + Mortality |
| Reproductive cells (grouped) | Grouped cell | 34 | HPA cell-type pooled (germ cells, spermatids, oocytes, granulosa) | Overlap | 20 | Age + Mortality |
| Glia (grouped) | Grouped cell | 35 | HPA cell-type pooled (astrocytes, oligodendrocytes, microglia) | Overlap | 20 | Age + Mortality |
| Adrenal cells (grouped) | Grouped cell | 36 | HPA cell-type pooled (cortical and medullary adrenal cells) | Overlap | 20 | Age + Mortality |
| Fibroblasts (raw) | Raw cell type | 37 | HPA single-cell enriched fibroblast subtype | Overlap | 20 | Age + Mortality |
| Adipose progenitor cells (raw) | Raw cell type | 38 | HPA single-cell enriched adipose progenitor subtype | Overlap | 20 | Age + Mortality |
| Macrophages (raw) | Raw cell type | 39 | HPA single-cell enriched macrophage subtype | Overlap | 20 | Age + Mortality |
| Smooth muscle cells (raw) | Raw cell type | 40 | HPA single-cell enriched smooth muscle subtype | Overlap | 20 | Age + Mortality |
| Cardiomyocytes (raw) | Raw cell type | 41 | HPA single-cell enriched cardiomyocyte subtype | Overlap | 20 | Age + Mortality |
| Enteric glia (raw) | Raw cell type | 42 | HPA single-cell enriched enteric glia subtype | Overlap | 10 | Age + Mortality |
| Early spermatids (raw) | Raw cell type | 43 | HPA single-cell enriched early spermatid subtype | Overlap | 10 | Age only |
| Minor salivary glandular cells (raw) | Raw cell type | 44 | HPA single-cell enriched minor salivary glandular subtype | Overlap | 10 | Age + Mortality |
| Proximal tubular cells (raw) | Raw cell type | 45 | HPA single-cell enriched proximal tubular subtype | Overlap | 20 | Age + Mortality |
| Plasma membrane | Organelle | 46 | HPA subcellular “plasma membrane” annotation | Overlap (organelle) | 30 | Age + Mortality |
| Cytosol | Organelle | 47 | HPA subcellular “cytosol” annotation | Overlap (organelle) | 50 | Age + Mortality |
| Nucleoplasm | Organelle | 48 | HPA subcellular “nucleoplasm” annotation | Overlap (organelle) | 30 | Age + Mortality |
| Vesicles / Golgi apparatus | Organelle | 49 | HPA subcellular “vesicles”/“Golgi apparatus” annotations (pooled) | Overlap (organelle) | 30 | Age + Mortality |

**Table S4.** *ChEMBL drugs targeting cardiomyocyte-clock genes.*

| ChEMBL ID | Molecule Name | Target ChEMBL ID | Gene Symbol | Group |
|---|---|---|---|---|
| CHEMBL4084119 | Liraglutide | CHEMBL1784 | GLP1R | GLP1R |
| CHEMBL414357 | Exenatide | CHEMBL1784 | GLP1R | GLP1R |
| CHEMBL18 | Ethoxzolamide | CHEMBL2095180 | CA14 | CA14 |
| CHEMBL1703 | Metformin Hydrochloride | CHEMBL2363065 | NDUFB7 | NDUFB7 |
| CHEMBL1703 | Metformin Hydrochloride | CHEMBL2363065 | NDUFS6 | NDUFS6 |
| CHEMBL2107841 | Albiglutide | CHEMBL1784 | GLP1R | GLP1R |
| CHEMBL2108027 | Dulaglutide | CHEMBL1784 | GLP1R | GLP1R |
| CHEMBL2108724 | Semaglutide | CHEMBL1784 | GLP1R | GLP1R |
| CHEMBL2108336 | Lixisenatide | CHEMBL1784 | GLP1R | GLP1R |
| CHEMBL3707229 | Tezepelumab | CHEMBL3712931 | TSLP | TSLP |
| CHEMBL4297839 | Tirzepatide | CHEMBL1784 | GLP1R | GLP1R |
| CHEMBL4297517 | Mavacamten | CHEMBL3831286 | MYL4 | MYL4 |

**Table S5.** *Drug targets and effects on aging-related outcomes in diabetic participants.*

| Drug | Target | Outcomes Improved | Clock System(s) | Effect Size |
|---|---|---|---|---|
| Gabapentin | CACNB3 | ↓ gap, ↓ pred. age, ↓ mortality | Fibroblasts, Brain | Very strong (~−2.88 gap) |
| Hydroxyurea | RRM2 | ↓ gap, ↓ pred. age, ↓ mortality | Proteome, Lymphoid | Very strong (~−1.79 gap) |
| Estradiol | ESR1 | ↓ gap, ↓ mortality | Proteome, Epithelial | Strong (~−1.32 gap) |
| Estriol | ESR1 | ↓ gap, ↓ mortality | Proteome, Epithelial | Strong (similar to estradiol) |
| Gabapentin | CACNB1 | ↓ gap, ↓ pred. age, ↓ mortality | Adipose prog., Muscle | Strong (~−0.80 gap) |
| Gabapentin | CACNA1H | ↓ gap, improved survival | Proteome, Intestine | Moderate–strong (~−1.50 gap) |
| Clonazepam | GABRA4 | ↓ gap | Proteome | Moderate (~−1.94 gap) |
| Clorazepate | GABRA4 | ↓ gap | Proteome | Moderate (similar to clonazepam) |
| Insulin | INSR | ↓ mortality | Epithelial cells | Moderate (mortality only) |
| Brinzolamide | CA2 | ↓ mortality | Epithelial cells | Moderate (cell-specific) |
| Amlodipine | CACNA1C | ↑ time-to-death | Intestine | Weak–mod. (~+0.47 TTD) |
| Diltiazem | CACNA1C | ↑ time-to-death | Intestine | Weak–mod. (similar) |
| Felodipine | CACNA1C | ↑ time-to-death | Intestine | Weak–mod. (similar) |

**Table S6.** *Subsystem-specific clock proteins, performance, and optimization parameters.*

| Level | Group | N proteins | N train | N test | $R^2$ (test) | MAE (test, yr) | Pearson r | Best alpha | Best L1 ratio |
|---|---|---|---|---|---|---|---|---|---|
| secretome | Blood | 473 | 42384 | 10597 | 0.7325 | 3.3788 | 0.8559 | 0.0078 | 0.5 |
| secretome | Brain | 31 | 42386 | 10597 | 0.3398 | 5.4638 | 0.5829 | 0.0056 | 0.7 |
| secretome | ECM | 90 | 42385 | 10597 | 0.6238 | 4.0163 | 0.7898 | 0.0048 | 1 |
| secretome | Not secreted | 2105 | 42386 | 10597 | 0.8063 | 2.8614 | 0.8979 | 0.0091 | 0.7 |
| secretome | Secreted (other) | 197 | 42386 | 10597 | 0.5999 | 4.1631 | 0.7746 | 0.0051 | 0.7 |
| tissue | All tissue | 1074 | 42386 | 10597 | 0.6769 | 3.7346 | 0.8228 | 0.0060 | 0.8 |
| tissue | adipose tissue | 40 | 42386 | 10597 | 0.3175 | 5.5828 | 0.5635 | 0.0038 | 0.8 |
| tissue | blood vessel | 73 | 42385 | 10597 | 0.6067 | 4.1099 | 0.7790 | 0.0048 | 1 |
| tissue | bone marrow | 122 | 42386 | 10597 | 0.2681 | 5.8001 | 0.5180 | 0.0059 | 0.6 |
| tissue | brain | 241 | 42386 | 10597 | 0.6444 | 3.9036 | 0.8028 | 0.0046 | 1 |
| tissue | choroid plexus | 31 | 42386 | 10597 | 0.2693 | 5.7645 | 0.5190 | 0.0026 | 1 |
| tissue | esophagus | 61 | 42386 | 10597 | 0.3434 | 5.4334 | 0.5862 | 0.0040 | 0.9 |
| tissue | heart muscle | 44 | 42386 | 10597 | 0.2785 | 5.7780 | 0.5280 | 0.0065 | 1 |
| tissue | intestine | 104 | 42386 | 10597 | 0.4490 | 4.9476 | 0.6702 | 0.0044 | 0.7 |
| tissue | kidney | 39 | 42385 | 10597 | 0.3371 | 5.4123 | 0.5807 | 0.0049 | 0.8 |
| tissue | liver | 210 | 42384 | 10597 | 0.4753 | 4.8039 | 0.6895 | 0.0070 | 0.5 |
| tissue | lung | 32 | 42384 | 10597 | 0.2551 | 5.8351 | 0.5051 | 0.0064 | 1 |
| tissue | lymphoid tissue | 220 | 42386 | 10597 | 0.4965 | 4.7297 | 0.7048 | 0.0051 | 0.6 |
| tissue | pancreas | 52 | 42384 | 10596 | 0.1923 | 6.1013 | 0.4389 | 0.0022 | 1 |
| tissue | parathyroid gland | 33 | 42386 | 10597 | 0.2799 | 5.7468 | 0.5294 | 0.0033 | 0.7 |
| tissue | placenta | 55 | 42385 | 10597 | 0.2603 | 5.8121 | 0.5107 | 0.0053 | 1 |

| Level | Group | N proteins | N train | N test | $R^2$ (test) | MAE (test, yr) | Pearson r | Best alpha | Best L1 ratio |
|---|---|---|---|---|---|---|---|---|---|
| tissue | retina | 32 | 42384 | 10596 | 0.0849 | 6.5784 | 0.2922 | 0.0055 | 1 |
| tissue | skeletal muscle | 88 | 42386 | 10597 | 0.3504 | 5.4121 | 0.5921 | 0.0055 | 1 |
| tissue | skin 1 | 48 | 42386 | 10597 | 0.3038 | 5.6291 | 0.5512 | 0.0050 | 1 |
| tissue | testis | 75 | 42366 | 10592 | 0.2059 | 6.0477 | 0.4540 | 0.0070 | 1 |
| cell_grouped | Adipose cells | 244 | 42386 | 10597 | 0.6438 | 3.8949 | 0.8024 | 0.0048 | 1 |
| cell_grouped | Adrenal cells | 299 | 42386 | 10597 | 0.5215 | 4.5524 | 0.7222 | 0.0053 | 1 |
| cell_grouped | Cardiomyocytes | 89 | 42381 | 10596 | 0.2764 | 5.7606 | 0.5258 | 0.0050 | 1 |
| cell_grouped | Endothelial cells | 213 | 42386 | 10597 | 0.4478 | 4.9597 | 0.6693 | 0.0040 | 1 |
| cell_grouped | Epithelial cells | 253 | 42386 | 10597 | 0.5857 | 4.2144 | 0.7654 | 0.0049 | 0.8 |
| cell_grouped | Fibroblasts | 149 | 42386 | 10597 | 0.6286 | 4.0036 | 0.7929 | 0.0048 | 1 |
| cell_grouped | Glia cells | 34 | 42384 | 10596 | 0.1853 | 6.1012 | 0.4308 | 0.0058 | 0.4 |
| cell_grouped | Immune | 358 | 42386 | 10597 | 0.5876 | 4.2217 | 0.7666 | 0.0071 | 0.5 |
| cell_grouped | Liver cells | 72 | 42386 | 10597 | 0.3271 | 5.5067 | 0.5720 | 0.0081 | 1 |
| cell_grouped | Mesothelial cells | 95 | 42386 | 10597 | 0.3495 | 5.4186 | 0.5913 | 0.0051 | 1 |
| cell_grouped | Reproductive cells | 141 | 42375 | 10594 | 0.2894 | 5.7065 | 0.5380 | 0.0038 | 0.9 |
| cell_grouped | Smooth muscle cells | 118 | 42386 | 10597 | 0.5308 | 4.5190 | 0.7286 | 0.0038 | 1 |
| cell_raw | All cell raw | 403 | 42381 | 10596 | 0.6124 | 4.0917 | 0.7825 | 0.0047 | 1 |
| cell_raw | Adipocytes (Subcutaneous) | 116 | 42386 | 10597 | 0.3582 | 5.3910 | 0.5985 | 0.0073 | 1 |
| cell_raw | Adipose progenitor cells | 128 | 42385 | 10597 | 0.6094 | 4.0532 | 0.7807 | 0.0048 | 1 |

| Level | Group | N proteins | N train | N test | $R^2$ (test) | MAE (test, yr) | Pearson r | Best alpha | Best L1 ratio |
|---|---|---|---|---|---|---|---|---|---|
| cell_raw | Adrenal cortex cells | 241 | 42386 | 10597 | 0.3659 | 5.3628 | 0.6051 | 0.0055 | 1 |
| cell_raw | Adrenal medulla cells | 58 | 42372 | 10594 | 0.4108 | 5.1032 | 0.6410 | 0.0057 | 0.7 |
| cell_raw | Breast glandular cells | 63 | 42386 | 10597 | 0.3584 | 5.3834 | 0.5988 | 0.0077 | 1 |
| cell_raw | Cardiomyocytes | 89 | 42381 | 10596 | 0.2764 | 5.7606 | 0.5258 | 0.0050 | 1 |
| cell_raw | Colon enterocytes | 49 | 42386 | 10597 | 0.2385 | 5.9123 | 0.4886 | 0.0026 | 0.9 |
| cell_raw | Early spermatids | 114 | 42381 | 10596 | 0.2823 | 5.7353 | 0.5314 | 0.0052 | 1 |
| cell_raw | Endothelial cells | 213 | 42386 | 10597 | 0.4478 | 4.9597 | 0.6693 | 0.0040 | 1 |
| cell_raw | Enteric glia cells | 34 | 42384 | 10596 | 0.1853 | 6.1012 | 0.4308 | 0.0058 | 0.4 |
| cell_raw | Fibroblasts | 149 | 42386 | 10597 | 0.6286 | 4.0036 | 0.7929 | 0.0048 | 1 |
| cell_raw | Hepatocytes | 71 | 42386 | 10597 | 0.3241 | 5.5218 | 0.5694 | 0.0094 | 1 |
| cell_raw | Macrophages | 198 | 42386 | 10597 | 0.4843 | 4.7592 | 0.6959 | 0.0071 | 0.5 |
| cell_raw | Mesothelial cells | 95 | 42386 | 10597 | 0.3495 | 5.4186 | 0.5913 | 0.0051 | 1 |
| cell_raw | Minor salivary glandular cells | 64 | 42386 | 10597 | 0.3290 | 5.4759 | 0.5740 | 0.0051 | 1 |
| cell_raw | Neutrophils | 48 | 42384 | 10596 | 0.2354 | 5.9345 | 0.4852 | 0.0035 | 1 |
| cell_raw | Proximal tubular cells | 50 | 42386 | 10597 | 0.3092 | 5.5914 | 0.5561 | 0.0033 | 0.8 |
| cell_raw | Smooth muscle cells | 118 | 42386 | 10597 | 0.5308 | 4.5190 | 0.7286 | 0.0038 | 1 |
| cell_raw | T-cells | 98 | 42386 | 10597 | 0.3322 | 5.5127 | 0.5765 | 0.0038 | 1 |
